# A Study Protocol for CommuniT1D : A Customized Virtual Peer Support Program For People Living With Type 1 Diabetes In Canada

**DOI:** 10.64898/2026.01.30.25336170

**Authors:** Abdoulaye Thiaw, Gaël Précieux Ayihounton, Virtue Bajurny, Aya Benabess, Jacqueline Bender, Denis Boutin, Sasha Delorme, Maman Joyce Dogba, Michaëlla Etienne, Marie Pierre Gagnon, Marley Greenberg, Hina Hakim, Shayla Hele, Elodie Hetu, Korey Hood, Annie LeBlanc, Andréa Limbourg, Dylan MacKay, Matthew Menear, Crescence Joëlle Mefou Tasong, Adhiyat Najam, Ruth Ndjaboue, Justin Presseau, Charles Racine, Renza Scibilia, Peter Senior, Cherise Shockley, Dana Tannenbaum Greenberg, Holly O. Witteman

## Abstract

**Background and objectives:** Type 1 diabetes (T1D) is a largely self-managed condition that requires ongoing daily tasks and decisions. Many people living with T1D in Canada manage it alone, which can feel very isolating and negatively affect physical and mental health. Connecting with other people in similar situations may help to reduce the potential burden associated with managing this health condition. We aim to co-develop and evaluate a virtual peer support program called CommuniT1D, led by people with T1D, to improve the overall wellbeing of people across Canada whose lives are affected by T1D.

**Methods and analysis:** Using a community-based participatory design and action research approach and a realist evaluation framework, we will first co-develop CommuniT1D by working together as a group of people with T1D, researchers, and clinicians. Over two thirds of steering committee members live with T1D (their own or their child’s.) This collective lived expertise is complemented by experts in mental health, social support, health services research, and other relevant fields. Once the program is ready to welcome members, we will work with our partner organizations, networks, and use tailored ads to recruit CommuniT1D peer leaders and members. We will then form virtual peer support groups of people with shared lived experience. Within the program, we will hold monthly small group meetings led by peer leaders via an online platform. We will also hold monthly large webinars open to all CommuniT1D members and other interested people. To evaluate CommuniT1D, we will conduct surveys at baseline and every 6 months, collecting data about diabetes distress, life challenges, quality of life, wellbeing, management indicators, and access and use of management tools and services. We will analyze quantitative data using repeated measures analysis of variance. We will also conduct individual interviews with CommuniT1D members and peer leaders at two time points. We will analyze interview data thematically, and create a logic model by triangulating results from qualitative and quantitative analyses, applying a realist evaluation lens.

**Discussion:** Peer support may help people with T1D feel less alone and better supported. This protocol outlines the design of a virtual peer support program called CommuniT1D to improve the wellbeing of people whose lives are affected by T1D in Canada. We hope that this program will help better equip people with T1D to cope with T1D-related stressors, thus improving the lives of people with T1D in Canada.

## INTRODUCTION

### Background

Type 1 diabetes (T1D) is a chronic autoimmune condition where the immune system attacks the insulin-producing beta cells in the pancreas resulting in the body producing very little or no insulin, a hormone essential to life. [1] In Canada, T1D is estimated to account for 9% in all cases of diabetes. [2] According to a study published in The Lancet Diabetes and Endocrinology, an estimated 285,000 Canadians were living with type 1 diabetes in 2022. [3] This number could rise to over 455,000 by 2040 if current trends continue, with an annual increase of approximately 4.4% in the number of people affected. [3] This observed annual increase reflects a consistent trend over the past two decades. Specifically, between 2000 and 2022, the incidence of T1D rose by 34%, aligning with this growth rate. [4,5] This suggests that the average annual increase has been sustained over at least the last 20 years.

Living with T1D means constantly juggling a number of daily tasks. These multiple tasks typically include regular monitoring of blood glucose levels, administering insulin injections or managing insulin pumps, regulating exercise patterns, measuring and analyzing food, and ensuring nighttime glucose management. [6] Additionally, people with T1D and/or their families may need to regularly source medications and supplies, deal with public or private insurance coverage and its associated paperwork and phone calls [7], attend medical appointments, ensure devices like continuous glucose monitors and insulin pumps remain operational (charging devices, changing infusion sets, replacing sensors, etc.), track glucose trends and make adjustments [8], and seek accommodations if needed within environments such as schools, workplaces, and group activities. All these tasks must be managed alongside all other usual life tasks as well as school, work, family, or other commitments.

The logistical, physical, emotional, psychological, and mental labour of this workload is substantial and can be difficult to understand for those who do not manage the condition. Many people managing T1D in Canada know few or no others in their situation, possibly due to its often invisible nature and relatively low prevalence, especially compared to the much higher prevalence of type 2 diabetes. This means they have few or no people to talk to who truly understand the experience of living with T1D. [9] As noted by participants in Balfe and colleagues’ qualitative study of the experiences of people with T1D, “I don’t think people really understand what’s involved. It’s pretty much on my mind the whole time. There’s no day off from it (Female, 30),” and, “It’s a huge thing. But no one can see it so it’s always there and it’s massive for you but it’s invisible for everybody else (Male, 24).” [10] Previous studies suggest that feelings of loneliness and isolation are common among people with T1D, reaching severe levels in one-fifth of individuals. [9] Such feelings of isolation may be especially pervasive among people going through less common experiences or facing intersectional disparities; for example, living as a racialized person with T1D [11], caring for a very young child with T1D, going through menopause with T1D [12], coping with complications of diabetes, or aging out of parental or provincial insurance coverage. [13] The complexity and chronicity of T1D can similarly have major mental health consequences. Distress due to diabetes has been observed in at least 30% of individuals with T1D [14]and people with T1D have an approximately 10-fold higher risk of developing mental disorders like depressive, anxiety, and eating disorders than individuals without T1D. [15]

Social support is defined as the perceived support available for an individual through social ties with other people, groups, or the community in general. [16] Social support is a broad concept that includes various forms of support, one of which is peer support. Peer support, “could be defined as the giving of assistance and encouragement by an individual considered equal.” [17] The available support could be virtual, implied, imagined, real, transitory and/or continuing. Social support can be delivered in many different forms, including instrumental support (e.g., bringing someone a juice box to treat a low blood sugar), emotional support (e.g., listening to the person, making them feel less alone, reassuring the person), informational support (e.g., sharing ideas, resources), financial support (e.g., paying for supplies, filing paperwork, dealing with insurance), provision of care and social connections to others. [18] In a systematic review examining the relationships between social support and diabetes self-management in adults with T1D or type 2 diabetes, greater perceived social support was significantly associated with improved self-management. [19] Among young adults with T1D, high parental support is associated with better treatment adherence, fewer depressive symptoms, and reduced risky behaviors (e.g., cigarette smoking, heavy episodic alcohol use), while also buffering the negative impact of peer conflicts on blood glucose control. [20] Longitudinal studies further demonstrate that family support during adolescence predicts better psychosocial well-being and blood glucose control in adulthood. [20,21] Additionally, social support helps alleviate the negative effects of diabetes distress and depressive symptoms on disease self-management and can significantly predict health-related quality of life. [22] Lack of social support increases distress and depression among people with T1D, which in turn leads to withdrawal and less social support, creating a vicious cycle. [23] In other words, positive social support including emotional, instrumental, and informational support can influence well-being through a variety of pathways, for example, by improving health behaviors, enhancing self-esteem with regard to performing self-care activities, and reducing distress. [24]

Peer support is an important form of social support, particularly for adults who are coping with their own T1D or that of a loved one. [25,26] In a recent systematic review on technology-mediated peer support interventions for children living with T1D, participants reported feeling emotionally supported, more confident in managing T1D and better equipped to overcome diabetes related challenges. [27] Adults with T1D have similarly reported that peer support from others with T1D alleviates the, “burdensome feeling of diabetes-specific loneliness.’’ [25] Peer support has been shown to significantly benefit parents of children with type 1 diabetes by reducing stress, improving coping strategies, and fostering confidence in diabetes management. For instance, interventions such as peer coaching programs have provided emotional reassurance and practical advice, leading to decreased parental distress and improved glycemic control in children. [28,29] Additionally, online peer support platforms enable parents to connect with others facing similar challenges, offering a sense of community and shared experiences that alleviate feelings of isolation. [30,31] While potentially beneficial, peer supports for T1D appear to be largely inaccessible to people living in Canada. The need to enhance access to virtual peer supports was noted in the Framework for Diabetes in Canada published by the Government of Canada on October 5, 2022. [32]

We therefore aim to co-develop and evaluate CommuniT1D, a program of customized digital health peer support delivered via virtual meetings, in which people managing T1D in Canada can connect with others like them across the country, learn from peers and other knowledgeable people, and receive emotional and informational peer support. [33] With the increased uptake of virtual platforms in recent years, virtual meetings using tools like Zoom, Teams, or Google Meet are increasingly common and accessible. [34] These platforms also offer mechanisms for people to join meetings by phone if they do not have sufficient internet access, which may be the case for people in Canada who live in rural or remote regions where bandwidth may be limited and/or who live with limited financial resources. [35]

### Objectives and hypotheses

This protocol outlines the design of a digital health program called CommuniT1D (*CommunauDT1* in French) for peer support among people in Canada whose lives are affected by type 1 diabetes, be it their own T1D or that of a loved one. Our overall objective is to develop and evaluate a sustainable platform, led by people with T1D, to improve well being, lower diabetes distress and help people living with T1D feel less alone.

We hypothesize that members of CommuniT1D will report reduced diabetes distress (primary outcome), improved quality of life/wellbeing and may have improved physical/mental health outcomes related to diabetes (secondary outcomes). We also postulate that these effects will be modulated by mechanisms such as shared experience, a sense of validation and the ability to adapt to individual contexts. [36,37]

Our **specific aims** are to:

1. Create CommuniT1D, a digital health program consisting of: a) a community of practice of peer leaders living with T1D to moderate virtual peer support meetings, b) a network of virtual peer support groups composed of people with T1D who also share other characteristics or situations (e.g., preferred language, ethnocultural group, stage of life), and c) a bank of other experts who can provide additional support to those living with T1D by sharing relevant knowledge in meetings and webinars.
2. Assess the extent to which participation in CommuniT1D may influence diabetes-related distress and mental and physical health outcomes via mixed methods including repeated surveys and interviews.
3. Determine which aspects of CommuniT1D work or don’t work, for whom, and why.
4. Ensure CommuniT1D’s long-term sustainability.

## METHODS

### Study design

This is a community-based participatory design and action research project. [38] Our steering committee is composed of a majority of people living with T1D: 10 members of the steering committee out of 14 members live with their own and/or their child’s T1D. Led by this committee, we will work with the T1D community in Canada and subgroups within it to design and implement research that aims to improve the lives of people living with T1D.

As recommended in the literature [39], we will explicitly include human-centered design strategies, specifically, transdisciplinarity and rapid iteration [40], to further enhance the effectiveness of our approach. This means that our plans below may shift along the way in response to the needs, preferences, and ideas of CommuniT1D members.

We held a kickoff meeting with all team members to discuss initial steps, timelines, roles and responsibilities. Throughout the project, the steering committee will meet monthly to discuss project tasks, new issues that have arisen, and questions that need the group’s consideration, seeking always to optimize the experiences of participants at every step of the project. The full team will meet quarterly to have broader discussions among the larger group. We formed an international advisory panel consisting of 4 people who live with T1D and have expertise in peer support and/or mental health in T1D. This group will meet every six months with the project lead (HOW) and steering committee members who wish to attend. International panel members will provide advice from their expertise and experiences in other countries. Throughout these meetings, we will review the overall program plan and solicit feedback about pitfalls we should avoid and ideas we could use to ensure the success of the program.

In parallel to our work to develop and evaluate CommuniT1D, we also plan to conduct a scoping review of peer support in T1D. [41] In addition to helping us put our study in context, we anticipate that we may draw ideas and lessons learned from other peer support interventions around the world, and potentially integrate some of them into CommuniT1D.

### Study Setting

This study is a digital health program designed to facilitate support meetings online. In other words, we will conduct our research activities on online platforms such as Zoom or other similar platforms selected by participants (e.g., Google Meet, Teams, WebEx). Throughout the study, we will collect data through online questionnaires and virtual interviews.

### Ethical Approval

This project has been approved by the Research Ethics Committee of the CIUSSS Capitale-Nationale, Approval No. 2024-2887. Consent to participate in the research study will be obtained from participants prior to completing the baseline survey. The participants may withdraw consent at any time throughout the course of the study. Participants who agree to participate in the research study will have the option to be entered in a draw for a CAD$250 gift card each time they complete a survey. All survey data will be deidentified before sharing results.

### Intervention description and design

People will join CommuniT1D through an online sign-up questionnaire available in English or French. In the questionnaire, people will answer questions about themselves and their relationship to diabetes, then they will select and rank up to six attributes they deem the most important for grouping them with others. For example, if someone indicates that the most important attribute for them is to be grouped with other parents of teenagers with T1D, that will be the priority item for placing them in a group. Similarly, if someone indicates that they would prefer to be grouped with people whose faith is an important part of their lives, that would be the priority item. The sign-up questionnaires are available in online appendix 1.

After allowing enough time to gather a critical mass of CommuniT1D members, we will form initial groups, using a clustering algorithm to start the grouping process, then using human judgment to complete the process. Starting the process with an algorithm will allow us to develop an efficient way to enable future group formation as CommuniT1D grows. We will group people based on their similarities to one another and their expressed priorities. For example, a group might be proposed for 5 people who all indicated that they speak French, can meet on weekday evenings, and are interested in talking with other parents of teenagers with T1D. The study coordinator would use that information to begin contacting potential group members about proposed groupings.

Each group will be led by one or two volunteer peer facilitators. When people sign up for CommuniT1D, they will be offered the option to indicate whether they are willing to be a peer facilitator for a group. Peer facilitators will be offered orientation to the role (including but not limited to: program ground rules, expectations for peer facilitators, where to turn for help, options for getting groups started such as ice-breaker or getting-to-know-you activities, and strategies for effective group discussions), access to a listserv of other peer facilitators, and ongoing support from CommuniT1D staff. The orientation program will be described separately in another manuscript but briefly, it consists of a short web page including videos and text, an interactive quiz, and a webinar with CommuniT1D staff and steering committee members who can answer questions. The orientation will be specific to CommuniT1D and informed by training offered in similar programs [42] guidelines for the Practice and Training of Peer Support issued by the Mental Health Commission of Canada [43] and other best practices. [44] We will offer funding to peer leaders who wish to pursue formal certification as a peer leader through Peer Support Canada [45] but will not require certification if a peer leader does not wish to devote the time to that additional process.

Once the groups are formed and peer facilitators identified, monthly meeting sessions will be held. These sessions will be relatively open, allowing each group to discuss relevant topics based on their common needs and interests. The discussion topics will be related to T1D and will adhere to the preliminary ground rules established collectively by the CommuniT1D steering committee and larger research team (see Box below).

#### Box 1. CommuniT1D Community Guidelines

1. **Mutual respect.** We are a diverse community and CommuniT1D is a welcoming and inclusive space. We respect everyone’s backgrounds, social identities (e.g., culture, education level, ethnicity, gender, language, pronouns, religion, appearance, etc.), diabetes management choices, and other aspects of our lives. We welcome people as they are and encourage everyone to include their pronouns in their on-screen name if they wish (e.g., he/him, she/her, they/them, she/they, etc.) We show respect through our words and attitudes towards other group members. **This is our core principle. Many rules below offer specific examples of how we enact mutual respect.**
2. **No medical advice.** We are careful not to give medical advice. It’s ok to tell someone else, “XYZ worked for me,” or, “I’ve heard that XYZ can be helpful in that situation.” It is not ok to tell someone else, “You need to do XYZ.” We also always keep in mind that what works for someone may not work for everyone.
3. **No judging others’ food, blood sugars, A1c, management choices, or life decisions.** Everyone is welcome and respected in CommuniT1D, no matter how they choose to eat, what their target blood sugars or A1c are, how they choose to manage diabetes, or other life decisions. We reject diabetes stigma in all its forms and we avoid stereotypes of people with any kind of diabetes.
4. **Respect preferences about numbers.** T1D involves a lot of numbers; for example, A1c, blood glucose, time in range, and units of insulin. Even though we know the numbers can vary due to variations in labs, technology, genetics, hormones, and body chemistry, these numbers can still feel like a report card. Some people find discussions about these numbers useful while others find such discussions difficult. Each CommuniT1D group will determine how and whether they want to talk about numbers through regular anonymous polls.
5. **Discussions are confidential.** We don’t take screenshots nor do we repeat stories, names, or other details outside of our meetings.
6. **We are careful when sharing details about our loved one(s).** If you have an adult loved one with T1D, please seek their consent before sharing any of their health details. If you have a child loved one with T1D who is old enough for such a discussion, please take the opportunity to have an age-appropriate discussion with them about you talking to others about their T1D. When experiencing difficulties with a loved one with T1D, the focus should be on your need for support (e.g., “How can I cope with this?” “What should I keep in mind in this situation?”)
7. **Build and maintain trust by turning cameras on when possible.** We build trust by keeping cameras on when we can, at least at the beginning of a meeting. Some people may be unable or unwilling to keep their camera on due to poor internet connection or for other reasons. If they can, it may be helpful to turn the camera on briefly at the beginning to say hello and/or post in the chat.
8. **Everyone gets a turn to talk and decides for themselves what they are comfortable sharing.** We make sure everyone has a chance to contribute to the conversation and share their story. If people don’t want to talk, they can always pass on their turn to talk. Peer facilitators will check in with people who never or rarely participate to make sure they feel welcome to share with the group.
9. **Take care with potentially divisive topics; avoid group members feeling excluded**. CommuniT1D is focused on mutual support for everyone affected by T1D. People may have varied experiences with and opinions on topics like politics, current events, etc. Although people’s lives may be affected by these topics, it does not serve our goals to make anyone feel excluded or unwelcome. For topics that may be relevant to T1D but also potentially divisive (for example, government decisions about coverage of medications and supplies) please be sensitive to diverse views, don’t assume you know everyone’s views, and avoid making individuals feel excluded from the group. Groups should check in using anonymous voting to make sure that everyone is comfortable with the topic of discussion and that everyone continues to feel welcome, even if their opinions differ from those of others in the group. We always aim to show empathy, compassion, and mutual respect.
10. **Mistakes happen.** We give each other the benefit of the doubt when we occasionally mess up on these rules. **However, repeated rule violations are not ok.** Repeatedly ignoring the rules will result in being removed from the group.
11. **No selling, donating, or trading devices, supplies, or services.** While we value the community spirit that often motivates such activities, CommuniT1D cannot provide a platform for them. Please keep such activities out of the group.
12. **Email** **if you need help.** If something in the group has made you uncomfortable and you would like to reach out to CommuniT1D staff, please email us at.

In addition to small group meetings of people with T1D, we will also organize larger webinars and question & answer sessions approximately monthly or bi-monthly with guest speakers to present about issues relevant to mental health and T1D, and offer resources to share with community members. Webinars will be held in English, French, or other languages depending on speakers. We will live-caption webinars in English and French. Captioned webinars will be recorded and archived on the CommuniT1D website. We anticipate that potential webinar topics may include: T1D, depression and anxiety [46]; strategies for better sleep with T1D [47]; resources for avoiding or recovering from disordered eating in T1D [48]; meditation and mindfulness as skills and tools for your health [49]; financial help available in different provinces/territories for people with T1D [50]; managing work or a career with T1D; caring for your teen with T1D [51]; strategies for building exercise into life with T1D [52] and more.

### Participants

#### Eligibility Criteria

##### Inclusion Criteria

Participants in CommuniT1D must be people living with T1D or another rare, insulin-requiring form of diabetes, namely, Latent autoimmune diabetes of adults (LADA), Maturity-Onset Diabetes of the Youth (MODY), or type 3c, which occurs due to an injury to or removal of the pancreas. Loved ones such as a parent, child, spouse, caregiver, or friend of an individual with T1D or other rarer forms of diabetes will also be eligible to participate in CommuniT1D. Participants must be 18 years and older, reside in Canada, and be able to read the consent form in English or French.

##### Exclusion Criteria

Individuals with type 2 diabetes, pre-diabetes, or gestational diabetes and no connection to T1D, LADA, MODY, type 3c, or other rare, insulin-requiring forms of diabetes will not be included in CommuniT1D. Individuals under 18 years old will also be ineligible to participate. As the program grows, we hope to expand to also include adolescents and children, who may need parental consent for them to join, depending on their Canadian province or territory of residence. Should we be able to pursue this option, we will work with institutional research ethics boards to ensure that we welcome participation by youth while also ensuring their safety within CommuniT1D. In such a case, the age restrictions in our eligibility criteria would change.

#### Recruitment

##### Strategies for Recruitment and Retention

We will recruit potential participants through partner organizations of people with T1D in Canada (e.g, Diabetes Action Canada, Breakthrough T1D), social media (e.g, Facebook, Instagram) and advertisements in pharmacies and clinics. Specifically, we will send out invitations via such organizations, ask peer leaders to share invitations in their circles, put targeted ads on social media platforms and work with our networks of health professional colleagues (e.g., endocrinologists, family physicians, dietitians) to put up posters or ePosters in their clinics, and work additionally with our networks of pharmacist colleagues to develop and implement recruitment strategies with pharmacies across Canada. We will specifically aim to recruit via pharmacies (both in-person and pharmacy delivery services) because people with T1D who are not otherwise engaging with the health care system must at least be purchasing insulin to stay alive.

Invitations will direct potential CommuniT1D members to referral websites or an email address for a bilingual study coordinator. The referral website will use a simple form to collect basic information, including name, age, province or territory of residence, relationship(s) to T1D (e.g., “I have T1D myself, I am a caregiver to a child with T1D, I am a caregiver to an adult with T1D”), time since diagnosis, language(s) spoken, and other relevant information. Appendix 1 shows the full sign-up form in English and French. Based on previous answers, the sign-up form will ask people to identify and then rank by importance up to six characteristics on which they would like to be grouped with others (e.g., being a parent of a very young child with T1D, living as a racialized person with T1D, being a spouse of a person with T1D, living with T1D while also participating in competitive sports, going through menopause with T1D, etc.) Immediately after completing the sign-up form, prospective CommuniT1D members will be offered the option to join the associated research study. If they indicate interest in doing so, they will read the informed consent form and decide whether or not to participate. Those who decide to participate at that point will then have the option to complete their first study questionnaire immediately, or be reminded by email to complete them later. The study questionnaire is shown in Appendix 2.

All prospective CommuniT1D members will receive an email response from the study coordinator welcoming them and asking them to complete informed consent to take part in the research project, if they didn’t already do so following completion of the sign-up form. Prospective members will be allowed to join CommuniT1D even if they are not participating in the research project, but will not have access to activities that are specific to the research project (e.g., interviews discussing what is working or not for them in CommuniT1D.) The rapid response will also let them know that they will hear back by a given date about options to join a group or multiple groups.

##### Participant Withdrawal or Termination

Participants in the research study associated with CommuniT1D may withdraw at any time by contacting the study coordinator. Their individual-level data will be destroyed at that point. Data already reported in manuscripts or datasets will remain reported.

As for participants in the CommuniT1D program, we wish for CommuniT1D to be an inclusive program where every member feels respected, supported, valued and welcome. To achieve this, the steering committee will continually co-design the guidelines of CommuniT1D meetings, informed by feedback from CommuniT1D members and other team members. The rules will be posted on the CommuniT1D website and peer facilitators will post the bolded items with a link to the full rules in the chat at the beginning of each meeting. Participants who do not adhere to these rules may be subject to exclusion for the program.

### Outcomes

#### Primary outcome

Our primary outcome will be diabetes distress. We are specifically measuring diabetes distress as our primary outcome rather than, for example, depression, because it is specific to diabetes and more predictive of other relevant health outcomes in the T1D population than depression measures. [53–55]

We will use existing adaptations for Type 1 Diabetes Distress Scale [56] and work with our steering committee to adapt the scale in order to address specific life situations relevant to our participants. For example, for CommuniT1D members who are employed, we will add two additional questions about work-related diabetes distress. [57] For parents of teens with T1D, we will use the Parent Diabetes Distress Scale (PARENT-DSS) [58] and will adapt this scale for parents of younger children with T1D. For partners of individuals with T1D, we will use the Partner Diabetes Distress Scale (PARTNER-DDS). [59] Sample items from each of these scales are shown in Table 1 below.

**Table 1.** Primary outcomes and sample items.

| Scale | Sample item(s) |
| --- | --- |
| Type 1 Diabetes Distress Scale. [56] | *Thinking back over the past month, please indicate the degree to which each of the following may have been a problem for you.*<br>Feeling that I've got to be perfect with my diabetes management. |
| Additional questions about work-related diabetes distress. [57] | *Which of the following issues are currently a problem for you?*<br>Worrying about your ability to do your job due to your [your child's] diabetes. |
| Parent Diabetes Distress Scale (PARENT-DSS). [58] | *During the past month, I have been*<br>Feeling that no one notices that diabetes is hard on me, not just on my teen. |
| Parent Diabetes Distress Scale (PARENT-DSS) adapted for younger children | *During the past month, I have been*<br>Feeling that no one notices that diabetes is hard on me, not just on my child. |
| Partner Diabetes Distress Scale (PARTNER-DDS) | *During the past month, I have been*<br>Feeling that no one notices that diabetes is hard on me, not just on my partner. |

#### Secondary outcomes

We will assess secondary outcomes using a series of validated instruments, selected to capture psychosocial, behavioral, and clinical aspects relevant to people living with T1D and their caregivers.

For participants who identify as caregivers of a person living with T1D, we will use Kingston Caregiver Stress Scale. [60] We will measure symptoms of anxiety and depression using the Hospital Anxiety and Depression Scale (HADS), which captures the emotional distress frequently associated with chronic conditions such as T1D. [61] To assess unmet needs for care, we will use selected items adapted from the Canadian Community Health Survey (CCHS). These items will allow us to identify perceived gaps in access to appropriate and timely diabetes-related care. [62] We will evaluate diabetes-specific self-efficacy using the Confidence in Diabetes Scale, which measures participants’ confidence in their ability to perform essential diabetes self-care activities. [63]

To examine fear of hypoglycemia, we will use the short form of the Hypoglycemia Fear Survey II, a validated tool that captures behavioral and emotional responses related to hypoglycemic episodes. [64] We will assess general psychological well-being with the WHO-5 Well-Being Index. This brief instrument has demonstrated high sensitivity for detecting depressive symptoms and is validated in populations with T1D. [51] We will measure diabetes-specific quality of life using the Audit of Diabetes-Dependent Quality of Life (ADDQoL). [65] This instrument assesses the perceived impact of diabetes on various life domains, accounting for personal values and priorities. To capture perceived social support, we will use the Social Provisions Scale [66], which includes items reflecting the availability of guidance, reassurance, and a sense of belonging.

We will measure self-compassion using the Self-Compassion Scale-Short Form (SCS-SF). [67] This scale assesses how individuals treat themselves during challenging experiences. We will use the Ten-Item Personality Inventory (TIPI) to describe participants’ personality traits. [68] While not a direct outcome measure, this tool will allow us to explore how individual personality profiles may moderate the effects of the intervention, and may also capture shifts in constructs such as conscientiousness, which may be useful for diabetes management.

To assess broader outcomes of participating in CommuniT1D, we will use the CommuniT1D check-in measure, a custom tool developed to evaluate emotional well-being, engagement, and the perceived benefits of participating in a peer support initiative. For participants who identify as caregivers of a person living with T1D, we will use the Caregiver check-in, another custom tool to assess caregiver workload. We will also assess health care and access by measuring how easily participants or their family members can access diabetes-related health services and resources. For participants with T1D or caregivers of children with T1D, we will additionally assess diabetes management behaviors through self-reported practices and monitoring routines.

To complement self-reported outcomes, we will collect physical health indicators. Participants will report the number of severe hypoglycemia episodes experienced over the study period. They will also report hemoglobin A1c values and time in range (for those using continuous glucose monitoring or similar technologies). Finally, we will monitor participation in the CommuniT1D program, including attendance at monthly peer meetings, engagement in webinars, and withdrawal where applicable. These participation metrics will inform program reach and fidelity.

For all measures, whenever a French Canadian version of a measure did not already exist, we have translated, back translated, and culturally adapted the scale, using established best practices of dual forward and back-translation. [69]

All measures except those whose licensing agreement precludes sharing them publicly are shown in online appendix 5. Sample items from each of these scales are shown in Table 2 below.

**Table 2.** Secondary outcome measures and sample items.

| Scale | Sample item(s) |
| --- | --- |
| Kingston Caregiver Stress Scale | To what extent are you having feelings of being overwhelmed, overworked, and/or overburdened? |
| Hospital Anxiety and Depression Scale (HADS) | [T]ick the box that comes closest to how you have been feeling in the past week: I get a sort of frightened feeling as if something awful is about to happen |
| Unmet needs for care (adapted from Canadian Community Health Survey (CCHS)) | During the past 12 months, was there ever a time when you felt that you needed care for your diabetes but didn't receive it? |
| The Confidence in Diabetes Scale | I believe I can keep my blood glucose in the normal range when under stress. |
| Short form of the Hypoglycaemia Fear Survey II | Because my blood sugar could go low, I worried about passing out in public. |
| WHO-5 Well-being Index | [I]n the last two weeks: I have felt active and vigorous. |
| ADDQoL | If I did not have diabetes, my quality of life would be: very much better, much better, a little better, the same, worse. |
| Social Provisions Scale | I feel part of a group of people who share my attitudes and beliefs. |
| Self-Compassion Scale Short Form (SCS-SF) | When I feel inadequate in some way, I try to remind myself that feelings of inadequacy are shared by most people. |
| Ten-Item Personality Inventory | Please indicate the extent to which you agree or disagree the traits apply to you: Dependable, self-disciplined. |
| CommuniT1D check-in measure | Participating in CommuniT1D is helping me to: Feel less alone. |
| Caregiver check-in | In the past 6 months, have you done any of the following things to help your loved one manage their diabetes: Brought them something when their blood sugar is low. |
| Health care & access | Do you have (Does your child have) one or more health professionals (doctors, nurses, dietitians, pharmacists, etc.) who provide regular T1D care for you (your child)? |

#### Sample Size Determination

We will estimate a sample size to ensure adequate statistical power to detect a clinically meaningful difference on the Diabetes Distress Scale among study participants. We will select a standardized effect size of 0.4, corresponding to a moderate effect according to Cohen’s criteria. [70] This value is commonly used in studies involving diabetic populations that evaluate psychosocial and behavioral interventions using the Diabetes Distress Scale. [71] Accordingly, we will consider a minimum mean difference of 0.4 units on the 6-point Diabetes Distress Scale clinically relevant in this context. This effect size corresponds to an R² value of 0.3, based on the formula f² = R² / (1 – R²). We will set the alpha level at 0.05. We will assume no clustering effect, as participants may be involved in multiple groups within the CommuniT1D program. The baseline measurement, taken prior to participation in CommuniT1D, will serve as the reference value.

We aim to achieve a minimum statistical power of 80%, using a model that includes five predictor variables, including time and relevant covariates. Based on these parameters (R² = 0.3, five predictors, 80% power, and an alpha of 0.05), the estimated minimum required sample size is 100 participants, as determined using standard sample size estimation procedures for multiple regression models. [72]

### Quantitative data collection

As new CommuniT1D members join, if they provide informed consent and can complete a form in English or French, they will be invited to join the research study evaluating CommuniT1D and answer a series of questionnaires before participating in a meeting to establish baseline measures that will then be re-assessed every 6 months. These questionnaires will focus on outcomes measurements outlined above. Every six months after the first baseline measurements of outcomes, we will collect repeated measurements of the same outcomes. We will continue collecting data every six months for each CommuniT1D member, using automated questionnaire emails. When a CommuniT1D member has not clicked on the questionnaire link one week after it was sent, the study coordinator will reach out to ensure they still wish to remain in the study. The study coordinator will resend the link if the person did not receive it for whatever reason.

Starting with each person’s second repeated questionnaire, each time they complete a questionnaire, study participants will be offered an optional report of their responses, generated automatically by Qualtrics survey software. We believe that for those who are interested, being able to see their responses and potentially examine how and whether their responses have changed over time may provide useful or at least interesting information without causing undue influence on subsequent responses 6 months later.

### Qualitative data collection

Nine to twelve months after the first meetings took place, we will conduct a first set of semi-structured, audio-recorded interviews with CommuniT1D members (including peer facilitators) lasting approximately 60 minutes. The interviews will explore participants’ perspectives on the influence of CommuniT1D on their mental and physical health outcomes (specific aim 2) and on the aspects of the program that have worked well (or not) and why (specific aim 3). We will recruit 30 CommuniT1D members (20 who are not peer facilitators and 10 who are peer facilitators), using maximum variation sampling and limiting interviewees to maximum 2 per group. We will also seek to interview about 4 people who joined CommuniT1D and withdrew from participating, but did not formally withdraw from the research study, to better understand what aspects of CommuniT1D fail to work for people in a given context.

This qualitative data collection phase will follow a realist evaluation approach. [73] Based on previous work [36,37], we have developed an initial program theory for CommuniT1D that will be refined using data from the questionnaires and from participant interviews. According to our initial theory, peer supports, when delivered from peers in similar situations, guided by sufficiently-skilled facilitators, and within life contexts that allow for small or large changes, activate psychological (e.g., sense of connection, emotional validation) and behavioral (e.g., better T1D management strategies and coping strategies) mechanisms, thus leading to positive outcomes such as reductions in diabetes distress and improved quality of life and well-being. The key elements of our initial program theory, represented as a Contexts-Mechanisms-Outcomes model, are presented in Table 2.

**Table 2.**
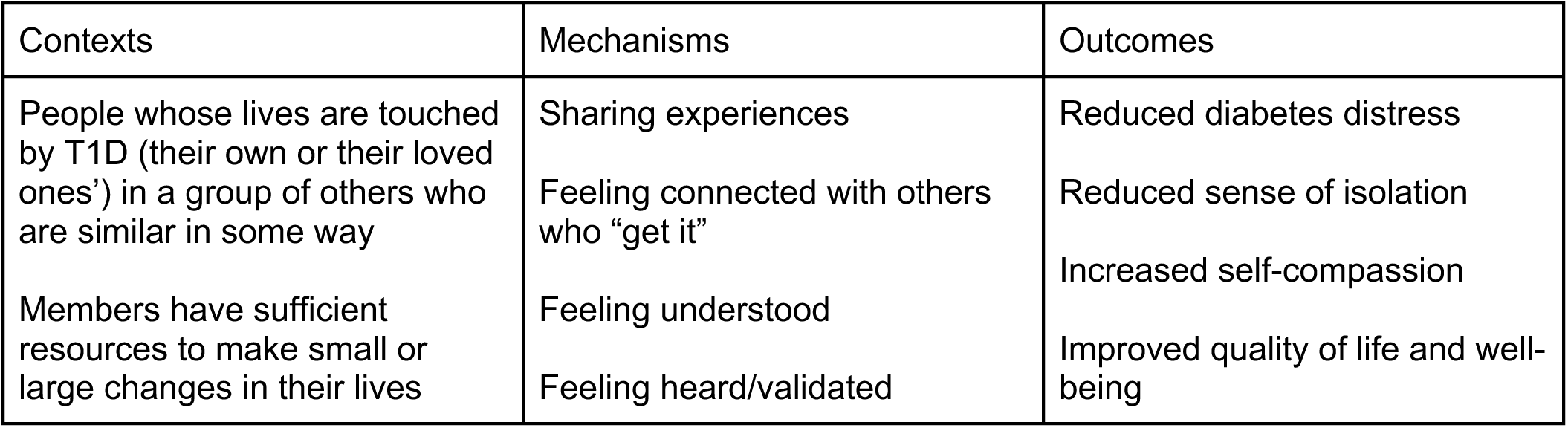
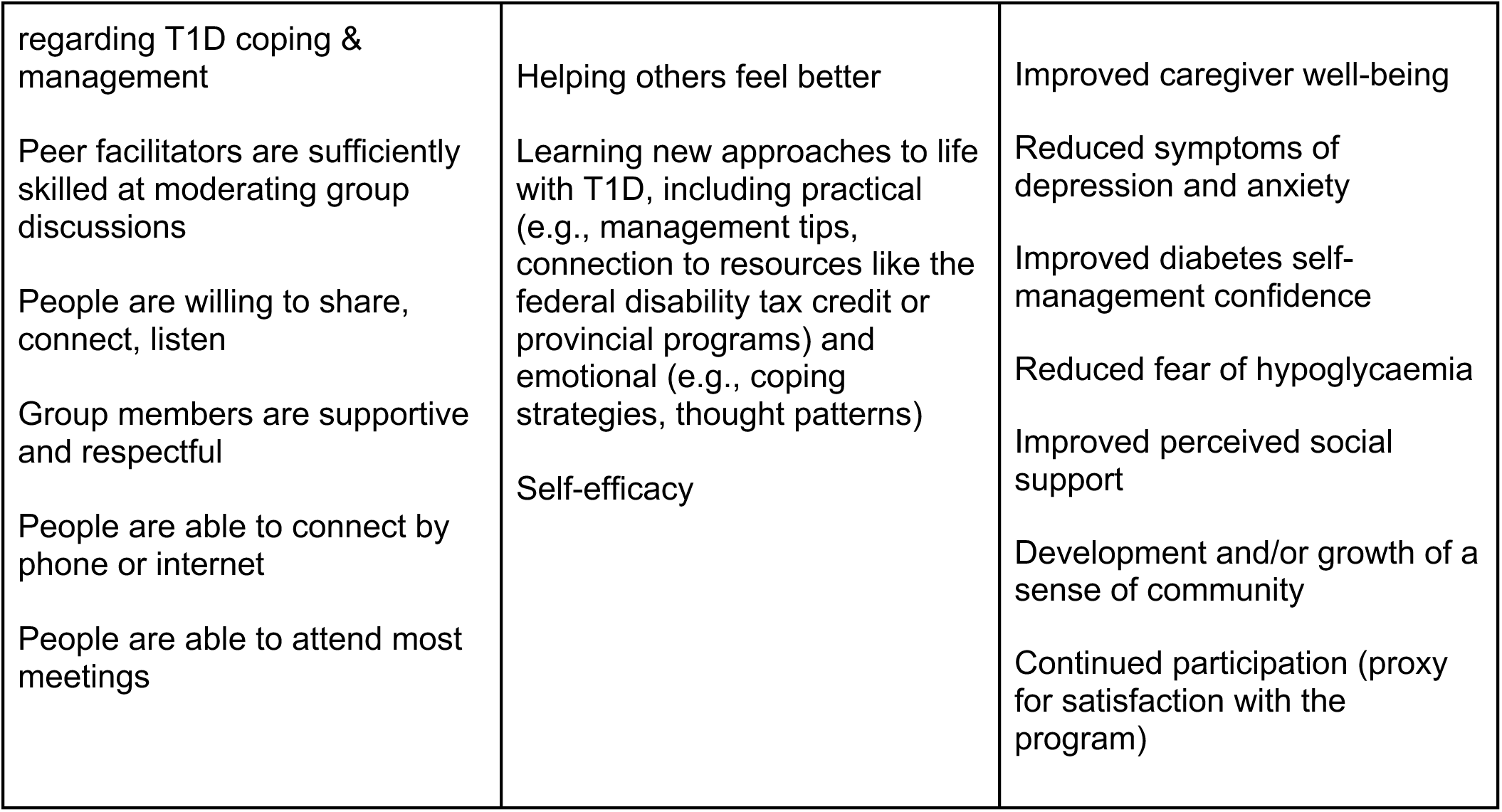
Initial Contexts-Mechanisms-Outcomes model.

Our qualitative interview guide will be co-developed with the steering committee and a sample of CommuniT1D peer leaders and national/international experts. We anticipate asking participants questions about their personal history with T1D, why they joined CommuniT1D, the extent to which CommuniT1D has or has not enabled them to form meaningful connections with others in similar situations, the perceived impact of the program on their mental and physical health, what has worked well for them in CommuniT1D, why it worked well, what has worked less well and why. We will also explore with them how CommuniT1D may or may not have influenced their experience of living with T1D since they joined, and ask questions about their use of CommuniT1D and other resources to help us thoughtfully start planning for sustainability with the needs and views of community members in mind. Interviews will be conducted by the bilingual study coordinator and a trained, bilingual trainee. Interviews will be conducted using the same platform as CommuniT1D meetings or by telephone, according to participants’ preferences and logistical constraints. Interviewers will record field notes after each interview.

Approximately six to twelve months after the first set of interviews, we will explore whether conducting a second set of interviews is likely to further refine our program theory. We expect that further interviews will prove useful and will therefore seek to interview approximately 30 people. We anticipate recruiting 20 CommuniT1D members and 10 CommuniT1D peer facilitators who had not been interviewed last time, and reinterviewing 10 members and 5 CommuniT1D peer facilitators who participated in the first set of interviews. We anticipate also again seeking to interview 4 people who joined CommuniT1D and withdrew from participating, but did not formally withdraw from the research study. We will use the same or similar methods as in the first set of interviews, and will also specifically ask people who are being interviewed for a second time to answer questions about what might have changed from their first interview.

Ultimately, throughout our qualitative data collection, we hope to interview participants who might shed light on the evolving program theory. While we plan for given numbers of interviewees as detailed above, the final number will be determined by theoretical saturation. [74]

### Data management

All the information we collect will be anonymous, or, in the case of qualitative data, will be anonymized to facilitate analysis and data sharing. We will use data for research purposes only. In other words, we will not present individually identifying information in reports, publications or presentations. We will present all quantitative data in aggregated form without individual identifiers. We will present selected verbatim quotes from interviews in anonymized forms and will validate presentations of quotes with the people whose words are being quoted. We will store data on Qualtrics servers located in Canada and on university-based servers located in Canada. We will restrict access to data to research team members who have completed relevant research ethics training. We will keep all data for ongoing monitoring and evaluating purposes, then destroy it 7 years after the end of the project.

When agreed to by study participants, we will deposit anonymized data in Boréalis, the data repository associated with Université Laval. In this way, we will make the data available to the larger scientific community under an open access license.

### Data Analysis

#### Quantitative Analyses

To help us estimate potential effects of participation in CommuniT1D, we will analyze data about diabetes physical and mental health outcomes. We will begin conducting analyses after 1 to 2 years of group meetings, including data from all CommuniT1D members who have been active for at least 6 months and have therefore completed both the baseline questionnaire and one subsequent questionnaire. This means that all people whose data will be analyzed will have between 2 and 4 measures of each outcome variable. We will work with a trained biostatistician to perform statistical analyses. The threshold of statistical significance will be set at α = 0.05, and results will be presented with 95% confidence intervals.

We will begin by describing participants’ sociodemographic and clinical characteristics at baseline. We will summarize continuous variables using the mean, standard deviation, median, and interquartile range, and we will present categorical variables as absolute and relative frequencies. We will also examine the distribution of key variables and identify missing data, which we will systematically document. Additionally, we will monitor changes in the number of participants over time.

We will then conduct bivariate analyses to assess associations between the primary outcome (mean diabetes distress score) and a range of independent variables (sociodemographic, clinical, and other relevant characteristics). We will use linear regression models to identify participant characteristics that are significantly associated with diabetes distress.

Finally, we will use multivariate models to evaluate the effect of the CommuniT1D on our outcomes of interest while controlling for potential confounding factors. Since we will collect longitudinal data (i.e., at least 2 observations per participant), we will use mixed-effects models to account for intra-individual correlation. We will analyze changes in continuous outcomes scores using linear mixed-effects models with random effects. We will estimate mean differences between baseline (pre-intervention) and follow-up while adjusting for relevant covariates. If we observe a highly skewed distribution, we will consider a log transformation or mixed quantile regression. If we dichotomize outcomes (e.g., score ≥ 1 vs. 0), we will use generalized linear mixed models (GLMMs) with a logistic link. For count outcomes (e.g., episodes of severe hypoglycemia), we will use Poisson mixed-effects models.

We will apply a stepwise modeling strategy across three levels to reduce the risk of overfitting. First, we will construct a baseline model including only the time variable (pre- vs post-intervention). Next, we will expand this into a sociodemographic model by adding age, gender, language, province of residence, and education level. Finally, we will develop a comprehensive model by incorporating other variables such as diabetes duration, type of treatment (for example, automated insulin delivery versus manual insulin pump versus multiple daily injections), comorbidities (for example, celiac disease, kidney disease), participant role (person with diabetes and/or caregiver), and perceived social support.

#### Qualitative Analyses

We will analyze interview data using MAXQDA software to conduct thematic analysis, a structured method for organizing data and analyzing themes. We will follow an established, 6-step approach. [75] We will use a combination of deductive analysis, in which we will assign themes within our initial program theory, and inductive analysis, in which we will refine our program theory by generating new themes within the Context-Mechanism-Outcome framework from what CommuniT1D members tell us. All interviews will be analyzed by two independent analysts: the study coordinator and a trained graduate student. Analyses will be reviewed by team members with expertise in realist evaluation and qualitative analyses in mental health and diabetes. We will member-check findings with interviewees and discuss and interpret preliminary findings at meetings of the steering committee, full team, and international advisory committee.

We will analyze the second set of interview data the same way as the first set, with the added analytic goal of determining how and whether the themes we identified in the first set of interviews apply to the data we collected in the second set of interviews or not.

In both instances of qualitative analysis, we will perform member-checking by returning individualized summaries to each interviewee and asking them to comment on whether we may have misinterpreted or missed anything in their interview. We will update our findings in response to nuances that they may add at this point. We will then discuss and interpret findings with meetings of the steering committee, full team, international panel, and CommuniT1D peer leaders and members who agreed to help co-develop the interview guide.

#### Triangulate qualitative and quantitative data to refine program theory

Once our qualitative and quantitative analyses are complete, we will triangulate these data into a refined Context-Mechanism-Outcome program theory evaluating how the CommuniT1D program works, for whom, and in what circumstances. This process will be collaborative and will take place over multiple meetings of the steering committee and full team in the final year of the study, with oversight by a team member with particular expertise in triangulation, particularly in patient-partnered research. [76] We will present preliminary findings at multiple CommuniT1D webinars in English and French and at an international advisory committee meeting to seek comments from CommuniT1D members, peer leaders, and international experts before finalizing the program theory.

## DISCUSSION

At the conclusion of this research project, we hope to have reduced isolation among people living in Canada whose lives are touched by T1D and produced new knowledge about the effects of this form of peer support on people in Canada whose lives are impacted by T1D. Throughout the project, we will continue to monitor for potential challenges such as difficulties in recruiting peer leaders, obstacles related to digital literacy, and sustaining engagement in peer groups. We will be monitoring these issues and continuously seeking to address them, consistent with our participatory action research approach.

Ultimately, by determining which aspects of CommuniT1D work or don’t work, for whom, and why, we hope to establish a framework within which CommuniT1D can improve and help more people. We aim to accomplish this by growing throughout Canada, increasing sustainability of the program, and possibly expanding to other locations or other conditions, such as type 2 diabetes.

## Supporting information

Appendix 1 (English only)

Appendix 2 (English only)

## Data Availability

Manuscript describes a protocol. Data are not presented yet.

## Acknowledgments

We thank M. Souleymane Gadio, Scientific Advisor in Biostatistics at VITAM-Centre de recherche en santé durable, for his support in developing the statistical analysis plan for this research protocol, and Selma Chipenda Dansokho, retired research associate at Université Laval, for her support in developing the overall study protocol and plan.

## APPENDICES

**Appendix 1: Sign up form and baseline questionnaire**

**Appendix 2: Outcomes questionnaire**

