## Appendix 1 (English only) for "A Study Protocol for CommuniT1D : A Customized Virtual Peer Support Program For People Living With Type 1 Diabetes In Canada"

### communiT1D_signup_and_baseline_questionnaire

Start of Block: Welcome

**WELCOME** CommuniT1D is a free program designed to foster social support and friendship among people in Canada whose lives are affected by type 1 diabetes (T1D) because: they have T1D themselves, they have one or more loved ones with T1D, or all of the above CommuniT1D is a new program funded by Breakthrough T1D Canada (formerly JDRF Canada) and the Canadian Institutes of Health Research. The team of experts creating the program is led by people who themselves have T1D or who are the parent of a child with T1D. Read about our team here. CommuniT1D will match small groups of adults: whose lives are affected by T1D, who want to connect with others in similar situations, and who are interested in participating in virtual meet-ups (like a Zoom meeting) for about an hour about once a month. We want to match up small groups of people who can meet at the same time, speak the same language, have things in common, and will get along well. **Interested in trying it out? Help us find the right group for you!** We will ask you to answer some questions about yourself and how T1D affects you and/or those around you. Then, someone from CommuniT1D will contact you to offer you one or more groups you can join. It's always up to you whether you stay in a group or not. You can leave a group at any time and join one or more other groups. Our team will help find the right group so that you can get the support and friendship that is right for you.

---

Important: This is not a test! If you don't know the answers, please don't worry. Feel free to choose, "I don't know," or, "I prefer not to answer," for any of the questions. **But please answer carefully! Because of the way the survey software is designed, some pages do not allow you to go back and change your previous answers.** If you want to change your answers, you may need to close the survey and to start over.

End of Block: Welcome

Start of Block: About you and T1D

**About you and T1D!** Getting to know you a bit better will help us find the right group(s) for you. Answering these questions will take about 10-20 minutes. While you are answering, you can go back to change your earlier answers, but once you submit your answers, you can't change them anymore. If you close the window, you will need to start over from the beginning. **We will**

**never share any of your information without your consent.** Your answers will only be seen by one or a few members of the research team, all of whom have strict research ethics training and certification and have signed confidentiality forms. Later, you will have the option to allow some of your answers to be used anonymously for research if you want.

---

About T1D: T1D is an autoimmune type of diabetes that happens when the immune system attacks the cells in the pancreas that make insulin. About 5-10% of people with diabetes have T1D. T1D often presents in childhood but can develop at any age. About CommuniT1D: CommuniT1D welcomes people whose lives are affected by T1D or other rare, insulin-requiring types of diabetes that are typically grouped with T1D, including Latent Autoimmune Diabetes of Adults (LADA), some forms of Maturity-Onset Diabetes of the Young (MODY), and type 3c, which is a type of diabetes that occurs due to damage to the pancreas or having had the pancreas removed. People with these types of diabetes often start using insulin right away after diagnosis, or within 3 years of diagnosis at most. We are currently unable to offer groups for people with type 2 diabetes or gestational diabetes. If you are unsure if your type of diabetes fits, you can contact:.

---

What is your relationship to diabetes? (check all that apply)

- ☐ I have T1D (or another rare type of diabetes like LADA, MODY, or type 3c)  (1)
- ☐ I have a loved one with T1D (or another rare type of diabetes like LADA, MODY, or type 3c)  (4)
- ☐ I have a more common type of diabetes (type 2 diabetes, gestational diabetes) (5)
- ☐ I have a loved one with a more common type of diabetes (type 2 diabetes, gestational diabetes)  (6)
- ☐ I have diabetes but I don't know what type of diabetes  (7)
- ☐ I have a loved one with diabetes but I don't know what type of diabetes  (8)
- ☐ I have no relationship to diabetes   (9)

*Skip To: errormessage1 If What is your relationship to diabetes? (check all that apply) = I have a more common type of diabetes (type 2 diabetes, gestational diabetes)*

*Skip To: errormessage1 If What is your relationship to diabetes? (check all that apply) = I have a loved one with a more common type of diabetes (type 2 diabetes, gestational diabetes)*

*Skip To: errormessage1 If What is your relationship to diabetes? (check all that apply) = I have a loved one with diabetes but I don't know what type of diabetes*

*Skip To: errormessage1 If What is your relationship to diabetes? (check all that apply) = I have a loved one with diabetes but I don't know what type of diabetes*

*Skip To: errormessage1 If What is your relationship to diabetes? (check all that apply) = I have no relationship to diabetes*

---

*Display this question:*

*If What is your relationship to diabetes? (check all that apply) != I have T1D (or another rare type of diabetes like LADA, MODY, or type 3c)*

*And What is your relationship to diabetes? (check all that apply) != I have a loved one with T1D (or another rare type of diabetes like LADA, MODY, or type 3c)*

We are sorry, but CommuniT1D is currently only able to support people with T1D or related types of diabetes. If you are seeking support for type 2 diabetes (the most common type of diabetes), here is a list of resources that may interest you: Diabetes Action Canada Diabetes Canada Diabète Québec National Indigenous Diabetes Association

Small Steps for Big Changes   If you are unsure if your type of diabetes fits, you can contact:.    We are wishing you a wonderful day.

*Skip To: End of Survey If  We are sorry, but CommuniT1D is currently only able to support people with T1D or related types o... Displayed*

**End of Block: About you and T1D**

---

**Start of Block: Languages**

CommuniT1D groups will hold discussions in different languages, depending what language(s) are spoken by people participating.   Out of the following list of languages spoken in Canada, which language(s) would be possible and preferable for you? (check all that apply)

| | I **would prefer to participate** in discussions in this language if possible (1) | I **could participate** in discussions in this language **but it is not my preference** (2) |
|---|---|---|
| English (1) | ☐ | ☐ |
| French (4) | ☐ | ☐ |
| Arabic (5) | ☐ | ☐ |
| Cantonese (6) | ☐ | ☐ |
| Cree (7) | ☐ | ☐ |
| German (8) | ☐ | ☐ |
| Hindi (9) | ☐ | ☐ |
| Italian (10) | ☐ | ☐ |
| Inuktitut (11) | ☐ | ☐ |
| Mandarin (12) | ☐ | ☐ |
| Mi'kmaq (13) | ☐ | ☐ |
| Ojibway (14) | ☐ | ☐ |

| | | |
|---|---|---|
| Oji-Cree (15) | ☐ | ☐ |
| Persian (Farsi) (16) | ☐ | ☐ |
| Punjabi (17) | ☐ | ☐ |
| Spanish (18) | ☐ | ☐ |
| Tagalog (19) | ☐ | ☐ |
| Urdu (20) | ☐ | ☐ |
| Other (22) | ☐ | ☐ |
| Other (24) | ☐ | ☐ |
| Other (26) | ☐ | ☐ |

End of Block: Languages

---

Start of Block: Your needs

*

Do you require any assistive services to be able to participate in a video meeting? (check all that apply)

- ☐ Automatic captioning (1)
- ☐ Screen reading (4)
- ☐ Other (5) ________________________________________________
- ☐ None of the above (6)

End of Block: Your needs

---

Start of Block: Your diabetes type

Do you have (choose one)

- ○ T1D (1)
- ○ LADA (2)
- ○ MODY (3)
- ○ Type 3c (4)
- ○ Another rare, insulin-requiring type of diabetes (5)
- ○ I prefer not to answer (6)

Page Break

End of Block: Your diabetes type

Start of Block: Your diabetes

X→

About how long has it been since you were diagnosed with ${e://Field/dtype}?

- less than 1 month (5)
- 1 to 5 months (19)
- 6 to 11 months (20)
- 1 to 2 years (21)
- 3 to 5 years (22)
- 6 to 10 years (23)
- 11 to 20 years (24)
- 21 to 30 years (25)
- 31 to 40 years (26)
- 41 to 50 years (27)
- 51 to 60 years (28)
- 61 to 70 years (29)
- 71 to 80 years (30)
- 81 years or more (31)
- I prefer not to answer (32)

*Display this question:*

*If Do you have (choose one) != T1D*

In the rest of the survey, we will refer to “T1D” to mean all rarer types of diabetes, including ${e://Field/dtype}

How do you take insulin? (check all that apply)

- ☐ A pump (Minimed, Omnipod, Tandem, Ypsomed, etc.)  (1)
- ☐ Needles/syringes  (2)
- ☐ Pens  (3)
- ☐ Other  (4) ________________________________________________
- ☐ I prefer not to answer  (5)

How do you check your blood sugar? (check all that apply)

- ☐ A continuous or flash glucose monitor (Libre, Dexcom)  (1)
- ☐ A meter (glucometer, finger pricking)  (2)
- ☐ Other  (3) ________________________________________________
- ☐ I prefer not to answer  (4)

Are you using an automated ("closed loop") insulin delivery system? This means a system that combines an insulin pump of some kind and a continuous glucose monitor of some kind so that the pump can automatically give you insulin based on your blood sugar. (check all that apply)

- ▢ Yes, I'm using a commercial system (Minimed 780G, Omnipod 5, Tandem Control IQ, YpsoPump & CamAPS, etc.) (1)
- ▢ Yes, I'm using a do-it-yourself system (AAPS, iAPS, Loop, OpenAPS, Trio, etc.) (14)
- ▢ Yes, I think so, but I don't know what kind (15)
- ▢ No, and I'm definitely not interested in them (16)
- ▢ No, but I'm interested in them or I might be interested in them (17)
- ▢ I don't know or I'm not sure (18)
- ▢ Other (12) ________________________________________________
- ▢ I prefer not to answer (13)

---

Do you currently use any adjuvant diabetes medicines? Adjuvant medicines are medicines that may have been developed for people with type 2 diabetes or other metabolic conditions but can also sometimes be recommended or prescribed to people with type 1 diabetes. (check all that apply)

- ☐ Blood pressure medicines (1)
- ☐ GLP-1 agonists (for example, Byetta, Mounjaro, Ozempic, Rybelsus, Trulicity, Victoza, Wegovy, Zepbound ) (2)
- ☐ Metformin (3)
- ☐ SGLT2 inhibitors (for example, Forxiga, Invokana, Jardiance) (4)
- ☐ Statins or other lipid-lowering drugs (5)
- ☐ None of the above (6)
- ☐ Other (7) ________________________________________________
- ☐ I don't know or I'm not sure (8)
- ☐ I prefer not to answer (9)

Do you currently follow a specific eating style? (check all that apply)

- [ ] Carnivore (1)
- [ ] Gluten-free (19)
- [ ] High fat (20)
- [ ] High fibre (21)
- [ ] High protein (22)
- [ ] Intermittent fasting (23)
- [ ] Intuitive eating (24)
- [ ] Low carb (25)
- [ ] Low fat (26)
- [ ] Low FODMAP (27)
- [ ] Keto (28)
- [ ] Paleo (29)
- [ ] Plant-based (30)
- [ ] Plant-forward (31)
- [ ] Vegetarian or vegan (32)
- [ ] Whole foods (33)

☐ None of the above (34)

☐ Other (39) ________________________________________________

☐ I prefer not to answer (41)

End of Block: Your diabetes

---

Start of Block: Your loved one's diabetes

How many loved ones do you have with T1D, LADA, MODY, type 3c, or another rare type of diabetes? (choose one)

○ 1 (1)

○ 2 (2)

○ 3 (3)

○ More than 3 (4)

○ I prefer not to answer (5)

- - - - - - - - - - - - - - - - - - - - - - - - - - - - - - - - - - - - - - - - - - - - - - - - - - - - - - - - - - - - - - - -

*Display this question:*

*If How many loved ones do you have with T1D, LADA, MODY, type 3c, or another rare type of diabetes?... = 2*

*Or How many loved ones do you have with T1D, LADA, MODY, type 3c, or another rare type of diabetes?... = 3*

Next, we will ask you the same 3 questions about each of your loved ones. Please focus on one loved one at a time, in whatever order you prefer. For example, if you have two children with T1D, you can answer for your older child first, then your younger child.

- - - - - - - - - - - - - - - - - - - - - - - - - - - - - - - - - - - - - - - - - - - - - - - - - - - - - - - - - - - - - - - -

*Display this question:*

*If How many loved ones do you have with T1D, LADA, MODY, type 3c, or another rare type of diabetes?... = More than 3*

You indicated that you have more than 3 loved ones with T1D or a similar type of diabetes. We will ask you questions about only 3 of your loved ones. Please answer for the 3 loved ones whose diabetes most impacts your life on a day-to-day basis.

**End of Block: Your loved one's diabetes**

---

**Start of Block: Your loved one's diabetes part 2**

Does your loved one have (choose one)

- ○ T1D (1)
- ○ LADA (2)
- ○ MODY (3)
- ○ Type 3c (4)
- ○ Another rare, insulin-requiring type of diabetes (5)
- ○ I prefer not to answer (6)

---

Page Break

End of Block: Your loved one’s diabetes part 2

Start of Block: Your loved one’s diabetes part 3

About how long has it been since your loved one was diagnosed with ${e://Field/dtypelovedone1}?

- ◯ less than 1 month (116)
- ◯ 1 to 5 months (117)
- ◯ 6 to 11 months (118)
- ◯ 1 to 2 years (119)
- ◯ 3 to 5 years (120)
- ◯ 6 to 10 years (121)
- ◯ 11 to 20 years (122)
- ◯ 21 to 30 years (123)
- ◯ 31 to 40 years (124)
- ◯ 41 to 50 years (125)
- ◯ 51 to 60 years (126)
- ◯ 61 to 70 years (127)
- ◯ 71 to 80 years (128)
- ◯ 81 years or more (129)
- ◯ I prefer not to answer (130)

*Display this question:*

*If Does your loved one have (choose one)  = LADA*

*Or Does your loved one have (choose one)  = MODY*

*Or Does your loved one have (choose one)  = Type 3c*

*Or Does your loved one have (choose one)  = Another rare, insulin-requiring type of diabetes*

In the rest of the survey, we will refer to “T1D” to mean all rarer types of diabetes, including ${e://Field/dtypelovedone1}

JS

How old is your loved one now?

- ○ 0 (102)
- ○ 1 (626)
- ○ 2 (627)
- ○ 3 (628)
- ○ 4 (629)
- ○ 5 (630)
- ○ 6 (631)
- ○ 7 (632)
- ○ 8 (633)
- ○ 9 (634)
- ○ 10 (635)
- ○ 11 (636)
- ○ 12 (637)
- ○ 13 (638)
- ○ 14 (639)
- ○ 15 (640)
- ○ 16 (641)
- ○ 17 (642)
- ○ 18 (643)
- ○ 19 (644)
- ○ 20 (645)

- 21 (646)
- 22 (647)
- 23 (648)
- 24 (649)
- 25 (650)
- 26 (651)
- 27 (652)
- 28 (653)
- 29 (654)
- 30 (655)
- 31 (656)
- 32 (657)
- 33 (658)
- 34 (659)
- 35 (660)
- 36 (661)
- 37 (662)
- 38 (663)
- 39 (664)
- 40 (665)
- 41 (666)

- ○ 42 (667)
- ○ 43 (668)
- ○ 44 (669)
- ○ 45 (670)
- ○ 46 (671)
- ○ 47 (672)
- ○ 48 (673)
- ○ 49 (674)
- ○ 50 (675)
- ○ 51 (676)
- ○ 52 (677)
- ○ 53 (678)
- ○ 54 (679)
- ○ 55 (680)
- ○ 56 (681)
- ○ 57 (682)
- ○ 58 (683)
- ○ 59 (684)
- ○ 60 (685)
- ○ 61 (686)
- ○ 62 (687)

- ○ 63 (688)
- ○ 64 (689)
- ○ 65 (690)
- ○ 66 (691)
- ○ 67 (692)
- ○ 68 (693)
- ○ 69 (694)
- ○ 70 (695)
- ○ 71 (696)
- ○ 72 (697)
- ○ 73 (698)
- ○ 74 (699)
- ○ 75 (700)
- ○ 76 (701)
- ○ 77 (702)
- ○ 78 (703)
- ○ 79 (704)
- ○ 80 (705)
- ○ 81 (706)
- ○ 82 (707)
- ○ 83 (708)

○ 84 (709)

○ 85 (710)

○ 86 (711)

○ 87 (712)

○ 88 (713)

○ 89 (714)

○ 90 (715)

○ 91 (716)

○ 92 (717)

○ 93 (718)

○ 94 (719)

○ 95 (720)

○ 96 (721)

○ 97 (722)

○ 98 (723)

○ 99 (724)

○ 100 or older (624)

○ I prefer not to answer (625)

**End of Block: Your loved one’s diabetes part 3**

---

**Start of Block: Your second loved one’s diabetes part 2**

Does your second loved one have (choose one)

- ○ T1D (1)
- ○ LADA (2)
- ○ MODY (3)
- ○ Type 3c (4)
- ○ Another rare, insulin-requiring type of diabetes (5)
- ○ I prefer not to answer (6)

Page Break

End of Block: Your second loved one's diabetes part 2

Start of Block: Your second loved one's diabetes part 3

About how long has it been since your second loved one was diagnosed with ${e://Field/dtypelovedone2}?

- less than 1 month (116)
- 1 to 5 months (117)
- 6 to 11 months (118)
- 1 to 2 years (119)
- 3 to 5 years (120)
- 6 to 10 years (121)
- 11 to 20 years (122)
- 21 to 30 years (123)
- 31 to 40 years (124)
- 41 to 50 years (125)
- 51 to 60 years (126)
- 61 to 70 years (127)
- 71 to 80 years (128)
- 81 years or more (129)
- I prefer not to answer (130)

*Display this question:*

*If Does your second loved one have (choose one) = LADA*

*Or Does your second loved one have (choose one) = MODY*

*Or Does your second loved one have (choose one) = Type 3c*

*Or Does your second loved one have (choose one) = Another rare, insulin-requiring type of diabetes*

In the rest of the survey, we will refer to “T1D” to mean all rarer types of diabetes, including ${e://Field/dtypelovedone2}

JS

How old is your second loved one now?

- 0 (102)
- 1 (626)
- 2 (627)
- 3 (628)
- 4 (629)
- 5 (630)
- 6 (631)
- 7 (632)
- 8 (633)
- 9 (634)
- 10 (635)
- 11 (636)
- 12 (637)
- 13 (638)
- 14 (639)
- 15 (640)
- 16 (641)
- 17 (642)
- 18 (643)
- 19 (644)
- 20 (645)

- ○ 21 (646)
- ○ 22 (647)
- ○ 23 (648)
- ○ 24 (649)
- ○ 25 (650)
- ○ 26 (651)
- ○ 27 (652)
- ○ 28 (653)
- ○ 29 (654)
- ○ 30 (655)
- ○ 31 (656)
- ○ 32 (657)
- ○ 33 (658)
- ○ 34 (659)
- ○ 35 (660)
- ○ 36 (661)
- ○ 37 (662)
- ○ 38 (663)
- ○ 39 (664)
- ○ 40 (665)
- ○ 41 (666)

- 42 (667)
- 43 (668)
- 44 (669)
- 45 (670)
- 46 (671)
- 47 (672)
- 48 (673)
- 49 (674)
- 50 (675)
- 51 (676)
- 52 (677)
- 53 (678)
- 54 (679)
- 55 (680)
- 56 (681)
- 57 (682)
- 58 (683)
- 59 (684)
- 60 (685)
- 61 (686)
- 62 (687)

- ○ 63 (688)
- ○ 64 (689)
- ○ 65 (690)
- ○ 66 (691)
- ○ 67 (692)
- ○ 68 (693)
- ○ 69 (694)
- ○ 70 (695)
- ○ 71 (696)
- ○ 72 (697)
- ○ 73 (698)
- ○ 74 (699)
- ○ 75 (700)
- ○ 76 (701)
- ○ 77 (702)
- ○ 78 (703)
- ○ 79 (704)
- ○ 80 (705)
- ○ 81 (706)
- ○ 82 (707)
- ○ 83 (708)

- 84 (709)
- 85 (710)
- 86 (711)
- 87 (712)
- 88 (713)
- 89 (714)
- 90 (715)
- 91 (716)
- 92 (717)
- 93 (718)
- 94 (719)
- 95 (720)
- 96 (721)
- 97 (722)
- 98 (723)
- 99 (724)
- 100 or older (624)
- I prefer not to answer (625)

**End of Block: Your second loved one’s diabetes part 3**

---

**Start of Block: Your third loved one’s diabetes part 2**

Does your third loved one have (choose one)

- ○ T1D (1)
- ○ LADA (2)
- ○ MODY (3)
- ○ Type 3c (4)
- ○ Another rare, insulin-requiring type of diabetes (5)
- ○ I prefer not to answer (6)

**End of Block: Your third loved one's diabetes part 2**

---

**Start of Block: Your third loved one's diabetes part 3**

About how long has it been since your third loved one was diagnosed with ${e://Field/dtypelovedone3}?

- less than 1 month (116)
- 1 to 5 months (117)
- 6 to 11 months (118)
- 1 to 2 years (119)
- 3 to 5 years (120)
- 6 to 10 years (121)
- 11 to 20 years (122)
- 21 to 30 years (123)
- 31 to 40 years (124)
- 41 to 50 years (125)
- 51 to 60 years (126)
- 61 to 70 years (127)
- 71 to 80 years (128)
- 81 years or more (129)
- I prefer not to answer (130)

*Display this question:*

*If Does your third loved one have (choose one) = LADA*

*Or Does your third loved one have (choose one) = MODY*

*Or Does your third loved one have (choose one) = Type 3c*

*Or Does your third loved one have (choose one) = Another rare, insulin-requiring type of diabetes*

In the rest of the survey, we will refer to “T1D” to mean all rarer types of diabetes, including ${e://Field/dtypelovedone3}

JS

How old is your third loved one now?

- 0 (102)
- 1 (626)
- 2 (627)
- 3 (628)
- 4 (629)
- 5 (630)
- 6 (631)
- 7 (632)
- 8 (633)
- 9 (634)
- 10 (635)
- 11 (636)
- 12 (637)
- 13 (638)
- 14 (639)
- 15 (640)
- 16 (641)
- 17 (642)
- 18 (643)
- 19 (644)
- 20 (645)

- 21 (646)
- 22 (647)
- 23 (648)
- 24 (649)
- 25 (650)
- 26 (651)
- 27 (652)
- 28 (653)
- 29 (654)
- 30 (655)
- 31 (656)
- 32 (657)
- 33 (658)
- 34 (659)
- 35 (660)
- 36 (661)
- 37 (662)
- 38 (663)
- 39 (664)
- 40 (665)
- 41 (666)

- ○ 42 (667)
- ○ 43 (668)
- ○ 44 (669)
- ○ 45 (670)
- ○ 46 (671)
- ○ 47 (672)
- ○ 48 (673)
- ○ 49 (674)
- ○ 50 (675)
- ○ 51 (676)
- ○ 52 (677)
- ○ 53 (678)
- ○ 54 (679)
- ○ 55 (680)
- ○ 56 (681)
- ○ 57 (682)
- ○ 58 (683)
- ○ 59 (684)
- ○ 60 (685)
- ○ 61 (686)
- ○ 62 (687)

- ○ 63 (688)
- ○ 64 (689)
- ○ 65 (690)
- ○ 66 (691)
- ○ 67 (692)
- ○ 68 (693)
- ○ 69 (694)
- ○ 70 (695)
- ○ 71 (696)
- ○ 72 (697)
- ○ 73 (698)
- ○ 74 (699)
- ○ 75 (700)
- ○ 76 (701)
- ○ 77 (702)
- ○ 78 (703)
- ○ 79 (704)
- ○ 80 (705)
- ○ 81 (706)
- ○ 82 (707)
- ○ 83 (708)

○ 84 (709)

○ 85 (710)

○ 86 (711)

○ 87 (712)

○ 88 (713)

○ 89 (714)

○ 90 (715)

○ 91 (716)

○ 92 (717)

○ 93 (718)

○ 94 (719)

○ 95 (720)

○ 96 (721)

○ 97 (722)

○ 98 (723)

○ 99 (724)

○ 100 or older (624)

○ I prefer not to answer (625)

**End of Block: Your third loved one’s diabetes part 3**

---

**Start of Block: Your loved one’s diabetes part 4**

*Display this question:*

*If What is your relationship to diabetes? (check all that apply) = I have a loved one with T1D (or another rare type of diabetes like LADA, MODY, or type 3c)*

*

What is your relationship to your loved one(s) with T1D or similar type of diabetes? (check all that apply)

- ▢ I am a babysitter or daycare provider of a child under age 18 with T1D (1)
- ▢ I am a child (e.g., son, daughter) of a person with T1D (2)
- ▢ I am a close friend of a person with T1D (3)
- ▢ I am a grandchild (e.g., grandson, granddaughter) of a person with T1D (4)
- ▢ I am a grandparent (e.g., grandmother, grandfather) of a child under age 18 with T1D (5)
- ▢ I am a grandparent (e.g., grandmother, grandfather) of a person 18 or older with T1D (6)
- ▢ I am a parent or guardian (e.g., mother, father, stepparent, foster parent) of a child under age 18 with T1D (7)
- ▢ I am a parent (e.g., mother, father, stepparent) of a person 18 or older with T1D (8)
- ▢ I am a partner (e.g., spouse, husband, wife, boyfriend, girlfriend) of a person with T1D (9)
- ▢ I am a sibling (e.g., brother, sister) of a person with T1D (10)
- ▢ Other (11) ________________________________________________
- ▢ I prefer not to answer (12)

*Display this question:*

*If What is your relationship to diabetes? (check all that apply) = I have a loved one with T1D (or another rare type of diabetes like LADA, MODY, or type 3c)*

JS

Have you ever done any of the following things to help your loved one manage their diabetes? (check all that apply)

- ▢ Brought them something when their blood sugar is low (1)
- ▢ Called 911 for them (2)
- ▢ Changed plans to accommodate diabetes-related needs (3)
- ▢ Changed a pump site for them or helped them change a pump site (4)
- ▢ Checked their blood sugar for them or help them check their blood sugar (5)
- ▢ Checked their blood sugar for them while they were sleeping (6)
- ▢ Counted carbs for them or helped them count carbs (7)
- ▢ Figured out their insulin doses or helped them figure out their doses (8)
- ▢ Filled out and/or submitted diabetes-related paperwork (e.g., private or government insurance paperwork) or helped them fill out and/or submit paperwork (9)
- ▢ Gave them insulin (injected insulin, used their insulin pump to give them insulin) (10)
- ▢ Helped them build a do-it-yourself system of some kind or built a system for them (e.g., xDrip, Nightscout site, AAPS, Loop, iAPS, etc.) (11)
- ▢ Helped them deal with high blood sugar while awake (12)
- ▢ Helped them deal with low blood sugar while awake (13)
- ▢ Helped them deal with high blood sugar while sleeping (14)
- ▢ Helped them deal with low blood sugar while sleeping (15)

- [ ] Inserted a continuous glucose monitoring (CGM) sensor or helped them insert a sensor (16)
- [ ] Made them meals or snacks (17)
- [ ] Ordered or picked up medications or supplies (at the pharmacy or from a supplier like Diabetes Express) (18)
- [ ] Paid for diabetes medications or supplies in whole or in part (19)
- [ ] Reminded them about diabetes-related tasks or goals (20)
- [ ] Supported them emotionally about their diabetes (21)
- [ ] Took them to medical appointments (22)
- [ ] Went to their school, work, or other location to give them insulin, bring them something they needed to manage diabetes (e.g., a blood test meter or pump supplies), or for another diabetes-related reason (23)
- [ ] Other (24) ________________________________________________
- [ ] Other (25) ________________________________________________
- [ ] Other (26) ________________________________________________

Page Break

*Display this question:*

*If What is your relationship to diabetes? (check all that apply) = I have a loved one with T1D (or another rare type of diabetes like LADA, MODY, or type 3c)*

How does (do) your loved one(s) take insulin? (check all that apply)

- ▢ A pump (Minimed, Omnipod, Tandem, Ypsomed, etc.) (1)
- ▢ Needles/syringes (2)
- ▢ Pens (3)
- ▢ Other (4) ________________________________________________
- ▢ I don't know (5)
- ▢ I prefer not to answer (6)

---

*Display this question:*

*If What is your relationship to diabetes? (check all that apply) = I have a loved one with T1D (or another rare type of diabetes like LADA, MODY, or type 3c)*

How does (do) your loved one(s) check their blood sugar? (check all that apply)

- ▢ A continuous or flash glucose monitor (Libre, Dexcom) (1)
- ▢ A meter (glucometer, finger pricking) (2)
- ▢ Other (3) ________________________________________________
- ▢ I don't know (4)
- ▢ I prefer not to answer (5)

---

*Display this question:*

*If What is your relationship to diabetes? (check all that apply) = I have a loved one with T1D (or another rare type of diabetes like LADA, MODY, or type 3c)*

Is (are) your loved one(s) using an automated ("closed loop") insulin delivery system? This means a system that combines an insulin pump of some kind and a continuous glucose monitor of some kind so that the pump can automatically give you insulin based on your blood sugar. (check all that apply)

- ☐ Yes, they're using a commercial system (Minimed 780G, Tandem Control IQ, etc.)  (1)
- ☐ Yes, they're using a do-it-yourself system (AAPS, iAPS, Loop, OpenAPS, etc.)  (2)
- ☐ Yes, I think so, but I don't know what kind  (3)
- ☐ No, and they're/we're definitely not interested in them  (4)
- ☐ No, but they're/we're interested in them or they/we might be interested in them  (5)
- ☐ I don't know or I'm not sure  (6)
- ☐ Other  (7) ________________________________________________
- ☐ I prefer not to answer  (10)

---

*Display this question:*

*If What is your relationship to diabetes? (check all that apply) = I have a loved one with T1D (or another rare type of diabetes like LADA, MODY, or type 3c)*

Does (do) your loved one(s) currently use any adjuvant diabetes medicines? Adjuvant medicines are medicines that may have been developed for people with type 2 diabetes or other metabolic conditions but can also sometimes be recommended or prescribed to people with type 1 diabetes. (check all that apply)

- ☐ Blood pressure medicines (1)
- ☐ GLP-1 agonists (for example, Byetta, Mounjaro, Ozempic, Rybelsus, Trulicity, Victoza) (2)
- ☐ Metformin (3)
- ☐ SGLT2 inhibitors (for example, Forxiga, Invokana, Jardiance) (4)
- ☐ Statins or other lipid-lowering drugs (5)
- ☐ None of the above (6)
- ☐ Other (7) ________________________________________________
- ☐ I don't know or I'm not sure (8)
- ☐ I prefer not to answer (9)

---

*Display this question:*

*If What is your relationship to diabetes? (check all that apply) = I have a loved one with T1D (or another rare type of diabetes like LADA, MODY, or type 3c)*

Does (do) your loved one(s) currently follow a specific eating style? (check all that apply)

- ☐ Carnivore (20)
- ☐ Gluten-free (40)
- ☐ High fat (41)
- ☐ High fibre (42)
- ☐ High protein (43)
- ☐ Intermittent fasting (44)
- ☐ Intuitive eating (45)
- ☐ Low carb (46)
- ☐ Low fat (47)
- ☐ Low FODMAP (48)
- ☐ Keto (49)
- ☐ Paleo (50)
- ☐ Plant-based (51)
- ☐ Plant-forward (52)
- ☐ Vegetarian or vegan (53)
- ☐ Whole foods (54)

☐ None of the above (55)

☐ Other (56) ________________________________________________

☐ I don’t know (57)

☐ I prefer not to answer (58)

End of Block: Your loved one’s diabetes part 4

---

Start of Block: Contacting you

*

Please provide your email address and a phone number as a backup. We will primarily contact you by email, but if we are unable to reach you by email, we will use your phone number.

○ Email: (6) ________________________________________________

○ Email (repeat): (7) ________________________________________________

○ Phone number: (8) ________________________________________________

○ Phone number (repeat): (9) ________________________________________________

○ Comments (optional; for example, let us know here if you’d like to be in a group with a specific person; if no comments enter "NONE"): (10) ________________________________________________

End of Block: Contacting you

---

Start of Block: About you

How old are you (in years)?

- ○ 18 (1)
- ○ 19 (2)
- ○ 20 (3)
- ○ 21 (4)
- ○ 22 (5)
- ○ 23 (6)
- ○ 24 (7)
- ○ 25 (8)
- ○ 26 (9)
- ○ 27 (10)
- ○ 28 (11)
- ○ 29 (12)
- ○ 30 (13)
- ○ 31 (14)
- ○ 32 (15)
- ○ 33 (16)
- ○ 34 (17)
- ○ 35 (18)
- ○ 36 (19)
- ○ 37 (20)
- ○ 38 (21)

- 39 (22)
- 40 (23)
- 41 (24)
- 42 (25)
- 43 (26)
- 44 (27)
- 45 (28)
- 46 (29)
- 47 (30)
- 48 (31)
- 49 (32)
- 50 (33)
- 51 (34)
- 52 (35)
- 53 (36)
- 54 (37)
- 55 (38)
- 56 (39)
- 57 (40)
- 58 (41)
- 59 (42)

- 60 (43)
- 61 (44)
- 62 (45)
- 63 (46)
- 64 (47)
- 65 (48)
- 66 (49)
- 67 (50)
- 68 (51)
- 69 (52)
- 70 (53)
- 71 (54)
- 72 (55)
- 73 (56)
- 74 (57)
- 75 (58)
- 76 (59)
- 77 (60)
- 78 (61)
- 79 (62)
- 80 (63)

- 81 (64)
- 82 (65)
- 83 (66)
- 84 (67)
- 85 (68)
- 86 (69)
- 87 (70)
- 88 (71)
- 89 (72)
- 90 (73)
- 91 (74)
- 92 (75)
- 93 (76)
- 94 (77)
- 95 (78)
- 96 (79)
- 97 (80)
- 98 (81)
- 99 (82)
- 100 (83)
- I prefer not to answer (84)

In which province or territory do you currently live? (choose one)

- ◯ Alberta (1)
- ◯ British Columbia (2)
- ◯ Manitoba (3)
- ◯ New Brunswick (4)
- ◯ Newfoundland/Labrador (5)
- ◯ Northwest Territories (6)
- ◯ Nova Scotia (7)
- ◯ Nunavut (8)
- ◯ Ontario (9)
- ◯ Prince Edward Island (10)
- ◯ Quebec (11)
- ◯ Saskatchewan (12)
- ◯ Yukon (13)
- ◯ I am a Canadian or permanent resident living outside Canada (14)
- ◯ I prefer not to answer (15)

How would you best describe your current employment status? (choose one)

- ○ Employed (includes part-time employment and/or self-employed) (1)
- ○ Retired (2)
- ○ Semi-retired (3)
- ○ Student (4)
- ○ Unemployed (5)
- ○ Other, please specify: (6) ________________________________________________
- ○ I prefer not to answer (7)

Which of the following best describes your racial or ethnic group(s)? (check all that apply)

- ☐ Aboriginal person from outside of Canada (e.g., Native American, Maori, Quechua) (1)
- ☐ Asian - East (e.g., Chinese, Japanese, Korean) (2)
- ☐ Asian - Central (e.g., Kazakhstani, Uzbekistani) (3)
- ☐ Asian - South (e.g., Indian, Pakistani, Sri Lankan) (4)
- ☐ Asian - South-East (e.g., Malaysian, Filipino, Vietnamese) (5)
- ☐ Black - African (e.g., Ghanaian, Kenyan, Somali) (6)
- ☐ Black - Caribbean (e.g., Barbadian, Jamaican) (7)
- ☐ Black - North American (e.g., Canadian, American) (8)
- ☐ First Nations (9)
- ☐ Inuit (10)
- ☐ Latin American (e.g., Chilean, Mexican, Salvadorian) (11)
- ☐ Metis (Metis Nation in Canada) (12)
- ☐ Middle Eastern (e.g., Egyptian, Iranian, Lebanese) (13)
- ☐ North African (e.g., Moroccan, Tunisian) (14)
- ☐ White / European (e.g., English, Italian, Portuguese, Russian) (15)
- ☐ White / North American (e.g., Canadian, American) (16)

☐ Other, please specify : (17) ________________________________________________

☐ I prefer not to answer (18)

Were you born in Canada? (choose one)

- ○ Yes (1)
- ○ No (2)
- ○ I prefer not to answer (3)

*Display this question:*

*If Were you born in Canada? (choose one) = No*

When did you arrive in Canada?

- ○ Less than 5 years ago (1)
- ○ 5-10 years ago (2)
- ○ More than 10 years ago (3)
- ○ I prefer not to answer (4)

Do you have a specific religious or spiritual practice that is important in your life? (choose one)

- ○ Yes (1)
- ○ No (2)
- ○ I prefer not to answer (3)

*Display this question:*

*If Do you have a specific religious or spiritual practice that is important in your life? (choose one) = Yes*

What is your religious or spiritual practice? (check all that apply)

- ☐ Anglican (20)
- ☐ Baptist (39)
- ☐ Buddhist (40)
- ☐ Catholic (41)
- ☐ Christian (42)
- ☐ Church of Jesus Christ of Latter-Day Saints (43)
- ☐ Hindu (44)
- ☐ Indigenous Spirituality (45)
- ☐ Islam (46)
- ☐ Jehovah's Witness (47)
- ☐ Jewish (48)
- ☐ Lutheran (49)
- ☐ Pentecostal (50)
- ☐ Presbyterian (51)
- ☐ Sikh (52)
- ☐ United Church (53)

- [ ] Other, please specify: (54) ________________________________________________
- [ ] No religious affiliation (55)
- [ ] I prefer not to answer (56)

---

What sex* were you assigned at birth, meaning what was put on your original birth certificate? (choose one)

- ○ Female (1)
- ○ Intersex (2)
- ○ Male (3)
- ○ I prefer not to answer (4)

---

With which gender identity* do you align? (choose one)

- ○ A man (1)
- ○ A woman (2)
- ○ Gender-fluid (3)
- ○ Indigenous or other cultural gender minority identity (e.g., two-spirit) (4)
- ○ Non-binary (5)
- ○ Other (6) ________________________________________________
- ○ I prefer not to answer (7)

---

*For more information, see “What is gender? What is sex?” on the Canadian Institutes of Health Research website: https://cihr-irsc.gc.ca/e/48642.html

Do you consider yourself a member of the 2SLGBTQIA+ community? (choose one)

○ Yes (1)

○ No (2)

○ I don’t know (3)

○ I prefer not to answer (4)

Page Break

*Display this question:*

*If What is your relationship to diabetes? (check all that apply) = I have T1D (or another rare type of diabetes like LADA, MODY, or type 3c)*

Do you live with or have you had any of the following health conditions in addition to T1D? (check all that apply)

- ▢ Anxiety (1)
- ▢ Attention deficit disorder (ADD) or attention deficit hyperactive disorder (ADHD) (34)
- ▢ Autism (35)
- ▢ Bipolar disorder (36)
- ▢ Cancer (37)
- ▢ Celiac disease (38)
- ▢ Depression (39)
- ▢ Diabetic retinopathy or other eye problems (e.g., glaucoma, blindness) (40)
- ▢ Disordered eating (including diabulimia, orthorexia, and other forms of disordered eating) (41)
- ▢ Food allergies (42)
- ▢ Food sensitivities (43)
- ▢ Frozen shoulder (44)
- ▢ Gastroparesis or slow gastric emptying (45)
- ▢ Gluten sensitivity (46)
- ▢ Heart disease or other heart problems (47)

- [ ] Hypo unawareness (not feeling low blood sugars) (48)
- [ ] Insulin allergies or insulin hypersensitivity (49)
- [ ] Insulin resistance (50)
- [ ] Kidney problems (51)
- [ ] Learning disabilities (52)
- [ ] Nerve problems (lack of sensation in the feet, slow healing, other nerve-related problems) (53)
- [ ] Obesity (54)
- [ ] Polycystic ovarian syndrome (PCOS) (55)
- [ ] Raynaud’s syndrome or phenomenon (56)
- [ ] Rheumatoid arthritis (57)
- [ ] Sexual dysfunction (58)
- [ ] Skin sensitivities, allergies, or other dermatological conditions (including allergies to adhesives) (59)
- [ ] Thyroid conditions (hypo or hyper, Hashimoto’s, Graves’) (60)
- [ ] Transplant (kidney, islet cells, pancreas, other organ) (61)
- [ ] Other autoimmune conditions (62) ________________________________________________

☐ Other mental health conditions (63) ________________________________________________

☐ Other (64) ________________________________________________

☐ I don’t know (65)

☐ I prefer not to answer (66)

Page Break

*Display this question:*

*If What is your relationship to diabetes? (check all that apply) = I have a loved one with T1D (or another rare type of diabetes like LADA, MODY, or type 3c)*

Does (do) your loved one(s) live with or have they had any of the following health conditions in addition to T1D? (check all that apply)

- ☐ Anxiety (33)
- ☐ Attention deficit disorder (ADD) or attention deficit hyperactive disorder (ADHD) (65)
- ☐ Autism (66)
- ☐ Bipolar disorder (67)
- ☐ Cancer (68)
- ☐ Celiac disease (69)
- ☐ Depression (70)
- ☐ Diabetic retinopathy or other eye problems (e.g., glaucoma, blindness) (71)
- ☐ Disordered eating (including diabulimia, orthorexia, and other forms of disordered eating) (72)
- ☐ Food allergies (73)
- ☐ Food sensitivities (74)
- ☐ Frozen shoulder (75)
- ☐ Gastroparesis or slow gastric emptying (76)
- ☐ Gluten sensitivity (77)
- ☐ Heart disease or other heart problems (78)

- [ ] Hypo unawareness (not feeling low blood sugars)  (79)
- [ ] Insulin allergies or insulin hypersensitivity  (80)
- [ ] Insulin resistance  (81)
- [ ] Kidney problems  (82)
- [ ] Learning disabilities  (83)
- [ ] Nerve problems (lack of sensation in the feet, slow healing, other nerve-related problems)  (84)
- [ ] Obesity  (85)
- [ ] Raynaud’s syndrome or phenomenon  (86)
- [ ] Rheumatoid arthritis  (87)
- [ ] Skin sensitivities, allergies, or other dermatological conditions (including allergies to adhesives)  (88)
- [ ] Thyroid conditions (hypo or hyper, Hashimoto’s, Graves’)  (89)
- [ ] Transplant (kidney, islet cells, pancreas, other organ)  (90)
- [ ] Other autoimmune conditions  (91)
- [ ] Other mental health conditions  (92)
- [ ] Other  (93)
- [ ] I don’t know  (94)

☐ I prefer not to answer (95)

Page Break

Below is a list of life situations and activities that may have positive or negative impacts on a person’s life, and may also be more challenging or require extra planning when T1D is in the picture. Are any of these relevant to you, either because you recently experienced this, you are currently experiencing this, or you will soon experience this? (check all that apply)

- ▢ Aging out of insurance coverage (e.g., parents' insurance coverage, provincial pump coverage that ends at a certain age) (1)
- ▢ Caregiving (in addition to T1D; e.g., caring for another person in the family who has a different condition) (2)
- ▢ Changing countries (3)
- ▢ Changing provinces (4)
- ▢ Discrimination (being discriminated against because of diabetes or because of other aspects of your identity like your gender, ethnicity, religion, etc.) (5)
- ▢ Disordered eating (eating more than you'd like, eating less than you'd like, overly strict eating, eating without taking enough insulin, etc.) (6)
- ▢ Divorce, separation, or other relationship breakdown (7)
- ▢ Exercise/physical activity (8)
- ▢ Endurance sports (running, triathlon, cycling, swimming, cross-country skiing, etc.) (9)
- ▢ Functional fitness training (CrossFit, F45, Orange Theory, etc.) (10)
- ▢ Outdoor activities (downhill skiing, hiking, snowshoeing, etc.) (11)
- ▢ Strength training (lifting weights, using other resistance tools like bands, following a bodyweight plan, etc.) (13)
- ▢ Physical labour (any job that requires lots of movement) (12)

- Team sports (school sports teams, club sports teams, recreational leagues, etc.) (14)

- Other exercise or physical activities (15) ________________________________________________

- Environmental disruption (wildfire, flood, etc.) (16)

- Family conflict (about diabetes or about other issues) (17)

- Financial difficulties or stress (due to diabetes or for other reasons) (18)

- Food insecurity (having trouble affording food, being unsure if food will be available when you need it) (19)

- Frequent travel (for work, family, or pleasure) (20)

- Grandparenting (21)

- Growth hormones (natural hormones typically experienced by children and sometimes young adults) (22)

- Health care work (e.g., dietitian, nurse, physician) (23)

- High-pressure job (24)

- Hormone blockers (taking medication to prevent hormones) (25)

- Housing difficulties (having trouble finding a place to live) (26)

- Incarceration (being in jail, being in a mental health institution) (27)

- Insurance difficulties (e.g., not having insurance coverage for diabetes-related expenses, having difficulty getting life insurance) (28)

- ☐ Living alone (29)
- ☐ Job difficulties or job-related stress (30)
- ☐ Managing multiple chronic health conditions (31)
- ☐ Menopause (32)
- ☐ Military service (33)
- ☐ New baby/child in the family (34)
- ☐ New diabetes diagnosis (35)
- ☐ New job (36)
- ☐ New romantic relationship (37)
- ☐ Parenting babies/toddlers (38)
- ☐ Parenting school-aged children (39)
- ☐ Parenting teens (40)
- ☐ Partying (with or without substance use) (41)
- ☐ Performing arts (e.g., actor, dancer, musician, theatre crew, tv or film crew) (42)
- ☐ Perimenopause (43)
- ☐ Pregnancy (44)

- ☐ Puberty (45)
- ☐ Relationships & dating (46)
- ☐ Retirement (47)
- ☐ School activities and clubs (chess club, math club, school paper, etc.) (48)
- ☐ School difficulties or stress (49)
- ☐ Sexual assault (unwanted sexual actions by others) (50)
- ☐ Shared custody (51)
- ☐ Shift work (e.g., night shift, 2-week-in-2-week-out rotation, etc.) (52)
- ☐ Single parenting (53)
- ☐ Trying to conceive (54)
- ☐ Trying to gain weight/muscle (55)
- ☐ Trying to lose weight/fat (56)
- ☐ Unemployment (57)
- ☐ Violence (in the home, at school, in other contexts) (58)
- ☐ Other (59) ________________________________________________
- ☐ I prefer not to answer (60)

Page Break

*Display this question:*

*If What is your relationship to diabetes? (check all that apply) = I have a loved one with T1D (or another rare type of diabetes like LADA, MODY, or type 3c)*

Below is a list of life situations and activities that may have positive or negative impacts on a person's life, and may also be more challenging or require extra planning when T1D is in the picture. **We previously asked these questions about you. Now we are asking about your loved one(s).** Are any of these **relevant to your loved one(s)**, either because your loved one recently experienced this, your loved one is currently experiencing this, or your loved one will soon experience this? (check all that apply)

- ▢ Aging out of insurance coverage (e.g., parents' insurance coverage, provincial pump coverage that ends at a certain age) (1)
- ▢ Caregiving (in addition to T1D; e.g., caring for another person in the family who has a different condition) (2)
- ▢ Changing countries (3)
- ▢ Changing provinces (4)
- ▢ Discrimination (being discriminated against because of diabetes or because of other aspects of your identity like your gender, ethnicity, religion, etc.) (5)
- ▢ Disordered eating (eating more than you'd like, eating less than you'd like, overly strict eating, eating without taking enough insulin, etc.) (6)
- ▢ Divorce, separation, or other relationship breakdown (7)
- ▢ Exercise/physical activity (8)
- ▢ Endurance sports (running, triathlon, cycling, swimming, cross-country skiing, etc.) (9)
- ▢ Functional fitness training (CrossFit, F45, Orange Theory, etc.) (10)
- ▢ Outdoor activities (downhill skiing, hiking, snowshoeing, etc.) (11)
- ▢ Strength training (lifting weights, using other resistance tools like bands, following a bodyweight plan, etc.) (13)
- ▢ Physical labour (any job that requires lots of movement) (12)

☐ Team sports (school sports teams, club sports teams, recreational leagues, etc.) (14)

☐ Other exercise or physical activities (15) ________________________________________________

☐ Environmental disruption (wildfire, flood, etc.) (16)

☐ Family conflict (about diabetes or about other issues) (17)

☐ Financial difficulties or stress (due to diabetes or for other reasons) (18)

☐ Food insecurity (having trouble affording food, being unsure if food will be available when you need it) (19)

☐ Frequent travel (for work, family, or pleasure) (20)

☐ Grandparenting (21)

☐ Growth hormones (natural hormones typically experienced by children and sometimes young adults) (22)

☐ Health care work (e.g., dietitian, nurse, physician) (23)

☐ High-pressure job (24)

☐ Hormone blockers (taking medication to prevent hormones) (25)

☐ Housing difficulties (having trouble finding a place to live) (26)

☐ Incarceration (being in jail, being in a mental health institution) (27)

☐ Insurance difficulties (e.g., not having insurance coverage for diabetes-related expenses, having difficulty getting life insurance) (28)

- [ ] Living alone (29)
- [ ] Job difficulties or job-related stress (30)
- [ ] Managing multiple chronic health conditions (31)
- [ ] Menopause (32)
- [ ] Military service (33)
- [ ] New baby/child in the family (34)
- [ ] New diabetes diagnosis (35)
- [ ] New job (36)
- [ ] New romantic relationship (37)
- [ ] Parenting babies/toddlers (38)
- [ ] Parenting school-aged children (39)
- [ ] Parenting teens (40)
- [ ] Partying (with or without substance use) (41)
- [ ] Performing arts (e.g., actor, dancer, musician, theatre crew, tv or film crew) (42)
- [ ] Perimenopause (43)
- [ ] Pregnancy (44)

- [ ] Puberty (45)
- [ ] Relationships & dating (46)
- [ ] Retirement (47)
- [ ] School activities and clubs (chess club, math club, school paper, etc.) (48)
- [ ] School difficulties or stress (49)
- [ ] Sexual assault (unwanted sexual actions by others) (50)
- [ ] Shared custody (51)
- [ ] Shift work (e.g., night shift, 2-week-in-2-week-out rotation, etc.) (52)
- [ ] Single parenting (53)
- [ ] Trying to conceive (54)
- [ ] Trying to gain weight/muscle (55)
- [ ] Trying to lose weight/fat (56)
- [ ] Unemployment (57)
- [ ] Violence (in the home, at school, in other contexts) (58)
- [ ] Other (59) ________________________________________________
- [ ] I prefer not to answer (60)

Page Break

Diabetes involves a lot of numbers; for example, A1c, blood glucose, time in range, and units of insulin. Even though we know the numbers can vary due to variations in labs, technology, genetics, hormones, and body chemistry, these numbers can still feel like a report card. Some people find discussions about these numbers useful while others find such discussions difficult. Each CommuniT1D group will determine how and whether they want to talk about numbers through regular anonymous polls. To help us connect you with the right group for you, **please tell us how you might feel about discussions of these numbers.** (choose one)

- ○ I would prefer to be in a group where talking about numbers (A1c, blood glucose, time in range, units of insulin, etc.) is completely fine (1)
- ○ I would prefer to be in a group where talking about numbers (A1c, blood glucose, time in range, units of insulin, etc.) is not allowed (4)
- ○ I don't have a preference (5)
- ○ I don't know (6)
- ○ I prefer not to answer (7)

**End of Block: About you**

---

**Start of Block: What matters most to you?**

The goal of CommuniT1D is to connect groups of people who have T1D and other things in common. To help us find the best fit for you, we need to know what is most important to you when connecting with others. We will ask you to: Identify up to 6 things you'd like us to consider when matching you with a group of others, then Arrange the things you've identified from most important to least important.

**End of Block: What matters most to you?**

---

**Start of Block: What matters most to you: Step 1**

*

Below are the things that you listed about yourself and/or your loved one(s) with T1D. **Please identify up to 6 things about you and/or your loved one(s) you'd like us to consider when matching you with a group of others** (check all that apply)

*Display this choice:*
*If matchvar1 Is Not Empty*

☐ ${e://Field/matchvar1} (76)

*Display this choice:*
*If matchvar2 Is Not Empty*

☐ ${e://Field/matchvar2} (77)

*Display this choice:*
*If matchvar3 Is Not Empty*

☐ ${e://Field/matchvar3} (78)

*Display this choice:*
*If matchvar4 Is Not Empty*

☐ ${e://Field/matchvar4} (79)

*Display this choice:*
*If matchvar5 Is Not Empty*

☐ ${e://Field/matchvar5} (80)

*Display this choice:*
*If matchvar6 Is Not Empty*

☐ ${e://Field/matchvar6} (81)

*Display this choice:*
*If matchvar7 Is Not Empty*

☐ ${e://Field/matchvar7} (82)

*Display this choice:*
*If matchvar46 Is Not Empty*

☐ ${e://Field/matchvar46} (83)

*Display this choice:*
*If matchvar47 Is Not Empty*

☐ ${e://Field/matchvar47} (84)

*Display this choice:*
*If matchvar48 Is Not Empty*

☐ ${e://Field/matchvar48} (85)

*Display this choice:*
*If matchvar49 Is Not Empty*

☐ ${e://Field/matchvar49} (86)

*Display this choice:*
*If matchvar50 Is Not Empty*

☐ ${e://Field/matchvar50} (87)

*Display this choice:*
*If matchvar8 Is Not Empty*

☐ ${e://Field/matchvar8} (88)

*Display this choice:*
*If matchvar9 Is Not Empty*

☐ ${e://Field/matchvar9} (89)

*Display this choice:*
*If matchvar10 Is Not Empty*

☐ ${e://Field/matchvar10} (90)

*Display this choice:*
*If matchvar11 Is Not Empty*

☐ ${e://Field/matchvar11} (91)

*Display this choice:*
*If matchvar12 Is Not Empty*

☐ ${e://Field/matchvar12} (92)

*Display this choice:*

*If matchvar13 Is Not Empty*

☐ ${e://Field/matchvar13} (93)

*Display this choice:*

*If matchvar14 Is Not Empty*

☐ ${e://Field/matchvar14} (94)

*Display this choice:*

*If matchvar15 Is Not Empty*

☐ ${e://Field/matchvar15} (95)

*Display this choice:*

*If matchvar16 Is Not Empty*

☐ ${e://Field/matchvar16} (96)

*Display this choice:*

*If matchvar17 Is Not Empty*

☐ ${e://Field/matchvar17} (97)

*Display this choice:*

*If matchvar18 Is Not Empty*

☐ ${e://Field/matchvar18} (98)

*Display this choice:*

*If matchvar19 Is Not Empty*

☐ ${e://Field/matchvar19} (99)

*Display this choice:*

*If matchvar20 Is Not Empty*

☐ ${e://Field/matchvar20} (100)

*Display this choice:*

*If matchvar21 Is Not Empty*

☐ ${e://Field/matchvar21} (101)

*Display this choice:*

*If matchvar22 Is Not Empty*

☐ ${e://Field/matchvar22} (102)

*Display this choice:*

*If matchvar23 Is Not Empty*

☐ ${e://Field/matchvar23} (103)

*Display this choice:*

*If matchvar24 Is Not Empty*

☐ ${e://Field/matchvar24} (104)

*Display this choice:*

*If matchvar25 Is Not Empty*

☐ ${e://Field/matchvar25} (105)

*Display this choice:*

*If matchvar26 Is Not Empty*

☐ ${e://Field/matchvar26} (106)

*Display this choice:*

*If matchvar27 Is Not Empty*

☐ ${e://Field/matchvar27} (107)

*Display this choice:*

*If matchvar28 Is Not Empty*

☐ ${e://Field/matchvar28} (108)

*Display this choice:*

*If matchvar29 Is Not Empty*

☐ ${e://Field/matchvar29} (109)

*Display this choice:*

*If matchvar30 Is Not Empty*

☐ ${e://Field/matchvar30} (110)

*Display this choice:*

*If matchvar31 Is Not Empty*

☐ ${e://Field/matchvar31} (111)

*Display this choice:*

*If matchvar32 Is Not Empty*

☐ ${e://Field/matchvar32} (112)

*Display this choice:*

*If matchvar33 Is Not Empty*

☐ ${e://Field/matchvar33} (113)

*Display this choice:*

*If matchvar34 Is Not Empty*

☐ ${e://Field/matchvar34} (114)

*Display this choice:*

*If matchvar35 Is Not Empty*

☐ ${e://Field/matchvar35} (115)

*Display this choice:*

*If matchvar36 Is Not Empty*

☐ ${e://Field/matchvar36} (116)

*Display this choice:*

*If matchvar37 Is Not Empty*

☐ ${e://Field/matchvar37} (122)

*Display this choice:*

*If matchvar38 Is Not Empty*

${e://Field/matchvar38} (123)

*Display this choice:*

*If matchvar39 Is Not Empty*

${e://Field/matchvar39} (124)

*Display this choice:*

*If matchvar40 Is Not Empty*

${e://Field/matchvar40} (125)

*Display this choice:*

*If matchvar41 Is Not Empty*

${e://Field/matchvar41} (126)

*Display this choice:*

*If matchvar42 Is Not Empty*

${e://Field/matchvar42} (127)

*Display this choice:*

*If matchvar43 Is Not Empty*

${e://Field/matchvar43} (128)

*Display this choice:*

*If matchvar44 Is Not Empty*

${e://Field/matchvar44} (129)

*Display this choice:*

*If matchvar45 Is Not Empty*

${e://Field/matchvar45} (130)

*Display this choice:*

*If matchvar63 Is Not Empty*

${e://Field/matchvar63} (131)

*Display this choice:*

*If matchvar63-2 Is Not Empty*

☐ ${e://Field/matchvar63-2} (132)

*Display this choice:*
*If matchvar63-3 Is Not Empty*

☐ ${e://Field/matchvar63-3} (133)

*Display this choice:*
*If matchvar51 Is Not Empty*

☐ ${e://Field/matchvar51} (134)

*Display this choice:*
*If matchvar52 Is Not Empty*

☐ ${e://Field/matchvar52} (135)

*Display this choice:*
*If matchvar53 Is Not Empty*

☐ ${e://Field/matchvar53} (136)

*Display this choice:*
*If matchvar54 Is Not Empty*

☐ ${e://Field/matchvar54} (137)

*Display this choice:*
*If matchvar55 Is Not Empty*

☐ ${e://Field/matchvar55} (138)

*Display this choice:*
*If matchvar56 Is Not Empty*

☐ ${e://Field/matchvar56} (139)

*Display this choice:*
*If matchvar57 Is Not Empty*

☐ ${e://Field/matchvar57} (140)

*Display this choice:*

*If matchvar58 Is Not Empty*

☐ ${e://Field/matchvar58} (141)

*Display this choice:*

*If matchvar59 Is Not Empty*

☐ ${e://Field/matchvar59} (142)

*Display this choice:*

*If matchvar60 Is Not Empty*

☐ ${e://Field/matchvar60} (143)

*Display this choice:*

*If matchvar61 Is Not Empty*

☐ ${e://Field/matchvar61} (144)

*Display this choice:*

*If matchvar62 Is Not Empty*

☐ ${e://Field/matchvar62} (145)

*Display this choice:*

*If matchvar64 Is Not Empty*

☐ ${e://Field/matchvar64} (146)

*Display this choice:*

*If matchvar65 Is Not Empty*

☐ ${e://Field/matchvar65} (147)

*Display this choice:*

*If matchvar66 Is Not Empty*

☐ ${e://Field/matchvar66} (148)

*Display this choice:*

*If matchvar67 Is Not Empty*

☐ ${e://Field/matchvar67} (149)

*Display this choice:*

*If matchvar68 Is Not Empty*

☐ ${e://Field/matchvar68} (150)

*Display this choice:*

*If matchvar69 Is Not Empty*

☐ ${e://Field/matchvar69} (151)

*Display this choice:*

*If matchvar70 Is Not Empty*

☐ ${e://Field/matchvar70} (152)

*Display this choice:*

*If matchvar71 Is Not Empty*

☐ ${e://Field/matchvar71} (153)

*Display this choice:*

*If matchvar72 Is Not Empty*

☐ ${e://Field/matchvar72} (154)

*Display this choice:*

*If matchvar73 Is Not Empty*

☐ ${e://Field/matchvar73} (155)

*Display this choice:*

*If matchvar74 Is Not Empty*

☐ ${e://Field/matchvar74} (156)

*Display this choice:*

*If matchvar75 Is Not Empty*

☐ ${e://Field/matchvar75} (157)

*Display this choice:*

*If matchvar76 Is Not Empty*

☐ ${e://Field/matchvar76} (158)

*Display this choice:*

*If matchvar77 Is Not Empty*

☐ ${e://Field/matchvar77} (159)

*Display this choice:*

*If matchvar78 Is Not Empty*

☐ ${e://Field/matchvar78} (160)

*Display this choice:*

*If matchvar79 Is Not Empty*

☐ ${e://Field/matchvar79} (161)

*Display this choice:*

*If matchvar80 Is Not Empty*

☐ ${e://Field/matchvar80} (162)

*Display this choice:*

*If matchvar81 Is Not Empty*

☐ ${e://Field/matchvar81} (163)

*Display this choice:*

*If matchvar82 Is Not Empty*

☐ ${e://Field/matchvar82} (164)

*Display this choice:*

*If matchvar83 Is Not Empty*

☐ ${e://Field/matchvar83} (165)

*Display this choice:*

*If matchvar84 Is Not Empty*

☐ ${e://Field/matchvar84} (166)

*Display this choice:*

*If matchvar85 Is Not Empty*

☐ ${e://Field/matchvar85} (167)

*Display this choice:*

*If matchvar86 Is Not Empty*

☐ ${e://Field/matchvar86} (168)

*Display this choice:*

*If matchvar87 Is Not Empty*

☐ ${e://Field/matchvar87} (169)

*Display this choice:*

*If matchvar88 Is Not Empty*

☐ ${e://Field/matchvar88} (170)

*Display this choice:*

*If matchvar89 Is Not Empty*

☐ ${e://Field/matchvar89} (172)

*Display this choice:*

*If matchvar90 Is Not Empty*

☐ ${e://Field/matchvar90} (173)

*Display this choice:*

*If matchvar91 Is Not Empty*

☐ ${e://Field/matchvar91} (174)

*Display this choice:*

*If matchvar92 Is Not Empty*

☐ ${e://Field/matchvar92} (176)

*Display this choice:*

*If matchvar93 Is Not Empty*

☐ ${e://Field/matchvar93} (177)

*Display this choice:*
*If matchvar94 Is Not Empty*

☐ ${e://Field/matchvar94} (178)

*Display this choice:*
*If matchvar95 Is Not Empty*

☐ ${e://Field/matchvar95} (179)

*Display this choice:*
*If matchvar96 Is Not Empty*

☐ ${e://Field/matchvar96} (180)

*Display this choice:*
*If matchvar97 Is Not Empty*

☐ ${e://Field/matchvar97} (181)

*Display this choice:*
*If matchvar98 Is Not Empty*

☐ ${e://Field/matchvar98} (182)

*Display this choice:*
*If matchvar99 Is Not Empty*

☐ ${e://Field/matchvar99} (183)

*Display this choice:*
*If matchvar100 Is Not Empty*

☐ ${e://Field/matchvar100} (184)

*Display this choice:*
*If matchvar101 Is Not Empty*

☐ ${e://Field/matchvar101} (185)

*Display this choice:*

If matchvarW Is Not Empty

☐ ${e://Field/matchvarW} (186)

Display this choice:
If matchvarW1 Is Not Empty

☐ ${e://Field/matchvarW1} (187)

Display this choice:
If matchvarW2 Is Not Empty

☐ ${e://Field/matchvarW2} (188)

Display this choice:
If matchvarEthinicText Is Not Empty

☐ ${e://Field/matchvarEthinicText} (189)

Display this choice:
If matchvar102 Is Not Empty

☐ ${e://Field/matchvar102} (190)

Display this choice:
If matchvar103 Is Not Empty

☐ ${e://Field/matchvar103} (191)

Display this choice:
If matchvaryrlessthan5 Is Not Empty

☐ ${e://Field/matchvaryrlessthan5} (192)

Display this choice:
If matchvar5-10yrsagofr Is Not Empty

☐ ${e://Field/matchvar5-10yrsago} (193)

Display this choice:
If matchvarmorethan10yrs Is Not Empty

☐ ${e://Field/matchvarmorethan10yrs} (194)

*Display this choice:*

*If matchvar104 Is Not Empty*

☐ ${e://Field/matchvar104} (195)

*Display this choice:*

*If matchvar105 Is Not Empty*

☐ ${e://Field/matchvar105} (196)

*Display this choice:*

*If matchvar106 Is Not Empty*

☐ ${e://Field/matchvar106} (197)

*Display this choice:*

*If matchvar107 Is Not Empty*

☐ ${e://Field/matchvar107} (198)

*Display this choice:*

*If matchvar108 Is Not Empty*

☐ ${e://Field/matchvar108} (199)

*Display this choice:*

*If matchvar109 Is Not Empty*

☐ ${e://Field/matchvar109} (200)

*Display this choice:*

*If matchvar110 Is Not Empty*

☐ ${e://Field/matchvar110} (201)

*Display this choice:*

*If matchvar111 Is Not Empty*

☐ ${e://Field/matchvar111} (202)

*Display this choice:*

*If matchvar112 Is Not Empty*

☐ ${e://Field/matchvar112} (203)

*Display this choice:*

*If matchvar113 Is Not Empty*

☐ ${e://Field/matchvar113} (204)

*Display this choice:*

*If matchvar114 Is Not Empty*

☐ ${e://Field/matchvar114} (205)

*Display this choice:*

*If matchvar115 Is Not Empty*

☐ ${e://Field/matchvar115} (206)

*Display this choice:*

*If matchvar116 Is Not Empty*

☐ ${e://Field/matchvar116} (207)

*Display this choice:*

*If matchvar117 Is Not Empty*

☐ ${e://Field/matchvar117} (208)

*Display this choice:*

*If matchvar118 Is Not Empty*

☐ ${e://Field/matchvar118} (209)

*Display this choice:*

*If matchvar119 Is Not Empty*

☐ ${e://Field/matchvar119} (210)

*Display this choice:*

*If matchvar120 Is Not Empty*

☐ ${e://Field/matchvar120} (211)

*Display this choice:*

*If matchvar121 Is Not Empty*

☐ ${e://Field/matchvar121} (212)

*Display this choice:*

*If matchvar122 Is Not Empty*

☐ ${e://Field/matchvar122} (213)

*Display this choice:*

*If matchvar123 Is Not Empty*

☐ ${e://Field/matchvar123} (214)

*Display this choice:*

*If matchvar124 Is Not Empty*

☐ ${e://Field/matchvar124} (215)

*Display this choice:*

*If matchvar125 Is Not Empty*

☐ ${e://Field/matchvar125} (216)

*Display this choice:*

*If matchvar126 Is Not Empty*

☐ ${e://Field/matchvar126} (217)

*Display this choice:*

*If matchvar127 Is Not Empty*

☐ ${e://Field/matchvar127} (218)

*Display this choice:*

*If matchvar128 Is Not Empty*

☐ ${e://Field/matchvar128} (219)

*Display this choice:*

*If matchvar129 Is Not Empty*

☐ ${e://Field/matchvar129} (220)

*Display this choice:*

*If matchvar130 Is Not Empty*

☐ ${e://Field/matchvar130} (221)

*Display this choice:*

*If matchvar131 Is Not Empty*

☐ ${e://Field/matchvar131} (222)

*Display this choice:*

*If matchvar132 Is Not Empty*

☐ ${e://Field/matchvar132} (223)

*Display this choice:*

*If matchvar133 Is Not Empty*

☐ ${e://Field/matchvar133} (224)

*Display this choice:*

*If matchvar134 Is Not Empty*

☐ ${e://Field/matchvar134} (225)

*Display this choice:*

*If matchvar135 Is Not Empty*

☐ ${e://Field/matchvar135} (226)

*Display this choice:*

*If matchvar136 Is Not Empty*

☐ ${e://Field/matchvar136} (227)

*Display this choice:*

*If matchvar137 Is Not Empty*

☐ ${e://Field/matchvar137} (228)

*Display this choice:*

*If matchvar138 Is Not Empty*

☐ ${e://Field/matchvar138} (229)

*Display this choice:*
*If matchvar139 Is Not Empty*

☐ ${e://Field/matchvar139} (230)

*Display this choice:*
*If matchvar140 Is Not Empty*

☐ ${e://Field/matchvar140} (231)

*Display this choice:*
*If matchvar141 Is Not Empty*

☐ ${e://Field/matchvar141} (232)

*Display this choice:*
*If matchvar142 Is Not Empty*

☐ ${e://Field/matchvar142} (233)

*Display this choice:*
*If matchvar143 Is Not Empty*

☐ ${e://Field/matchvar143} (234)

*Display this choice:*
*If matchvar144 Is Not Empty*

☐ ${e://Field/matchvar144} (235)

*Display this choice:*
*If matchvar145 Is Not Empty*

☐ ${e://Field/matchvar145} (236)

*Display this choice:*
*If matchvar146 Is Not Empty*

☐ ${e://Field/matchvar146} (237)

*Display this choice:*

*If matchvar147 Is Not Empty*

☐ ${e://Field/matchvar147} (238)

*Display this choice:*

*If matchvar148 Is Not Empty*

☐ ${e://Field/matchvar148} (239)

*Display this choice:*

*If matchvar149 Is Not Empty*

☐ ${e://Field/matchvar149} (240)

*Display this choice:*

*If matchvar150 Is Not Empty*

☐ ${e://Field/matchvar150} (241)

*Display this choice:*

*If matchvar151 Is Not Empty*

☐ ${e://Field/matchvar151} (242)

*Display this choice:*

*If matchvar152 Is Not Empty*

☐ ${e://Field/matchvar152} (243)

*Display this choice:*

*If matchvar153 Is Not Empty*

☐ ${e://Field/matchvar153} (244)

*Display this choice:*

*If matchvar154 Is Not Empty*

☐ ${e://Field/matchvar154} (245)

*Display this choice:*

*If matchvar155 Is Not Empty*

☐ ${e://Field/matchvar155} (246)

*Display this choice:*

*If matchvar156 Is Not Empty*

☐ ${e://Field/matchvar156} (248)

*Display this choice:*

*If matchvar157 Is Not Empty*

☐ ${e://Field/matchvar157} (249)

*Display this choice:*

*If matchvar158 Is Not Empty*

☐ ${e://Field/matchvar158} (250)

*Display this choice:*

*If matchvar159 Is Not Empty*

☐ ${e://Field/matchvar159} (251)

*Display this choice:*

*If matchvar160 Is Not Empty*

☐ ${e://Field/matchvar160} (252)

*Display this choice:*

*If matchvar161 Is Not Empty*

☐ ${e://Field/matchvar161} (253)

*Display this choice:*

*If matchvar162 Is Not Empty*

☐ ${e://Field/matchvar162} (254)

*Display this choice:*

*If matchvar163 Is Not Empty*

☐ ${e://Field/matchvar163} (255)

*Display this choice:*

*If matchvar164 Is Not Empty*

☐ ${e://Field/matchvar164} (256)

*Display this choice:*

*If matchvar165 Is Not Empty*

☐ ${e://Field/matchvar165} (257)

*Display this choice:*

*If matchvar166 Is Not Empty*

☐ ${e://Field/matchvar166} (258)

*Display this choice:*

*If matchvar167 Is Not Empty*

☐ ${e://Field/matchvar167} (259)

*Display this choice:*

*If matchvar168 Is Not Empty*

☐ ${e://Field/matchvar168} (260)

*Display this choice:*

*If matchvar169 Is Not Empty*

☐ ${e://Field/matchvar169} (261)

*Display this choice:*

*If matchvar170 Is Not Empty*

☐ ${e://Field/matchvar170} (262)

*Display this choice:*

*If matchvar171 Is Not Empty*

☐ ${e://Field/matchvar171} (263)

*Display this choice:*

*If matchvar172 Is Not Empty*

☐ ${e://Field/matchvar172} (264)

*Display this choice:*

*If matchvar173 Is Not Empty*

☐ ${e://Field/matchvar173} (265)

*Display this choice:*

*If matchvar174 Is Not Empty*

☐ ${e://Field/matchvar174} (266)

*Display this choice:*

*If matchvar175 Is Not Empty*

☐ ${e://Field/matchvar175} (267)

*Display this choice:*

*If matchvar176 Is Not Empty*

☐ ${e://Field/matchvar176} (268)

*Display this choice:*

*If matchvar177 Is Not Empty*

☐ ${e://Field/matchvar177} (269)

*Display this choice:*

*If matchvar178 Is Not Empty*

☐ ${e://Field/matchvar178} (270)

*Display this choice:*

*If matchvar179 Is Not Empty*

☐ ${e://Field/matchvar179} (271)

*Display this choice:*

*If matchvar180 Is Not Empty*

☐ ${e://Field/matchvar180} (272)

*Display this choice:*

*If matchvar181 Is Not Empty*

☐ ${e://Field/matchvar181} (273)

*Display this choice:*
*If matchvar182 Is Not Empty*

☐ ${e://Field/matchvar182} (274)

*Display this choice:*
*If matchvar183 Is Not Empty*

☐ ${e://Field/matchvar183} (275)

*Display this choice:*
*If matchvar184 Is Not Empty*

☐ ${e://Field/matchvar184} (276)

*Display this choice:*
*If matchvar185 Is Not Empty*

☐ ${e://Field/matchvar185} (277)

*Display this choice:*
*If matchvar186 Is Not Empty*

☐ ${e://Field/matchvar186} (278)

*Display this choice:*
*If matchvar187 Is Not Empty*

☐ ${e://Field/matchvar187} (279)

*Display this choice:*
*If matchvar188 Is Not Empty*

☐ ${e://Field/matchvar188} (280)

*Display this choice:*
*If matchvar189 Is Not Empty*

☐ ${e://Field/matchvar189} (281)

*Display this choice:*
*If matchvar190 Is Not Empty*

☐ ${e://Field/matchvar190} (282)

*Display this choice:*
*If matchvar191 Is Not Empty*

☐ ${e://Field/matchvar191} (283)

*Display this choice:*
*If matchvar192 Is Not Empty*

☐ ${e://Field/matchvar192} (284)

*Display this choice:*
*If matchvar193 Is Not Empty*

☐ ${e://Field/matchvar193} (285)

*Display this choice:*
*If matchvar194 Is Not Empty*

☐ ${e://Field/matchvar194} (286)

*Display this choice:*
*If matchvar195 Is Not Empty*

☐ ${e://Field/matchvar195} (287)

*Display this choice:*
*If matchvar196 Is Not Empty*

☐ ${e://Field/matchvar196} (288)

*Display this choice:*
*If matchvar197 Is Not Empty*

☐ ${e://Field/matchvar197} (289)

*Display this choice:*
*If matchvar198 Is Not Empty*

☐ ${e://Field/matchvar198} (290)

*Display this choice:*

If matchvar199 Is Not Empty

☐ ${e://Field/matchvar199} (291)

Display this choice:
If matchvar200 Is Not Empty

☐ ${e://Field/matchvar200} (292)

Display this choice:
If matchvar201 Is Not Empty

☐ ${e://Field/matchvar201} (293)

Display this choice:
If matchvar202 Is Not Empty

☐ ${e://Field/matchvar202} (294)

Display this choice:
If matchvar203 Is Not Empty

☐ ${e://Field/matchvar203} (295)

Display this choice:
If matchvar204 Is Not Empty

☐ ${e://Field/matchvar204} (296)

Display this choice:
If matchvar205 Is Not Empty

☐ ${e://Field/matchvar205} (297)

Display this choice:
If matchvar206 Is Not Empty

☐ ${e://Field/matchvar206} (298)

Display this choice:
If matchvar207 Is Not Empty

☐ ${e://Field/matchvar207} (299)

*Display this choice:*

*If matchvar208 Is Not Empty*

☐ ${e://Field/matchvar208} (300)

*Display this choice:*

*If matchvar209 Is Not Empty*

☐ ${e://Field/matchvar209} (301)

*Display this choice:*

*If matchvar210 Is Not Empty*

☐ ${e://Field/matchvar210} (302)

*Display this choice:*

*If matchvar211 Is Not Empty*

☐ ${e://Field/matchvar211} (303)

*Display this choice:*

*If matchvar212 Is Not Empty*

☐ ${e://Field/matchvar212} (304)

*Display this choice:*

*If matchvar213 Is Not Empty*

☐ ${e://Field/matchvar213} (305)

*Display this choice:*

*If matchvar214 Is Not Empty*

☐ ${e://Field/matchvar214} (306)

*Display this choice:*

*If matchvar215 Is Not Empty*

☐ ${e://Field/matchvar215} (307)

*Display this choice:*

*If matchvar216 Is Not Empty*

${e://Field/matchvar216} (308)

*Display this choice:*
*If matchvar217 Is Not Empty*

${e://Field/matchvar217} (309)

*Display this choice:*
*If matchvar218 Is Not Empty*

${e://Field/matchvar218} (310)

*Display this choice:*
*If matchvar219 Is Not Empty*

${e://Field/matchvar219} (311)

*Display this choice:*
*If matchvar220 Is Not Empty*

${e://Field/matchvar220} (312)

*Display this choice:*
*If matchvar221 Is Not Empty*

${e://Field/matchvar221} (313)

*Display this choice:*
*If matchvar222 Is Not Empty*

${e://Field/matchvar222} (314)

*Display this choice:*
*If matchvar223 Is Not Empty*

${e://Field/matchvar223} (315)

*Display this choice:*
*If matchvar224 Is Not Empty*

${e://Field/matchvar224} (316)

End of Block: What matters most to you: Step 1

---

Start of Block: What matters most to you: Step 2

*Carry Forward Selected Choices from "Below are the things that you listed about yourself and/or your loved one(s) with T1D. Please identify up to 6 things about you and/or your loved one(s) you'd like us to consider when matching you with a group of others (check all that apply)"*

Below are the things you said you'd like us to consider when matching you with a group of others with similar interests or situations.  Please rank the items, with **the most important item on top**. We will try to match you up with people who have the same top characteristics.

*Display this choice:*
*If matchvar1 Is Not Empty*

_______ ${e://Field/matchvar1} (1)

*Display this choice:*
*If matchvar2 Is Not Empty*

_______ ${e://Field/matchvar2} (2)

*Display this choice:*
*If matchvar3 Is Not Empty*

_______ ${e://Field/matchvar3} (3)

*Display this choice:*
*If matchvar4 Is Not Empty*

_______ ${e://Field/matchvar4} (4)

*Display this choice:*
*If matchvar5 Is Not Empty*

_______ ${e://Field/matchvar5} (5)

*Display this choice:*
*If matchvar6 Is Not Empty*

_______ ${e://Field/matchvar6} (6)

*Display this choice:*
*If matchvar7 Is Not Empty*

_______ ${e://Field/matchvar7} (7)

*Display this choice:*
*If matchvar46 Is Not Empty*

_______ ${e://Field/matchvar46} (8)

*Display this choice:*
*If matchvar47 Is Not Empty*

_______ ${e://Field/matchvar47} (9)

*Display this choice:*
*If matchvar48 Is Not Empty*

_______ ${e://Field/matchvar48} (10)

*Display this choice:*
*If matchvar49 Is Not Empty*

_______ ${e://Field/matchvar49} (11)

*Display this choice:*

*If matchvar50 Is Not Empty*

_______ ${e://Field/matchvar50} (12)

*Display this choice:*

*If matchvar8 Is Not Empty*

_______ ${e://Field/matchvar8} (13)

*Display this choice:*

*If matchvar9 Is Not Empty*

_______ ${e://Field/matchvar9} (14)

*Display this choice:*

*If matchvar10 Is Not Empty*

_______ ${e://Field/matchvar10} (15)

*Display this choice:*

*If matchvar11 Is Not Empty*

_______ ${e://Field/matchvar11} (16)

*Display this choice:*

*If matchvar12 Is Not Empty*

_______ ${e://Field/matchvar12} (17)

*Display this choice:*

*If matchvar13 Is Not Empty*

_______ ${e://Field/matchvar13} (18)

*Display this choice:*

*If matchvar14 Is Not Empty*

_______ ${e://Field/matchvar14} (19)

*Display this choice:*

*If matchvar15 Is Not Empty*

_______ ${e://Field/matchvar15} (20)

*Display this choice:*

*If matchvar16 Is Not Empty*

_______ ${e://Field/matchvar16} (21)

*Display this choice:*

*If matchvar17 Is Not Empty*

_______ ${e://Field/matchvar17} (22)

*Display this choice:*

*If matchvar18 Is Not Empty*

_______ ${e://Field/matchvar18} (23)

*Display this choice:*

*If matchvar19 Is Not Empty*

_______ ${e://Field/matchvar19} (24)

*Display this choice:*

*If matchvar20 Is Not Empty*

_______ ${e://Field/matchvar20} (25)

*Display this choice:*

*If matchvar21 Is Not Empty*

_______ ${e://Field/matchvar21} (26)

*Display this choice:*

*If matchvar22 Is Not Empty*

_______ ${e://Field/matchvar22} (27)

*Display this choice:*

*If matchvar23 Is Not Empty*

_______ ${e://Field/matchvar23} (28)

*Display this choice:*

*If matchvar24 Is Not Empty*

_______ ${e://Field/matchvar24} (29)

*Display this choice:*

*If matchvar25 Is Not Empty*

_______ ${e://Field/matchvar25} (30)

*Display this choice:*

*If matchvar26 Is Not Empty*

_______ ${e://Field/matchvar26} (31)

*Display this choice:*

*If matchvar27 Is Not Empty*

_______ ${e://Field/matchvar27} (32)

*Display this choice:*

*If matchvar28 Is Not Empty*

_______ ${e://Field/matchvar28} (33)

*Display this choice:*

*If matchvar29 Is Not Empty*

_______ ${e://Field/matchvar29} (34)

*Display this choice:*

*If matchvar30 Is Not Empty*

_______ ${e://Field/matchvar30} (35)

*Display this choice:*

*If matchvar31 Is Not Empty*

_______ ${e://Field/matchvar31} (36)

*Display this choice:*

*If matchvar32 Is Not Empty*

_______ ${e://Field/matchvar32} (37)

*Display this choice:*

*If matchvar33 Is Not Empty*

_______ ${e://Field/matchvar33} (38)

*Display this choice:*

*If matchvar34 Is Not Empty*

_______ ${e://Field/matchvar34} (39)

*Display this choice:*

*If matchvar35 Is Not Empty*

_______ ${e://Field/matchvar35} (40)

*Display this choice:*

*If matchvar36 Is Not Empty*

_______ ${e://Field/matchvar36} (41)

*Display this choice:*

*If matchvar37 Is Not Empty*

_______ ${e://Field/matchvar37} (42)

*Display this choice:*

*If matchvar38 Is Not Empty*

_______ ${e://Field/matchvar38} (43)

*Display this choice:*

*If matchvar39 Is Not Empty*

_______ ${e://Field/matchvar39} (44)

*Display this choice:*

*If matchvar40 Is Not Empty*

_______ ${e://Field/matchvar40} (45)

*Display this choice:*

*If matchvar41 Is Not Empty*

_______ ${e://Field/matchvar41} (46)

*Display this choice:*

*If matchvar42 Is Not Empty*

_______ ${e://Field/matchvar42} (47)

*Display this choice:*

*If matchvar43 Is Not Empty*

_______ ${e://Field/matchvar43} (48)

*Display this choice:*

*If matchvar44 Is Not Empty*

_______ ${e://Field/matchvar44} (49)

*Display this choice:*

*If matchvar45 Is Not Empty*

_______ ${e://Field/matchvar45} (50)

*Display this choice:*

*If matchvar63 Is Not Empty*

_______ ${e://Field/matchvar63} (51)

*Display this choice:*

*If matchvar63-2 Is Not Empty*

_______ ${e://Field/matchvar63-2} (52)

*Display this choice:*

*If matchvar63-3 Is Not Empty*

_______ ${e://Field/matchvar63-3} (53)

*Display this choice:*

*If matchvar51 Is Not Empty*

_______ ${e://Field/matchvar51} (54)

*Display this choice:*

*If matchvar52 Is Not Empty*

_______ ${e://Field/matchvar52} (55)

*Display this choice:*

*If matchvar53 Is Not Empty*

_______ ${e://Field/matchvar53} (56)

*Display this choice:*

*If matchvar54 Is Not Empty*

_______ ${e://Field/matchvar54} (57)

*Display this choice:*

*If matchvar55 Is Not Empty*

_______ ${e://Field/matchvar55} (58)

*Display this choice:*

*If matchvar56 Is Not Empty*

_______ ${e://Field/matchvar56} (59)

*Display this choice:*

*If matchvar57 Is Not Empty*

_______ ${e://Field/matchvar57} (60)

*Display this choice:*

*If matchvar58 Is Not Empty*

_______ ${e://Field/matchvar58} (61)

*Display this choice:*

*If matchvar59 Is Not Empty*

_______ ${e://Field/matchvar59} (62)

*Display this choice:*

*If matchvar60 Is Not Empty*

_______ ${e://Field/matchvar60} (63)

*Display this choice:*

*If matchvar61 Is Not Empty*

_______ ${e://Field/matchvar61} (64)

*Display this choice:*

*If matchvar62 Is Not Empty*

_______ ${e://Field/matchvar62} (65)

*Display this choice:*

*If matchvar64 Is Not Empty*

_______ ${e://Field/matchvar64} (66)

*Display this choice:*

*If matchvar65 Is Not Empty*

_______ ${e://Field/matchvar65} (67)

*Display this choice:*

*If matchvar66 Is Not Empty*

_______ ${e://Field/matchvar66} (68)

*Display this choice:*

*If matchvar67 Is Not Empty*

_______ ${e://Field/matchvar67} (69)

*Display this choice:*

*If matchvar68 Is Not Empty*

_______ ${e://Field/matchvar68} (70)

*Display this choice:*

*If matchvar69 Is Not Empty*

_______ ${e://Field/matchvar69} (71)

*Display this choice:*

*If matchvar70 Is Not Empty*

_______ ${e://Field/matchvar70} (72)

*Display this choice:*

*If matchvar71 Is Not Empty*

_______ ${e://Field/matchvar71} (73)

*Display this choice:*

*If matchvar72 Is Not Empty*

_______ ${e://Field/matchvar72} (74)

*Display this choice:*

*If matchvar73 Is Not Empty*

_______ ${e://Field/matchvar73} (75)

*Display this choice:*

*If matchvar74 Is Not Empty*

_______ ${e://Field/matchvar74} (76)

*Display this choice:*

*If matchvar75 Is Not Empty*

_______ ${e://Field/matchvar75} (77)

*Display this choice:*

*If matchvar76 Is Not Empty*

_______ ${e://Field/matchvar76} (78)

*Display this choice:*

*If matchvar77 Is Not Empty*

_______ ${e://Field/matchvar77} (79)

*Display this choice:*

*If matchvar78 Is Not Empty*

_______ ${e://Field/matchvar78} (80)

*Display this choice:*

*If matchvar79 Is Not Empty*

_______ ${e://Field/matchvar79} (81)

*Display this choice:*

*If matchvar80 Is Not Empty*

_______ ${e://Field/matchvar80} (82)

*Display this choice:*

*If matchvar81 Is Not Empty*

_______ ${e://Field/matchvar81} (83)

*Display this choice:*

*If matchvar82 Is Not Empty*

_______ ${e://Field/matchvar82} (84)

*Display this choice:*

*If matchvar83 Is Not Empty*

_______ ${e://Field/matchvar83} (85)

*Display this choice:*

*If matchvar84 Is Not Empty*

_______ ${e://Field/matchvar84} (86)

*Display this choice:*

*If matchvar85 Is Not Empty*

_______ ${e://Field/matchvar85} (87)

*Display this choice:*

*If matchvar86 Is Not Empty*

_______ ${e://Field/matchvar86} (88)

*Display this choice:*

*If matchvar87 Is Not Empty*

_______ ${e://Field/matchvar87} (89)

*Display this choice:*

*If matchvar88 Is Not Empty*

_______ ${e://Field/matchvar88} (90)

*Display this choice:*

*If matchvar89 Is Not Empty*

_______ ${e://Field/matchvar89} (91)

*Display this choice:*

*If matchvar90 Is Not Empty*

_______ ${e://Field/matchvar90} (92)

*Display this choice:*

*If matchvar91 Is Not Empty*

_______ ${e://Field/matchvar91} (93)

*Display this choice:*

*If matchvar92 Is Not Empty*

_______ ${e://Field/matchvar92} (94)

*Display this choice:*

*If matchvar93 Is Not Empty*

_______ ${e://Field/matchvar93} (95)

*Display this choice:*

*If matchvar94 Is Not Empty*

_______ ${e://Field/matchvar94} (96)

*Display this choice:*

*If matchvar95 Is Not Empty*

_______ ${e://Field/matchvar95} (97)

*Display this choice:*

*If matchvar96 Is Not Empty*

_______ ${e://Field/matchvar96} (98)

*Display this choice:*

*If matchvar97 Is Not Empty*

_______ ${e://Field/matchvar97} (99)

*Display this choice:*

*If matchvar98 Is Not Empty*

_______ ${e://Field/matchvar98} (100)

*Display this choice:*

*If matchvar99 Is Not Empty*

_______ ${e://Field/matchvar99} (101)

*Display this choice:*

*If matchvar100 Is Not Empty*

_______ ${e://Field/matchvar100} (102)

*Display this choice:*

*If matchvar101 Is Not Empty*

_______ ${e://Field/matchvar101} (103)

*Display this choice:*

*If matchvarW Is Not Empty*

_______ ${e://Field/matchvarW} (104)

*Display this choice:*

*If matchvarW1 Is Not Empty*

_______ ${e://Field/matchvarW1} (105)

*Display this choice:*

*If matchvarW2 Is Not Empty*

_______ ${e://Field/matchvarW2} (106)

*Display this choice:*

*If matchvarEthinicText Is Not Empty*

_______ ${e://Field/matchvarEthinicText} (107)

*Display this choice:*

*If matchvar102 Is Not Empty*

_______ ${e://Field/matchvar102} (108)

*Display this choice:*

*If matchvar103 Is Not Empty*

_______ ${e://Field/matchvar103} (109)

*Display this choice:*

*If matchvaryrlessthan5 Is Not Empty*

_______ ${e://Field/matchvaryrlessthan5} (110)

*Display this choice:*

*If matchvar5-10yrsagofr Is Not Empty*

_______ ${e://Field/matchvar5-10yrsago} (111)

*Display this choice:*

*If matchvarmorethan10yrs Is Not Empty*

_______ ${e://Field/matchvarmorethan10yrs} (112)

*Display this choice:*

*If matchvar104 Is Not Empty*

_______ ${e://Field/matchvar104} (113)

*Display this choice:*

*If matchvar105 Is Not Empty*

_______ ${e://Field/matchvar105} (114)

*Display this choice:*

*If matchvar106 Is Not Empty*

_______ ${e://Field/matchvar106} (115)

*Display this choice:*

*If matchvar107 Is Not Empty*

_______ ${e://Field/matchvar107} (116)

*Display this choice:*

*If matchvar108 Is Not Empty*

_______ ${e://Field/matchvar108} (117)

*Display this choice:*

*If matchvar109 Is Not Empty*

_______ ${e://Field/matchvar109} (118)

*Display this choice:*

*If matchvar110 Is Not Empty*

_______ ${e://Field/matchvar110} (119)

*Display this choice:*

*If matchvar111 Is Not Empty*

_______ ${e://Field/matchvar111} (120)

*Display this choice:*

*If matchvar112 Is Not Empty*

_______ ${e://Field/matchvar112} (121)

*Display this choice:*

*If matchvar113 Is Not Empty*

_______ ${e://Field/matchvar113} (122)

*Display this choice:*

*If matchvar114 Is Not Empty*

_______ ${e://Field/matchvar114} (123)

*Display this choice:*

*If matchvar115 Is Not Empty*

_______ ${e://Field/matchvar115} (124)

*Display this choice:*

*If matchvar116 Is Not Empty*

_______ ${e://Field/matchvar116} (125)

*Display this choice:*

*If matchvar117 Is Not Empty*

_______ ${e://Field/matchvar117} (126)

*Display this choice:*

*If matchvar118 Is Not Empty*

_______ ${e://Field/matchvar118} (127)

*Display this choice:*

*If matchvar119 Is Not Empty*

_______ ${e://Field/matchvar119} (128)

*Display this choice:*

*If matchvar120 Is Not Empty*

_______ ${e://Field/matchvar120} (129)

*Display this choice:*

*If matchvar121 Is Not Empty*

_______ ${e://Field/matchvar121} (130)

*Display this choice:*

*If matchvar122 Is Not Empty*

_______ ${e://Field/matchvar122} (131)

*Display this choice:*

*If matchvar123 Is Not Empty*

_______ ${e://Field/matchvar123} (132)

*Display this choice:*

*If matchvar124 Is Not Empty*

_______ ${e://Field/matchvar124} (133)

*Display this choice:*

*If matchvar125 Is Not Empty*

_______ ${e://Field/matchvar125} (134)

*Display this choice:*

*If matchvar126 Is Not Empty*

_______ ${e://Field/matchvar126} (135)

*Display this choice:*

*If matchvar127 Is Not Empty*

_______ ${e://Field/matchvar127} (136)

*Display this choice:*

*If matchvar128 Is Not Empty*

_______ ${e://Field/matchvar128} (137)

*Display this choice:*

*If matchvar129 Is Not Empty*

_______ ${e://Field/matchvar129} (138)

*Display this choice:*

*If matchvar130 Is Not Empty*

_______ ${e://Field/matchvar130} (139)

*Display this choice:*

*If matchvar131 Is Not Empty*

_______ ${e://Field/matchvar131} (140)

*Display this choice:*

*If matchvar132 Is Not Empty*

_______ ${e://Field/matchvar132} (141)

*Display this choice:*

*If matchvar133 Is Not Empty*

_______ ${e://Field/matchvar133} (142)

*Display this choice:*

*If matchvar134 Is Not Empty*

_______ ${e://Field/matchvar134} (143)

*Display this choice:*

*If matchvar135 Is Not Empty*

_______ ${e://Field/matchvar135} (144)

*Display this choice:*

*If matchvar136 Is Not Empty*

_______ ${e://Field/matchvar136} (145)

*Display this choice:*

*If matchvar137 Is Not Empty*

_______ ${e://Field/matchvar137} (146)

*Display this choice:*

*If matchvar138 Is Not Empty*

_______ ${e://Field/matchvar138} (147)

*Display this choice:*

*If matchvar139 Is Not Empty*

_______ ${e://Field/matchvar139} (148)

*Display this choice:*

*If matchvar140 Is Not Empty*

_______ ${e://Field/matchvar140} (149)

*Display this choice:*

*If matchvar141 Is Not Empty*

_______ ${e://Field/matchvar141} (150)

*Display this choice:*

*If matchvar142 Is Not Empty*

_______ ${e://Field/matchvar142} (151)

*Display this choice:*
*If matchvar143 Is Not Empty*

_______ ${e://Field/matchvar143} (152)

*Display this choice:*
*If matchvar144 Is Not Empty*

_______ ${e://Field/matchvar144} (153)

*Display this choice:*
*If matchvar145 Is Not Empty*

_______ ${e://Field/matchvar145} (154)

*Display this choice:*
*If matchvar146 Is Not Empty*

_______ ${e://Field/matchvar146} (155)

*Display this choice:*
*If matchvar147 Is Not Empty*

_______ ${e://Field/matchvar147} (156)

*Display this choice:*
*If matchvar148 Is Not Empty*

_______ ${e://Field/matchvar148} (157)

*Display this choice:*
*If matchvar149 Is Not Empty*

_______ ${e://Field/matchvar149} (158)

*Display this choice:*
*If matchvar150 Is Not Empty*

_______ ${e://Field/matchvar150} (159)

*Display this choice:*
*If matchvar151 Is Not Empty*

_______ ${e://Field/matchvar151} (160)

*Display this choice:*
*If matchvar152 Is Not Empty*

_______ ${e://Field/matchvar152} (161)

*Display this choice:*
*If matchvar153 Is Not Empty*

_______ ${e://Field/matchvar153} (162)

*Display this choice:*

*If matchvar154 Is Not Empty*

_______ ${e://Field/matchvar154} (163)

*Display this choice:*

*If matchvar155 Is Not Empty*

_______ ${e://Field/matchvar155} (164)

*Display this choice:*

*If matchvar156 Is Not Empty*

_______ ${e://Field/matchvar156} (165)

*Display this choice:*

*If matchvar157 Is Not Empty*

_______ ${e://Field/matchvar157} (166)

*Display this choice:*

*If matchvar158 Is Not Empty*

_______ ${e://Field/matchvar158} (167)

*Display this choice:*

*If matchvar159 Is Not Empty*

_______ ${e://Field/matchvar159} (168)

*Display this choice:*

*If matchvar160 Is Not Empty*

_______ ${e://Field/matchvar160} (169)

*Display this choice:*

*If matchvar161 Is Not Empty*

_______ ${e://Field/matchvar161} (170)

*Display this choice:*

*If matchvar162 Is Not Empty*

_______ ${e://Field/matchvar162} (171)

*Display this choice:*

*If matchvar163 Is Not Empty*

_______ ${e://Field/matchvar163} (172)

*Display this choice:*

*If matchvar164 Is Not Empty*

_______ ${e://Field/matchvar164} (173)

*Display this choice:*

*If matchvar165 Is Not Empty*

_______ ${e://Field/matchvar165} (174)

*Display this choice:*

*If matchvar166 Is Not Empty*

_______ ${e://Field/matchvar166} (175)

*Display this choice:*

*If matchvar167 Is Not Empty*

_______ ${e://Field/matchvar167} (176)

*Display this choice:*

*If matchvar168 Is Not Empty*

_______ ${e://Field/matchvar168} (177)

*Display this choice:*

*If matchvar169 Is Not Empty*

_______ ${e://Field/matchvar169} (178)

*Display this choice:*

*If matchvar170 Is Not Empty*

_______ ${e://Field/matchvar170} (179)

*Display this choice:*

*If matchvar171 Is Not Empty*

_______ ${e://Field/matchvar171} (180)

*Display this choice:*

*If matchvar172 Is Not Empty*

_______ ${e://Field/matchvar172} (181)

*Display this choice:*

*If matchvar173 Is Not Empty*

_______ ${e://Field/matchvar173} (182)

*Display this choice:*

*If matchvar174 Is Not Empty*

_______ ${e://Field/matchvar174} (183)

*Display this choice:*

*If matchvar175 Is Not Empty*

_______ ${e://Field/matchvar175} (184)

*Display this choice:*

*If matchvar176 Is Not Empty*

_______ ${e://Field/matchvar176} (185)

*Display this choice:*

*If matchvar177 Is Not Empty*

_______ ${e://Field/matchvar177} (186)

*Display this choice:*

*If matchvar178 Is Not Empty*

_______ ${e://Field/matchvar178} (187)

*Display this choice:*

*If matchvar179 Is Not Empty*

_______ ${e://Field/matchvar179} (188)

*Display this choice:*

*If matchvar180 Is Not Empty*

_______ ${e://Field/matchvar180} (189)

*Display this choice:*

*If matchvar181 Is Not Empty*

_______ ${e://Field/matchvar181} (190)

*Display this choice:*

*If matchvar182 Is Not Empty*

_______ ${e://Field/matchvar182} (191)

*Display this choice:*

*If matchvar183 Is Not Empty*

_______ ${e://Field/matchvar183} (192)

*Display this choice:*

*If matchvar184 Is Not Empty*

_______ ${e://Field/matchvar184} (193)

*Display this choice:*

*If matchvar185 Is Not Empty*

_______ ${e://Field/matchvar185} (194)

*Display this choice:*

*If matchvar186 Is Not Empty*

_______ ${e://Field/matchvar186} (195)

*Display this choice:*

*If matchvar187 Is Not Empty*

_______ ${e://Field/matchvar187} (196)

*Display this choice:*

*If matchvar188 Is Not Empty*

_______ ${e://Field/matchvar188} (197)

*Display this choice:*

*If matchvar189 Is Not Empty*

_______ ${e://Field/matchvar189} (198)

*Display this choice:*

*If matchvar190 Is Not Empty*

_______ ${e://Field/matchvar190} (199)

*Display this choice:*

*If matchvar191 Is Not Empty*

_______ ${e://Field/matchvar191} (200)

*Display this choice:*

*If matchvar192 Is Not Empty*

_______ ${e://Field/matchvar192} (201)

*Display this choice:*

*If matchvar193 Is Not Empty*

_______ ${e://Field/matchvar193} (202)

*Display this choice:*

*If matchvar194 Is Not Empty*

_______ ${e://Field/matchvar194} (203)

*Display this choice:*

*If matchvar195 Is Not Empty*

_______ ${e://Field/matchvar195} (204)

*Display this choice:*

*If matchvar196 Is Not Empty*

_______ ${e://Field/matchvar196} (205)

*Display this choice:*

*If matchvar197 Is Not Empty*

_______ ${e://Field/matchvar197} (206)

*Display this choice:*

*If matchvar198 Is Not Empty*

_______ ${e://Field/matchvar198} (207)

*Display this choice:*

*If matchvar199 Is Not Empty*

_______ ${e://Field/matchvar199} (208)

*Display this choice:*

*If matchvar200 Is Not Empty*

_______ ${e://Field/matchvar200} (209)

*Display this choice:*

*If matchvar201 Is Not Empty*

_______ ${e://Field/matchvar201} (210)

*Display this choice:*

*If matchvar202 Is Not Empty*

_______ ${e://Field/matchvar202} (211)

*Display this choice:*

*If matchvar203 Is Not Empty*

_______ ${e://Field/matchvar203} (212)

*Display this choice:*

*If matchvar204 Is Not Empty*

_______ ${e://Field/matchvar204} (213)

*Display this choice:*

*If matchvar205 Is Not Empty*

_______ ${e://Field/matchvar205} (214)

*Display this choice:*

*If matchvar206 Is Not Empty*

_______ ${e://Field/matchvar206} (215)

*Display this choice:*

*If matchvar207 Is Not Empty*

_______ ${e://Field/matchvar207} (216)

*Display this choice:*

*If matchvar208 Is Not Empty*

_______ ${e://Field/matchvar208} (217)

*Display this choice:*

*If matchvar209 Is Not Empty*

_______ ${e://Field/matchvar209} (218)

*Display this choice:*

*If matchvar210 Is Not Empty*

_______ ${e://Field/matchvar210} (219)

*Display this choice:*

*If matchvar211 Is Not Empty*

_______ ${e://Field/matchvar211} (220)

*Display this choice:*

*If matchvar212 Is Not Empty*

_______ ${e://Field/matchvar212} (221)

*Display this choice:*

*If matchvar213 Is Not Empty*

_______ ${e://Field/matchvar213} (222)

*Display this choice:*

*If matchvar214 Is Not Empty*

_______ ${e://Field/matchvar214} (223)

*Display this choice:*

*If matchvar215 Is Not Empty*

_______ ${e://Field/matchvar215} (224)

*Display this choice:*

*If matchvar216 Is Not Empty*

_______ ${e://Field/matchvar216} (225)

*Display this choice:*

*If matchvar217 Is Not Empty*

_______ ${e://Field/matchvar217} (226)

*Display this choice:*

*If matchvar218 Is Not Empty*

_______ ${e://Field/matchvar218} (227)

*Display this choice:*

*If matchvar219 Is Not Empty*

_______ ${e://Field/matchvar219} (228)

*Display this choice:*

*If matchvar220 Is Not Empty*

_______ ${e://Field/matchvar220} (229)

*Display this choice:*

*If matchvar221 Is Not Empty*

_______ ${e://Field/matchvar221} (230)

*Display this choice:*

*If matchvar222 Is Not Empty*

_______ ${e://Field/matchvar222} (231)

*Display this choice:*

*If matchvar223 Is Not Empty*

_______ ${e://Field/matchvar223} (232)

*Display this choice:*

*If matchvar224 Is Not Empty*

_______ ${e://Field/matchvar224} (233)

End of Block: What matters most to you: Step 2

Start of Block: Your availability

People have different schedules and routines. Many people have time constraints due to family, work, or other life commitments. We understand that people may: be early birds who would love to chat at 5 a.m., be night owls who are typically awake and ready to talk at 11 p.m., have flexible schedules that allow them to meet mid-afternoon, not know their work schedule in advance, and so on. We want to group people who have similar schedules, preferences, and availability.

What time zone are you in?

○ Pacific Time PT (1)

○ Mountain Time MT (4)

○ Central Time CT (5)

○ Eastern Time ET (6)

○ Atlantic Time AT (7)

○ Newfoundland Standard Time NST (8)

○ Other (9) ________________________________________________

When could you usually participate in a 1-hour virtual meet-up, about once a month?  By checking a time slot, you are saying that time slot could usually work for you, even if you are not available every single time.

| | Mondays (1) | Tuesdays (2) | Wednesdays (3) | Thursdays (4) | Fridays (5) | Saturdays (6) | Sundays (7) |
|---|---|---|---|---|---|---|---|
| Midnight - 01:00 AM (1) | ☐ | ☐ | ☐ | ☐ | ☐ | ☐ | ☐ |
| 01:00 AM - 02:00 AM (37) | ☐ | ☐ | ☐ | ☐ | ☐ | ☐ | ☐ |
| 02:00 AM - 03:00 AM (38) | ☐ | ☐ | ☐ | ☐ | ☐ | ☐ | ☐ |
| 03:00 AM - 04:00 AM (39) | ☐ | ☐ | ☐ | ☐ | ☐ | ☐ | ☐ |
| 04:00 AM - 05:00 AM (40) | ☐ | ☐ | ☐ | ☐ | ☐ | ☐ | ☐ |
| 05:00 AM - 06:00 AM (41) | ☐ | ☐ | ☐ | ☐ | ☐ | ☐ | ☐ |
| 06:00 AM - 07:00 AM (42) | ☐ | ☐ | ☐ | ☐ | ☐ | ☐ | ☐ |
| 07:00 AM - 08:00 AM (43) | ☐ | ☐ | ☐ | ☐ | ☐ | ☐ | ☐ |
| 08:00 AM - 09:00 AM (44) | ☐ | ☐ | ☐ | ☐ | ☐ | ☐ | ☐ |
| 09:00 AM - 10:00 AM (45) | ☐ | ☐ | ☐ | ☐ | ☐ | ☐ | ☐ |

| | | | | | | | |
|---|---|---|---|---|---|---|---|
| 10:00 AM - 11:00 AM (46) | ☐ | ☐ | ☐ | ☐ | ☐ | ☐ | ☐ |
| 11:00 AM - 12:00 PM (47) | ☐ | ☐ | ☐ | ☐ | ☐ | ☐ | ☐ |
| 12:00 PM - 01:00 PM (48) | ☐ | ☐ | ☐ | ☐ | ☐ | ☐ | ☐ |
| 01:00 PM - 02:00 PM (49) | ☐ | ☐ | ☐ | ☐ | ☐ | ☐ | ☐ |
| 02:00 PM - 03:00 PM (50) | ☐ | ☐ | ☐ | ☐ | ☐ | ☐ | ☐ |
| 03:00 PM - 04:00 PM (51) | ☐ | ☐ | ☐ | ☐ | ☐ | ☐ | ☐ |
| 04:00 PM - 05:00 PM (52) | ☐ | ☐ | ☐ | ☐ | ☐ | ☐ | ☐ |
| 05:00 PM - 06:00 PM (53) | ☐ | ☐ | ☐ | ☐ | ☐ | ☐ | ☐ |
| 06:00 PM - 07:00 PM (54) | ☐ | ☐ | ☐ | ☐ | ☐ | ☐ | ☐ |
| 07:00 PM - 08:00 PM (55) | ☐ | ☐ | ☐ | ☐ | ☐ | ☐ | ☐ |
| 08:00 PM - 09:00 PM (56) | ☐ | ☐ | ☐ | ☐ | ☐ | ☐ | ☐ |

| | | | | | | | |
|---|---|---|---|---|---|---|---|
| 09:00 PM - 10:00 PM (57) | ☐ | ☐ | ☐ | ☐ | ☐ | ☐ | ☐ |
| 10:00 PM - 11:00 PM (58) | ☐ | ☐ | ☐ | ☐ | ☐ | ☐ | ☐ |
| 11:00 PM - Midnight (59) | ☐ | ☐ | ☐ | ☐ | ☐ | ☐ | ☐ |

Comments about your availability (optional)

________________________________________________________________

End of Block: Your availability

Start of Block: When you'll be matched with a group

Thank you for all your answers. We will do our best to find you a group that matches what matters to you as quickly as possible. Our goal is to find a group for everyone within a month, but for people who have less availability or uncommon priorities, it may take longer. Either way, we will keep you informed by email. To help us meet your needs, please tell us which you would prefer. "Better match" means you'd prefer to wait longer to find a group of people with whom I have more things in common. "Faster match" means you'd prefer to find a group quickly even if it means you have less in common with the other group members. "Neutral" means you have no preference between these two options.

| | Better match | Neutral | Faster match |
|---|---|---|---|
| Slide to choose () | | | |

End of Block: When you'll be matched with a group

Start of Block: Peer group facilitator

**We need peer group facilitators!**
A **peer group facilitator** is a volunteer who will start monthly meetings and help make sure the conversation is enjoyable for everyone, including themselves. Each small group will have one or two peer group facilitators. Are you willing to facilitate a small group? This is a volunteer position. You would be provided with support, training, access to CommuniT1D researchers and staff to answer your questions, and a network of other peer group facilitators. This network will allow all peer group facilitators to share ideas and strategies for making group discussions good for everyone.

○ Yes, I'm definitely interested! No hesitation! (1)

○ Yes, I'm interested, but I'd like to know more before I commit. Please send me more information. (4)

○ No, I am not interested. (5)

○ I prefer not to answer (6)

End of Block: Peer group facilitator

---

Start of Block: Your hopes

Please tell us how much you agree or disagree with the following statements. **I hope that by participating in CommuniT1D, I will …**

*Display this choice:*

*If What is your relationship to diabetes? (check all that apply) = I have a loved one with T1D (or another rare type of diabetes like LADA, MODY, or type 3c)*

*Display this choice:*

*If What is your relationship to diabetes? (check all that apply) = I have a loved one with T1D (or another rare type of diabetes like LADA, MODY, or type 3c)*

*Display this choice:*

*If What is your relationship to diabetes? (check all that apply) = I have a loved one with T1D (or another rare type of diabetes like LADA, MODY, or type 3c)*

| | Strongly disagree (1) | Disagree (2) | Neither agree nor disagree (3) | Agree (4) | Strongly agree (5) |
|---|---|---|---|---|---|
| Be more physically active with T1D or help my loved one be more physically active with T1D (1) | ○ | ○ | ○ | ○ | ○ |
| Build community (4) | ○ | ○ | ○ | ○ | ○ |
| Communicate better with others about T1D (e.g., teach another adult how to care for my child, talk to friends/family/teachers/coworkers/etc. about T1D) (5) | ○ | ○ | ○ | ○ | ○ |
| Feel less alone (6) | ○ | ○ | ○ | ○ | ○ |
| Have less conflict in my family/immediate circle (7) | ○ | ○ | ○ | ○ | ○ |
| Increase my T1D knowledge (8) | ○ | ○ | ○ | ○ | ○ |
| Improve my or my loved one's blood glucose numbers (9) | ○ | ○ | ○ | ○ | ○ |
| *Display this choice: If What is your relationship to diabetes? (check all that apply) = I have a loved one with T1D (or another rare type of diabetes like LADA, MODY, or type 3c)* Improve my loved one's mental health (10) | ○ | ○ | ○ | ○ | ○ |
| Improve my mental health (11) | ○ | ○ | ○ | ○ | ○ |

| | | | | | |
|---|---|---|---|---|---|
| *Display this choice:* *If What is your relationship to diabetes? (check all that apply) = I have a loved one with T1D (or another rare type of diabetes like LADA, MODY, or type 3c)* Improve my loved one’s overall well-being (12) | ○ | ○ | ○ | ○ | ○ |
| Improve my overall well-being (13) | ○ | ○ | ○ | ○ | ○ |
| *Display this choice:* *If What is your relationship to diabetes? (check all that apply) = I have a loved one with T1D (or another rare type of diabetes like LADA, MODY, or type 3c)* Improve my loved one’s quality of life (14) | ○ | ○ | ○ | ○ | ○ |
| Improve my quality of life (15) | ○ | ○ | ○ | ○ | ○ |
| Learn more T1D tips, tricks, and/or coping strategies (16) | ○ | ○ | ○ | ○ | ○ |
| Live more easily with T1D (17) | ○ | ○ | ○ | ○ | ○ |
| Make friends (18) | ○ | ○ | ○ | ○ | ○ |
| Manage stress better (19) | ○ | ○ | ○ | ○ | ○ |
| Reduce the burden T1D puts on me (20) | ○ | ○ | ○ | ○ | ○ |
| Sleep better (21) | ○ | ○ | ○ | ○ | ○ |
| Other (22) | ○ | ○ | ○ | ○ | ○ |
| I prefer not to answer (23) | ○ | ○ | ○ | ○ | ○ |

End of Block: Your hopes

Start of Block: We need your help

**We need your help** CommuniT1D is funded as a research project. **The research project is led by adults with T1D, parents of children with T1D, scientists, doctors, and other experts.** (Read more about our team here: https://communit1d.ca/about-us/) We want to make CommuniT1D a welcoming and helpful program for everyone. By submitting your answers to the previous questions, you give us permission to analyze anonymized versions (no identifying information included) of your answers to help improve the program. We will **never** share information that could identify you. To further help us make CommuniT1D a successful and sustainable program for people across Canada dealing with T1D, we would really appreciate you agreeing to be part of the research study about CommuniT1D. This is optional. You can participate in CommuniT1D without being part of the research study. Being part of the research study means: about every 6 months, we will invite you to fill out a 15- to 20-minute survey specific to type 1 diabetes and CommuniT1D, and we might invite you to participate in a research interview in 2026 or 2027 The survey questions have all been selected by adults with T1D and parents of children with T1D, who deemed the questions interesting and relevant to life with T1D. At the end of the survey, you will have the option to download a report of your answers if you like. **Everyone who participates in the research study will have the option to be entered in a draw for a $250 gift card each time they complete a survey.** Each time you complete a survey, with your permission, your name is entered in the draw and will stay in the draw throughout the multiyear study. This means that the more surveys you complete, the more times your name will be in the draw. We will randomly draw one or more names every 6 months. You will also have the option to see your answers each time you complete a survey. This means that, if you like, we will provide you with a personalized, confidential report of your answers. You can download the report if you like. **It would really help us and others with T1D if you agree to be part of the research study.**

End of Block: We need your help

Start of Block: Interested in study

Are you interested in participating in the research study?

○ Yes, I'm interested. Show me the consent form so I can make an informed decision to participate or not. (1)

○ No, I'm not interested in considering this right now. (4)

*Display this question:*

*If Are you interested in participating in the research study? = No, I’m not interested in considering this right now.*

Thank you for letting us know. If you are comfortable sharing, we would be interested to know why you are not interested in being part of the research study:

________________________________________________________________

________________________________________________________________

________________________________________________________________

________________________________________________________________

________________________________________________________________

**End of Block: Interested in study**
