## Appendix 2 (English only) for "A Study Protocol for CommuniT1D : A Customized Virtual Peer Support Program For People Living With Type 1 Diabetes In Canada"

### communiT1D_outcome_measures

Start of Block: consent

**Find Your CommuniT1D: Customized Virtual Peer Support for People Living with Type 1 Diabetes PROJECT DESCRIPTION AND INFORMED CONSENT FORM PARTICIPATING ADULTS About the researcher and team** This project is directed by Holly Witteman, PhD, full professor and scientist in the Faculty of Medicine at Laval University in Quebec City. She has lived with type 1 diabetes (T1D) since childhood. The project's steering committee has 14 members. Ten of them manage T1D daily—their own and/or their child's. You can learn more about the project team here: https://communit1d.ca/about-us/ This project is funded by the Canadian Institutes of Health Research (CIHR) with a matched grant from JDRF Canada. JDRF was formerly known as the Juvenile Diabetes Research Foundation. In 2024, JDRF changed its name to Breakthrough T1D. Before you agree to participate in this project, please take the time to read and understand the following information. This document provides information on the goals of this project, what is involved, and risks and benefits of participating. **Overview of the project** The purpose of this project, "Find Your CommuniT1D: Customized Virtual Peer Support for People Living with Type 1 Diabetes," is to develop and evaluate an online program to improve the mental health and wellbeing of people whose lives are affected by type 1 diabetes (T1D) or similar types of diabetes (LADA, MODY, type 3c) across Canada. The program's main focus will be regular meetings (about once a month) with small groups of people (about 4-6 people on average) who all live with T1D or a similar type of diabetes or who have a loved one (a child, a spouse, etc.) who lives with T1D or a similar type of diabetes. The program will also host regular open webinars with experts. People attending webinars will be able to ask questions if they like. The project is funded for an initial 4-year period. **What your participation entails** Participating in this project involves: First, completing a short online questionnaire so we can match you with other people who share other characteristics or situations (e.g., preferred language, ethnocultural group, stage of life). Once we match you with a group, you then attend one or more meetings online via Zoom, Webex, or a similar technology. Please let us know if the group is not a good fit for you. In that case, we will help you find a different group. Second, choosing to attend webinars or not. These are completely optional. Third and finally, helping us evaluate and improve CommuniT1D by completing very short (2 minute) meeting feedback forms after each monthly meeting, completing longer (20 minute) online surveys twice a year, and possibly receiving invitations to participate in one or two interviews sometime in the next several years. **Possible benefits, inconveniences, and risks related to your participation** The **benefits** to participating in this project are:
Connecting to others whose lives are affected by T1D or similar types of diabetes.
Accessing additional resources like webinars with experts. Connecting to others with shared experiences. Making friends. Feeling less alone. Helping to keep CommuniT1D going. As a research project, we need people to participate so that we can fund

the program beyond the initial 4-year grant.  Contributing to knowledge about how peer support might help people whose lives are affected by T1D. This can help others with T1D in the future. The **inconveniences** are:     Participating in the different activities (small group meetings, webinars, answering surveys, participating in interviews) will take time. We know that T1D and similar types of diabetes also consume a lot of time and energy. You can decide how involved you would like to be (number of groups or other activities).  The **risks** are:          Other CommuniT1D members or webinar experts may share information that is incorrect or does not apply to you or your loved one. Please always consult your health care team before making changes to your or your loved one's diabetes management. Please feel free to let the CommuniT1D team know if someone is sharing incorrect medical information.     Discussing health issues during meetings may cause some anxiety. If so, please do not hesitate to talk to your health care provider. Trustworthy information about T1D and mental health is also available at this address: https://jdrf.ca/life-with-t1d/mental-health/  *This research project is directed by Holly Witteman, PhD, professor and researcher at the Faculty of Medicine, Université Laval. This project is approved by the Research Ethics Committee of the CIUSSS Capitale-Nationale Approval number: CIUSSSCN- 2024-2887*

---

**Voluntary participation**  Your participation in this 4-year project is entirely voluntary and not compensated. You can decide whether or not to participate. You can also withdraw at any time. You can participate in CommuniT1D without being in the research study. To do this, you would attend group meetings and webinars but not fill out the surveys or participate in interviews. **Confidentiality and data protection**  All the information we collect will be anonymous and used for research purposes only. In other words:     No individual information will be presented in reports, publications or presentations.      All data will be presented in aggregate form without individual identifiers.   Data will be stored on Qualtrics secure servers located in Canada and on a secure server at Université Laval, Québec City, Québec.  Interview recordings will be stored on a secure server located on the campus of Université Laval, Québec City, Québec. Only members of the research team will have access to the recordings and data.        All research data will be kept for ongoing monitoring and evaluation purposes, and then destroyed 7 years after the end of the project.  **Acknowledgment**  We value your participation in this project and thank you for agreeing to participate.    **Contact person**  If you have any questions about the project, or if you experience a problem as a result of participating in the project, please contact the research team at.    **Complaints**  For ethical questions, please contact the research project management office at:   To file a complaint about this project, please contact the CIUSSS de la Capitale Nationale Ombudsperson at:  Tel: 418-691-0762  Teletype service: 418-649-3734     *This research project is directed by Holly Witteman, PhD, professor and researcher at the Faculty of Medicine, Université Laval. TThis project is approved by the Research Ethics Committee of the CIUSSS Capitale-Nationale Approval number: CIUSSSCN- 2024-2887*

---

By submitting this online form (by clicking “Next” or the right-pointing arrow), you agree to following: I have been informed about the nature and purpose of this project and the research procedures. I was informed in English, which is a language in which I am proficient. I have been informed of the benefits, inconveniences, and risks of participating. I understand that my participation in this project is voluntary and that I can withdraw at any time. I understand that the information collected is confidential and will only be used for research purposes. I was invited to ask questions and I am satisfied with the answers. Agreeing to participate in this project does not release the researchers from their obligations towards me. I know I will not be compensated for participating in this project. I understand that I can download or print this page to keep for my records, so that I have a copy of the informed consent form. I have read the project description and informed consent form and I voluntarily agree to participate in this project. I agree to participate not only in project activities but also in the evaluation of the project.

*This research project is directed by Holly Witteman, PhD, professor and researcher at the Faculty of Medicine, Université Laval. This project is approved by the Research Ethics Committee of the CIUSSS Capitale-Nationale Approval number: CIUSSSCN- 2024-2887*

---

Please enter your first name:

________________________________________________________________

---

Please enter your last (family) name:

________________________________________________________________

---

JS

Please enter your email address:

○ Email: (1) __________________________________________________

---

JS *

Please confirm your email address:

◯ Email (confirm): (1) ________________________________________________

Your help means a lot to us. Thank you for agreeing to participate in this project.

End of Block: consent

---

Start of Block: About you

First, we need to know a few things about you to make sure we ask you the right questions.

Do you yourself have type 1 diabetes (T1D) or a similar type of diabetes, such as LADA, MODY, or type 3c? (choose one)

◯ Yes (1)

◯ No (2)

◯ Prefer not to answer (4)

*Display this question:*

*If Do you yourself have type 1 diabetes (T1D) or a similar type of diabetes, such as LADA, MODY, or... = Yes*

To which age group do you belong? (choose one)

◯ 18-64 years old (4)

◯ 65+ years old (5)

◯ Prefer not to answer (6)

Are you a caregiver of a loved one with type 1 diabetes (T1D) or a similar type of diabetes, such as LADA, MODY, or type 3c? (choose one)

- ◯ Yes (4)
- ◯ No (5)
- ◯ Prefer not to answer (6)

---

Do you have a partner/spouse who has type 1 diabetes (T1D) or a similar type of diabetes, such as LADA, MODY, or type 3c? (choose one)

- ◯ Yes (4)
- ◯ No (5)
- ◯ Prefer not to answer (6)

---

Are you the parent or guardian of one or more children under age 18 with type 1 diabetes (T1D) or a similar type of diabetes, such as LADA, MODY, or type 3c? (choose one)

- ◯ Yes (4)
- ◯ No (5)
- ◯ Prefer not to answer (6)

---

*Display this question:*

*If Are you the parent or guardian of one or more children under age 18 with type 1 diabetes (T1D) or... = Yes*

How old is/are your child/children with type 1 diabetes (T1D) or a similar type of diabetes, such as LADA, MODY, or type 3c? (choose all that apply)

- ☐ Under 1 year old (11)
- ☐ 1 year old (12)
- ☐ 2 years old (13)
- ☐ 3-4 years old (14)
- ☐ 5-12 years old (15)
- ☐ 13 years old (16)
- ☐ 14-17 years old (17)
- ☐ Prefer not to answer (18)

---

*Display this question:*

*If Are you the parent or guardian of one or more children under age 18 with type 1 diabetes (T1D) or... = Yes*

How many children under age 18 with type 1 diabetes (T1D) or a similar type of diabetes (such as LADA, MODY, or type 3c) do you have? (choose one)

- ○ 1 (1)
- ○ 2 (2)
- ○ 3 (3)
- ○ More than 3 (4)
- ○ Prefer not to answer (5)

---

Are you employed? This includes full-time, part-time, self-employed, and other forms of employment. (choose one)

◯ Yes (4)

◯ No (5)

◯ Prefer not to answer (6)

End of Block: About you

---

Start of Block: Identification

*Display this question:*

*If If False*

Please enter your email address to help us make sure we are connecting your responses over time. Reminder: your data are protected and confidential.

◯ Your email address (1) ________________________________________________

◯ Your email address (repeat) (4) ________________________________________________

End of Block: Identification

---

Start of Block: Type 1 Diabetes Distress Scale

*Display this question:*

*If Do you yourself have type 1 diabetes (T1D) or a similar type of diabetes, such as LADA, MODY, or... = Yes*

Living with type 1 diabetes can be tough. Listed below are a variety of distressing things that many people with type 1 diabetes experience. Thinking back **over the past month**, please indicate the degree to which each of the following may have been a problem for you by selecting the appropriate number. For example, if you feel that a particular item was not a problem for you over the past month, you would select the option "1". If it was very tough for you over the past month, you might select the option "6".

| | Not a problem (1) | A slight problem (2) | A moderate problem (3) | A somewhat serious problem (4) | A serious problem (5) | A very serious problem (6) |
|---|---|---|---|---|---|---|
| Feeling that I am not as skilled at managing diabetes as I should be. (1) | ○ | ○ | ○ | ○ | ○ | ○ |
| Feeling that I don’t eat as carefully as I probably should. (2) | ○ | ○ | ○ | ○ | ○ | ○ |
| Feeling that I don’t notice the warning signs of hypoglycemia as well as I used to. (3) | ○ | ○ | ○ | ○ | ○ | ○ |
| Feeling that people treat me differently when they find out I have diabetes. (4) | ○ | ○ | ○ | ○ | ○ | ○ |
| Feeling discouraged when I see high blood glucose numbers that I can’t explain. (5) | ○ | ○ | ○ | ○ | ○ | ○ |
| Feeling that my family and friends make a bigger deal out of diabetes than they should. (6) | ○ | ○ | ○ | ○ | ○ | ○ |

| | | | | | | |
|---|---|---|---|---|---|---|
| Feeling that I can’t tell my diabetes doctor what is really on my mind. (7) | ○ | ○ | ○ | ○ | ○ | ○ |
| Feeling that I am not taking as much insulin as I should. (8) | ○ | ○ | ○ | ○ | ○ | ○ |
| Feeling that there is too much diabetes equipment and stuff I must always have with me. (9) | ○ | ○ | ○ | ○ | ○ | ○ |
| Feeling like I have to hide my diabetes from other people. (10) | ○ | ○ | ○ | ○ | ○ | ○ |
| Feeling that my friends and family worry more about hypoglycemia (low blood sugar) than I want them to. (11) | ○ | ○ | ○ | ○ | ○ | ○ |
| Feeling that I don’t check my blood glucose level as often as I probably should. (12) | ○ | ○ | ○ | ○ | ○ | ○ |

| | | | | | | |
|---|---|---|---|---|---|---|
| Feeling worried that I will develop serious long-term complications, no matter how hard I try. (13) | ○ | ○ | ○ | ○ | ○ | ○ |
| Feeling that I don’t get help I really need from my diabetes doctor about managing diabetes. (14) | ○ | ○ | ○ | ○ | ○ | ○ |
| Feeling frightened that I could have a serious hypoglycemic (low blood sugar) event when I’m asleep. (15) | ○ | ○ | ○ | ○ | ○ | ○ |
| Feeling that thoughts about food and eating control my life. (16) | ○ | ○ | ○ | ○ | ○ | ○ |
| Feeling that my friends or family treat me as if I were more fragile or sicker than I really am. (17) | ○ | ○ | ○ | ○ | ○ | ○ |

| | | | | | | |
|---|---|---|---|---|---|---|
| Feeling that my diabetes doctor doesn't really understand what it's like to have diabetes. (18) | ○ | ○ | ○ | ○ | ○ | ○ |
| Feeling concerned that diabetes may make me less attractive to employers. (19) | ○ | ○ | ○ | ○ | ○ | ○ |
| Feeling that my friends or family act like “diabetes police.” (20) | ○ | ○ | ○ | ○ | ○ | ○ |
| Feeling that I’ve got to be perfect with my diabetes management. (21) | ○ | ○ | ○ | ○ | ○ | ○ |
| Feeling frightened that I could have a serious hypoglycemic (low blood sugar) event while driving. (22) | ○ | ○ | ○ | ○ | ○ | ○ |
| Feeling that my eating is out of control. (23) | ○ | ○ | ○ | ○ | ○ | ○ |
| Feeling that people will think less of me if they knew I had diabetes. (24) | ○ | ○ | ○ | ○ | ○ | ○ |

| | | | | | | |
|---|---|---|---|---|---|---|
| Feeling that no matter how hard I try with my diabetes, it will never be good enough. (25) | ○ | ○ | ○ | ○ | ○ | ○ |
| Feeling that my diabetes doctor doesn't know enough about diabetes and diabetes care. (26) | ○ | ○ | ○ | ○ | ○ | ○ |
| Feeling that I can't ever be safe from the possibility of a serious hypoglycemic (low blood sugar) event. (27) | ○ | ○ | ○ | ○ | ○ | ○ |
| Feeling that I don't give my diabetes as much attention as I probably should. (28) | ○ | ○ | ○ | ○ | ○ | ○ |

**End of Block: Type 1 Diabetes Distress Scale**

---

**Start of Block: Parent_DDS_teen**

*Display this question:*

*If How old is/are your child/children with type 1 diabetes (T1D) or a similar type of diabetes, such... = 13 years old*

*Or How old is/are your child/children with type 1 diabetes (T1D) or a similar type of diabetes, such... = 14-17 years old*

The following questions ask about how you have been feeling as a parent of a teen with diabetes. For each item, select the response that gives the best answer for you. Please provide an answer for each question. During the past month, I have been:

| | Not at all (1) | A little (2) | Somewhat (3) | A lot (4) | A great deal (5) |
|---|---|---|---|---|---|
| Feeling that my teen and I just don't work well together when it comes to diabetes. (1) | ○ | ○ | ○ | ○ | ○ |
| Feeling unappreciated for all the ways I try to help my teen manage diabetes. (2) | ○ | ○ | ○ | ○ | ○ |
| Feeling that I can't trust my teen to take good care of their diabetes. (3) | ○ | ○ | ○ | ○ | ○ |
| Worrying about my teen's low blood sugars when they are away from home. (4) | ○ | ○ | ○ | ○ | ○ |
| Worrying that my teen will ignore or forget diabetes if I don't keep reminding them. (5) | ○ | ○ | ○ | ○ | ○ |
| Feeling that diabetes is taking up too much of my mental and physical energy every day. (6) | ○ | ○ | ○ | ○ | ○ |

| | | | | | |
|---|---|---|---|---|---|
| Feeling that trying to help my teen with their diabetes is always a battle. (7) | ○ | ○ | ○ | ○ | ○ |
| Worrying about my teen's low blood sugars when they are sleeping. (8) | ○ | ○ | ○ | ○ | ○ |
| Feeling that no one notices that diabetes is hard on me, not just on my teen. (9) | ○ | ○ | ○ | ○ | ○ |
| Feeling that my teen doesn't do enough to manage their diabetes. (10) | ○ | ○ | ○ | ○ | ○ |
| Worrying that my teen doesn't have the right doctor for them. (11) | ○ | ○ | ○ | ○ | ○ |
| Worrying that others will blame me if my teen's diabetes is not well-controlled. (12) | ○ | ○ | ○ | ○ | ○ |
| Worrying that my teen will soon leave home and I cannot protect them. (13) | ○ | ○ | ○ | ○ | ○ |

| | | | | | |
|---|---|---|---|---|---|
| Frustrated because my teen ignores my suggestions about diabetes. (14) | ○ | ○ | ○ | ○ | ○ |
| Frustrated by the lack of understanding and support for diabetes I get from friends and family members. (15) | ○ | ○ | ○ | ○ | ○ |
| Worrying that my teen doesn't get all of the expert medical help they need. (16) | ○ | ○ | ○ | ○ | ○ |
| Feeling uncertain about how to motivate my teen to take better care of their diabetes. (17) | ○ | ○ | ○ | ○ | ○ |
| Concerned that my teen is not prepared to deal with the world of insurance and doctors once they are an adult. (18) | ○ | ○ | ○ | ○ | ○ |

| | | | | | |
|---|---|---|---|---|---|
| Frustrated that I am the only one who takes responsibility for helping my teen manage diabetes. (19) | o | o | o | o | o |
| Worrying that my nagging about diabetes is hurting my relationship with my teen. (20) | o | o | o | o | o |
| Worrying that my teen will have diabetes-related complications in the future. (21) | o | o | o | o | o |

End of Block: Parent_DDS_teen

---

Start of Block: Parent_DDS_5-12

*Display this question:*

*If How old is/are your child/children with type 1 diabetes (T1D) or a similar type of diabetes, such... = 5-12 years old*

The following questions ask about how you have been feeling as a parent of a child with diabetes. For each item, select the response that gives the best answer for you. Please provide an answer for each question. During the past month, I have been:

| | Not at all (1) | A little (2) | Somewhat (3) | A lot (4) | A great deal (5) |
|---|---|---|---|---|---|
| Feeling that other caregivers in my child’s life (e.g., my child’s other parent, school staff, other relatives, etc.) and I just don't work well together when it comes to diabetes. (1) | ○ | ○ | ○ | ○ | ○ |
| Feeling unappreciated for all the ways I try to help my child manage diabetes. (2) | ○ | ○ | ○ | ○ | ○ |
| Feeling that I can't trust others to take good care of my child’s diabetes if I am not there. (3) | ○ | ○ | ○ | ○ | ○ |
| Worrying about my child's blood sugars when they are away from home (e.g., at school, at an activity, at a friend’s house, at another relative’s house). (4) | ○ | ○ | ○ | ○ | ○ |

| | | | | | |
|---|---|---|---|---|---|
| Worrying that my child will ignore or forget diabetes if I don't keep reminding them. (5) | ○ | ○ | ○ | ○ | ○ |
| Feeling that diabetes is taking up too much of my mental and physical energy every day. (6) | ○ | ○ | ○ | ○ | ○ |
| Feeling that trying to help my child with their diabetes is always a battle. (7) | ○ | ○ | ○ | ○ | ○ |
| Worrying about my child's low blood sugars when they are sleeping. (8) | ○ | ○ | ○ | ○ | ○ |
| Feeling that no one notices that diabetes is hard on me, not just on my child. (9) | ○ | ○ | ○ | ○ | ○ |
| Feeling that other adults in my child's life don't do enough to manage my child's diabetes. (10) | ○ | ○ | ○ | ○ | ○ |

| | | | | | |
|---|---|---|---|---|---|
| Worrying that my child doesn't have the right doctor for them. (11) | ○ | ○ | ○ | ○ | ○ |
| Worrying that others will blame me if my child's diabetes is not well-controlled. (12) | ○ | ○ | ○ | ○ | ○ |
| Worrying that my child will soon be more independent and I won't be able to protect them. (13) | ○ | ○ | ○ | ○ | ○ |
| Frustrated because other people ignore my suggestions about diabetes while my child is in their care. (14) | ○ | ○ | ○ | ○ | ○ |
| Frustrated by the lack of understanding and support for diabetes I get from friends and family members. (15) | ○ | ○ | ○ | ○ | ○ |

| | | | | | |
|---|---|---|---|---|---|
| Worrying that my child doesn't get all of the expert medical help they need. (16) | ○ | ○ | ○ | ○ | ○ |
| Feeling uncertain about how to motivate my child to take better care of their diabetes. (17) | ○ | ○ | ○ | ○ | ○ |
| Concerned that my child will not be prepared to deal with the world of insurance and doctors once they are an adult. (18) | ○ | ○ | ○ | ○ | ○ |
| Frustrated that I am the only one who takes responsibility for helping my child manage diabetes. (19) | ○ | ○ | ○ | ○ | ○ |
| Worrying that my “helicopter parenting” about diabetes is hurting my relationship with my child. (20) | ○ | ○ | ○ | ○ | ○ |

| | | | | | |
|---|---|---|---|---|---|
| Worrying that my child will have diabetes-related complications in the future. (21) | o | o | o | o | o |

End of Block: Parent_DDS_5-12

---

Start of Block: Parent_DDS_0-4

*Display this question:*

*If How old is/are your child/children with type 1 diabetes (T1D) or a similar type of diabetes, such... = Under 1 year old*

*Or How old is/are your child/children with type 1 diabetes (T1D) or a similar type of diabetes, such... = 1 year old*

*Or How old is/are your child/children with type 1 diabetes (T1D) or a similar type of diabetes, such... = 2 years old*

*Or How old is/are your child/children with type 1 diabetes (T1D) or a similar type of diabetes, such... = 3-4 years old*

The following questions ask about how you have been feeling as a parent of a child with diabetes. For each item, select the response that gives the best answer for you. Please provide an answer for each question. During the past month, I have been:

| | Not at all (1) | A little (2) | Somewhat (3) | A lot (4) | A great deal (5) |
|---|---|---|---|---|---|
| Feeling that other caregivers in my child’s life (e.g., my child’s other parent, school staff, other relatives, etc.) and I just don't work well together when it comes to diabetes. (22) | ○ | ○ | ○ | ○ | ○ |
| Feeling unappreciated for all the ways I try to help my child manage diabetes. (23) | ○ | ○ | ○ | ○ | ○ |
| Feeling that I can't trust others to take good care of my child’s diabetes if I am not there. (24) | ○ | ○ | ○ | ○ | ○ |
| Worrying about my child's blood sugars when they are away from home (e.g., at daycare, preschool, or school, at an activity, at a friend’s house, at another relative’s house). (25) | ○ | ○ | ○ | ○ | ○ |

| | | | | | |
|---|---|---|---|---|---|
| Worrying that other caregivers (e.g., my child’s other parent, daycare workers, other relatives, friends, etc.) will ignore or forget diabetes if I don't keep reminding them. (26) | ○ | ○ | ○ | ○ | ○ |
| Feeling that diabetes is taking up too much of my mental and physical energy every day. (27) | ○ | ○ | ○ | ○ | ○ |
| Feeling that trying to help my child with their diabetes is always a battle. (28) | ○ | ○ | ○ | ○ | ○ |
| Worrying about my child's low blood sugars when they are sleeping. (29) | ○ | ○ | ○ | ○ | ○ |
| Feeling that no one notices that diabetes is hard on me, not just on my child. (30) | ○ | ○ | ○ | ○ | ○ |

| | | | | | |
|---|---|---|---|---|---|
| Feeling that other adults in my child’s life don't do enough to manage my child’s diabetes. (31) | ○ | ○ | ○ | ○ | ○ |
| Worrying that my child doesn't have the right doctor for them. (32) | ○ | ○ | ○ | ○ | ○ |
| Worrying that others will blame me if my child's diabetes is not well-controlled. (33) | ○ | ○ | ○ | ○ | ○ |
| Worrying that my child will soon transition beyond early childhood care (for example, they may start school) and I won’t be able to protect them. (34) | ○ | ○ | ○ | ○ | ○ |
| Frustrated because other people ignore my suggestions about diabetes while my child is in their care. (35) | ○ | ○ | ○ | ○ | ○ |

| | | | | | |
|---|---|---|---|---|---|
| Frustrated by the lack of understanding and support for diabetes I get from friends and family members. (36) | ○ | ○ | ○ | ○ | ○ |
| Worrying that my child doesn't get all of the expert medical help they need. (37) | ○ | ○ | ○ | ○ | ○ |
| Feeling uncertain about how to motivate other caregivers (e.g., my child's other parent, daycare workers, other relatives, etc.) to take better care of my child's diabetes. (38) | ○ | ○ | ○ | ○ | ○ |
| Concerned that my child will not be prepared to deal with the world of insurance and doctors once they are an adult. (39) | ○ | ○ | ○ | ○ | ○ |

| | | | | | |
|---|---|---|---|---|---|
| Frustrated that I am the only one who takes responsibility for helping my child manage diabetes. (40) | ○ | ○ | ○ | ○ | ○ |
| Worrying that my constant involvement with diabetes is hurting my relationships with other people in my life. (41) | ○ | ○ | ○ | ○ | ○ |
| Worrying that my child will have diabetes-related complications in the future. (42) | ○ | ○ | ○ | ○ | ○ |

End of Block: Parent_DDS_0-4

---

Start of Block: Parent_DDS_partner

*Display this question:*

*If Do you have a partner/spouse who has type 1 diabetes (T1D) or a similar type of diabetes, such as... = Yes*

The following questions ask about how you have been feeling as a spouse or partner of someone with diabetes. For each item, circle the number that gives the best answer for you. Please provide an answer for each question. During the past month, I have been:

| | Not at all (1) | A little (2) | Somewhat (3) | A lot (4) | A great deal (5) |
|---|---|---|---|---|---|
| Worrying about my partner's low blood sugars. (1) | ○ | ○ | ○ | ○ | ○ |
| Feeling unclear about exactly how much I should be involved in managing my partner's diabetes. (2) | ○ | ○ | ○ | ○ | ○ |
| Frustrated that my partner shuts me out of their diabetes. (3) | ○ | ○ | ○ | ○ | ○ |
| Feeling that my partner doesn't try hard enough to manage their diabetes. (4) | ○ | ○ | ○ | ○ | ○ |
| Feeling overwhelmed by the constant demands of my partner's diabetes. (5) | ○ | ○ | ○ | ○ | ○ |
| Worrying that I don't know how to best help my partner manage diabetes. (6) | ○ | ○ | ○ | ○ | ○ |

| | | | | | |
|---|---|---|---|---|---|
| Feeling that I stay silent about my partner's diabetes more than I really should. (7) | ○ | ○ | ○ | ○ | ○ |
| Feeling that diabetes is taking up too much of my mental and physical energy every day. (8) | ○ | ○ | ○ | ○ | ○ |
| Feeling that no one notices that diabetes is hard on me, not just on my partner. (9) | ○ | ○ | ○ | ○ | ○ |
| Frustrated that the more I try to help my partner manage their diabetes, the worse things get between us. (10) | ○ | ○ | ○ | ○ | ○ |
| Feeling guilty about not doing enough to help my partner with diabetes. (11) | ○ | ○ | ○ | ○ | ○ |
| Frustrated that I can’t get my partner to improve their attitude about diabetes. (12) | ○ | ○ | ○ | ○ | ○ |

| | | | | | |
|---|---|---|---|---|---|
| Worrying that I am failing to help my partner manage diabetes more successfully. (13) | ○ | ○ | ○ | ○ | ○ |
| Feeling that trying to help my partner with their diabetes is always a battle. (14) | ○ | ○ | ○ | ○ | ○ |
| Frustrated because my partner ignores my suggestions about diabetes. (15) | ○ | ○ | ○ | ○ | ○ |
| Frustrated that diabetes often interrupts our plans. (16) | ○ | ○ | ○ | ○ | ○ |
| Worrying about my partner's low blood sugars when they are sleeping. (17) | ○ | ○ | ○ | ○ | ○ |
| Worrying about my partner's driving because of possible low blood sugars. (18) | ○ | ○ | ○ | ○ | ○ |

| | | | | | |
|---|---|---|---|---|---|
| Worrying about leaving my partner alone because of the possible danger of low blood sugars. (19) | ○ | ○ | ○ | ○ | ○ |
| Concerned that my partner and I are not working well together when it comes to diabetes. (20) | ○ | ○ | ○ | ○ | ○ |
| Feeling that I never get a break from worrying about my partner's diabetes. (21) | ○ | ○ | ○ | ○ | ○ |

**End of Block: Parent_DDS_partner**

**Start of Block: Work related diabetes distress**

*Display this question:*

*If Do you yourself have type 1 diabetes (T1D) or a similar type of diabetes, such as LADA, MODY, or... = Yes*

*Or Are you the parent or guardian of one or more children under age 18 with type 1 diabetes (T1D) or... = Yes*

*And If*

*Are you employed? This includes full-time, part-time, self-employed, and other forms of employmen... = Yes*

Which of the following issues are currently a problem for you?

*Display this choice:*

*If Do you yourself have type 1 diabetes (T1D) or a similar type of diabetes, such as LADA, MODY, or... = Yes*

*Display this choice:*

*If Are you the parent or guardian of one or more children under age 18 with type 1 diabetes (T1D) or... = Yes*

| | Not a problem (6) | A slight problem (7) | A moderate problem (8) | A somewhat serious problem (9) | A serious problem (10) |
|---|---|---|---|---|---|
| *Display this choice: If Do you yourself have type 1 diabetes (T1D) or a similar type of diabetes, such as LADA, MODY, or... = Yes* Worrying about your ability to do your job due to your diabetes (1) | ○ | ○ | ○ | ○ | ○ |
| *Display this choice: If Are you the parent or guardian of one or more children under age 18 with type 1 diabetes (T1D) or... = Yes* Worrying about your ability to do your job due to your child's diabetes (2) | ○ | ○ | ○ | ○ | ○ |
| Often feeling exhausted by simultaneously reconciling work and diabetes. (3) | ○ | ○ | ○ | ○ | ○ |

**End of Block: Work related diabetes distress**

Start of Block: Kingston Caregiver Stress Scale

*Display this question:*

*If Are you a caregiver of a loved one with type 1 diabetes (T1D) or a similar type of diabetes, such... = Yes*

Some people report feelings of stress surrounding certain aspects of caregiving. To what extent, if any, do these apply to you in your role of care giving to your spouse or relative? Using a 5 point rating scale, where 1 equals no stress and 5 equals extreme stress, indicate the extent of the stress or frustration you feel surrounding the following issues.

Page Break

*Display this question:*

*If Are you a caregiver of a loved one with type 1 diabetes (T1D) or a similar type of diabetes, such... = Yes*

TO WHAT EXTENT...

| | Feeling NO Stress (Coping fine, no problems) (1) | Some Stress (2) | Moderate Stress (3) | A lot of Stress (4) | Extreme Stress (Feeling at “end of rope”, health at risk) (5) |
|---|---|---|---|---|---|
| Are you having feelings of being overwhelmed, overworked, and/or over burdened? (1) | ○ | ○ | ○ | ○ | ○ |
| Has there been a change in your relationship with your loved one? (2) | ○ | ○ | ○ | ○ | ○ |
| Have you noticed any changes in your social life? (3) | ○ | ○ | ○ | ○ | ○ |
| Are you having any conflicts with your previous daily commitments (work/volunteering)? (4) | ○ | ○ | ○ | ○ | ○ |
| Do you have feelings of being confined or trapped by the responsibilities or demands of caregiving? (5) | ○ | ○ | ○ | ○ | ○ |
| Do you ever have feelings related to a lack of confidence in your ability to provide care? (6) | ○ | ○ | ○ | ○ | ○ |
| Do you have concerns regarding the future care needs of your loved one? (7) | ○ | ○ | ○ | ○ | ○ |

Page Break

*Display this question:*

*If Are you a caregiver of a loved one with type 1 diabetes (T1D) or a similar type of diabetes, such... = Yes*

TO WHAT EXTENT...

| | Feeling NO Stress (Coping fine, no problems) (1) | Some Stress (2) | Moderate Stress (3) | A lot of Stress (4) | Extreme Stress (Feeling at “end of rope”, health at risk) (5) |
| --- | --- | --- | --- | --- | --- |
| Are you having any conflicts within your family over care decisions? (1) | ○ | ○ | ○ | ○ | ○ |
| Are you having any conflicts within your family over the amount of support you are receiving in providing care? (2) | ○ | ○ | ○ | ○ | ○ |

Page Break

*Display this question:*

*If Are you a caregiver of a loved one with type 1 diabetes (T1D) or a similar type of diabetes, such... = Yes*

TO WHAT EXTENT...

| | Feeling NO Stress (Coping fine, no problems) (1) | Some Stress (2) | Moderate Stress (3) | A lot of Stress (4) | Extreme Stress (Feeling at “end of rope”, health at risk) (5) |
|---|---|---|---|---|---|
| Are you having any financial difficulties associated with caregiving? (1) | ○ | ○ | ○ | ○ | ○ |

End of Block: Kingston Caregiver Stress Scale

Start of Block: Hospital Anxiety and Depression Scale (HADS)

Please read each item below and tick the box that comes closest to how you have been feeling **in the past week.**

X→

I feel tense or 'wound up'

○ Most of the time (3)

○ A lot of the time (2)

○ Occasionally (1)

○ Not at all (4)

I still enjoy the things I used to enjoy

- Definitely as much (0)
- Not quite so much (1)
- Only a little (2)
- Hardly at all (3)

---

I get a sort of frightened feeling as if something awful is about to happen

- Very definitely and quite badly (3)
- Yes, but not too badly (2)
- A little, but it doesn't worry me (1)
- Not at all (0)

---

I can laugh and see the funny side of things

- As much as always (0)
- Not quite so much now (1)
- Definitely not so much now (2)
- Not at all (3)

---

Worrying thoughts go through my mind

- A great deal of the time (3)
- A lot of the time (2)
- Not too often (1)
- Very little (0)

---

I feel cheerful

- Never (3)
- Not often (2)
- Sometimes (1)
- Most of the time (0)

---

I can sit at ease and feel relaxed

- Definitely (0)
- Usually (1)
- Not often (2)
- Not at all (3)

---

I feel as if I am slowed down

- Nearly all the time (3)
- Very often (2)
- Sometimes (1)
- Not at all (0)

---

X→

I get a sort of frightened feeling like ‘butterflies' in the stomach

- Not at all (0)
- Occasionally (1)
- Quite often (2)
- Very often (3)

---

X→

I have lost interest in my appearance

- Definitely (3)
- I don’t care as much as I should (2)
- I may not take as much care (1)
- I take just as much care as ever (0)

---

X→

I feel restless as if I have to be on the move

- Very much indeed (3)
- Quite a lot (2)
- Not very much (1)
- Not at all (0)

I look forward to enjoyment to things

- As much as I ever did (0)
- Rather less that I used to (1)
- Definitely less than I used to (2)
- Hardly at all (3)

I get sudden feelings of panic

- Very often indeed (3)
- Quite often (2)
- Not very often (1)
- Not at all (0)

I can enjoy a good book or radio or TV programme

- Often (0)
- Sometimes (1)
- Not often (2)
- Very seldom (3)

End of Block: Hospital Anxiety and Depression Scale (HADS)

---

Start of Block: Unmet needs for care

*Display this question:*

*If Do you yourself have type 1 diabetes (T1D) or a similar type of diabetes, such as LADA, MODY, or... = Yes*

During the past 6 months, was there ever a time when you felt that you needed care for your diabetes but didn't receive it?

- Yes (1)
- No (2)

*Display this question:*

*If Are you the parent or guardian of one or more children under age 18 with type 1 diabetes (T1D) or... = Yes*

During the past 6 months, was there ever a time when you felt that your loved one needed care for their diabetes but didn't receive it?

- Yes (1)
- No (2)

*Display this question:*

*If During the past 6 months, was there ever a time when you felt that you needed care for your diabe... = Yes*

Thinking of the most recent time, why didn't you get care?

________________________________________________________________

________________________________________________________________

________________________________________________________________

________________________________________________________________

________________________________________________________________

*Display this question:*

*If During the past 6 months, was there ever a time when you felt that your loved one needed care for... = Yes*

Thinking of the most recent time, why didn't your loved one get care?

________________________________________________________________

________________________________________________________________

________________________________________________________________

________________________________________________________________

________________________________________________________________

**End of Block: Unmet needs for care**

**Start of Block: The Confidence in Diabetes Scale**

I believe I can ...

| | 1 No, I am sure I cannot (1) | 2 (2) | 3 (3) | 4 (4) | 5 Yes, I am sure I can (5) |
|---|---|---|---|---|---|
| Plan my meals and snacks according to dietary guidelines. (21) | ○ | ○ | ○ | ○ | ○ |
| Check my blood glucose at least two times a day. (22) | ○ | ○ | ○ | ○ | ○ |
| Perform the prescribed number of daily insulin injections or insulin pump boluses. (23) | ○ | ○ | ○ | ○ | ○ |
| Adjust my insulin for exercise, traveling, or celebrations. (24) | ○ | ○ | ○ | ○ | ○ |
| Adjust my insulin when I am sick. (25) | ○ | ○ | ○ | ○ | ○ |
| Detect high levels of blood glucose in time to correct. (26) | ○ | ○ | ○ | ○ | ○ |
| Detect low levels of blood glucose in time to correct. (27) | ○ | ○ | ○ | ○ | ○ |
| Treat a high blood glucose correctly. (28) | ○ | ○ | ○ | ○ | ○ |
| Treat a low blood glucose correctly. (29) | ○ | ○ | ○ | ○ | ○ |
| Keep daily records of my blood glucose. (30) | ○ | ○ | ○ | ○ | ○ |

| | | | | | |
|---|---|---|---|---|---|
| Decide when it’s necessary to contact my doctor or diabetes educator. (31) | ○ | ○ | ○ | ○ | ○ |
| Ask my doctor questions about my treatment plan. (32) | ○ | ○ | ○ | ○ | ○ |
| Keep my blood glucose in the normal range when under stress. (33) | ○ | ○ | ○ | ○ | ○ |
| Check my feet for sores or blisters every day. (34) | ○ | ○ | ○ | ○ | ○ |
| Ask my friends or relatives for help with my diabetes. (35) | ○ | ○ | ○ | ○ | ○ |
| Inform colleagues/others of my diabetes, if needed. (36) | ○ | ○ | ○ | ○ | ○ |
| Keep my medical appointments. (37) | ○ | ○ | ○ | ○ | ○ |
| Exercise two to three times weekly. (38) | ○ | ○ | ○ | ○ | ○ |
| Figure out what foods to eat when dining out. (39) | ○ | ○ | ○ | ○ | ○ |
| Read and hear about diabetes complications without getting discouraged. (40) | ○ | ○ | ○ | ○ | ○ |

**End of Block: The Confidence in Diabetes Scale**

Start of Block: Short form of the Hypoglycaemia Fear Survey II

Below is a list of things people with diabetes sometimes do in order to avoid low blood sugar and its consequences. Choose one of the numbers to the right that best describes what you have done during the last 6 months in your daily routine to AVOID low blood sugar and its consequences. (Please do not skip any!)

X→

To avoid low blood sugar and how it affects me I ...

| | Never (0) | Rarely (1) | Sometimes (2) | Often (3) | Almost always (4) |
|---|---|---|---|---|---|
| Limited my out of town travel. (1) | ○ | ○ | ○ | ○ | ○ |
| Avoided visiting friends. (2) | ○ | ○ | ○ | ○ | ○ |
| Made sure there were other people around. (3) | ○ | ○ | ○ | ○ | ○ |
| Kept my blood sugar higher than usual in social situations. (4) | ○ | ○ | ○ | ○ | ○ |
| Kept my blood sugar higher than usual when doing important tasks. (5) | ○ | ○ | ○ | ○ | ○ |

Page Break

Below is a list of concerns people with diabetes sometimes have about low blood sugar. Please read each item carefully (do not skip any). Choose one of the numbers to the right that best describes how often in the last 6 months you WORRIED about each item because of low blood sugar.

X→

Because my blood sugar could go low, I worried about ...

| | Never (0) | Rarely (1) | Sometimes (2) | Often (3) | Almost always (4) |
|---|---|---|---|---|---|
| Not recognizing/realizing I was having low blood sugar. (1) | ○ | ○ | ○ | ○ | ○ |
| Passing out in public. (2) | ○ | ○ | ○ | ○ | ○ |
| Having a hypoglycemic episode while driving. (3) | ○ | ○ | ○ | ○ | ○ |
| Low blood sugar interfering with important things I was doing. (4) | ○ | ○ | ○ | ○ | ○ |
| Becoming hypoglycemic during sleep. (5) | ○ | ○ | ○ | ○ | ○ |
| Getting emotionally upset and difficult to deal with. (6) | ○ | ○ | ○ | ○ | ○ |

**End of Block: Short form of the Hypoglycaemia Fear Survey II**

**Start of Block: WHO-5 Well-being Index**

X→

Please respond to each item by indicating how you felt in the last two weeks.

| | All of the time (5) | Most of the time (4) | More than half the time (3) | Less than half the time (2) | Some of the time (1) | At no time (0) |
|---|---|---|---|---|---|---|
| I have felt cheerful and in good spirits. (1) | ○ | ○ | ○ | ○ | ○ | ○ |
| I have felt calm and relaxed. (2) | ○ | ○ | ○ | ○ | ○ | ○ |
| I have felt active and vigorous. (3) | ○ | ○ | ○ | ○ | ○ | ○ |
| I woke up feeling fresh and rested. (4) | ○ | ○ | ○ | ○ | ○ | ○ |
| My daily life has been filled with things that interest me. (5) | ○ | ○ | ○ | ○ | ○ | ○ |

**End of Block: WHO-5 Well-being Index**

**Start of Block: ADDQoL**

ADDQoL questions removed from appendix to respect licensing and permissions.

**End of Block: ADDQoL**

**Start of Block: Social Provisions Scale**

X→

The next questions are about your current relationships with friends, family members, co-workers, community members, and so on. Please indicate to what extent each statement describes your current relationships with other people.

| | Strongly disagree (1) | Disagree (2) | Agree (3) | Strongly agree (4) |
|---|---|---|---|---|
| I have close relationships that provide me with a sense of emotional security and well-being. (1) | o | o | o | o |
| There is someone I could talk to about important decisions in my life. (2) | o | o | o | o |
| I have relationships where my competence and skill are recognized. (3) | o | o | o | o |
| I feel part of a group of people who share my attitudes and beliefs. (4) | o | o | o | o |
| There are people I can count on in an emergency. (5) | o | o | o | o |

End of Block: Social Provisions Scale

Start of Block: Self-Compassion Scale Short Form

X→

HOW I TYPICALLY ACT TOWARDS MYSELF IN DIFFICULT TIMES. Please read each statement carefully before answering. Indicate how often you behave in the stated manner.

| | Almost never (1) | 2 (2) | 3 (3) | 4 (4) | Almost always (5) |
|---|---|---|---|---|---|
| When I fail at something important to me I become consumed by feelings of inadequacy. (1) | o | o | o | o | o |
| I try to be understanding and patient towards those aspects of my personality I don't like. (2) | o | o | o | o | o |
| When something painful happens I try to take a balanced view of the situation. (3) | o | o | o | o | o |
| When I'm feeling down, I tend to feel like most other people are probably happier than I am. (4) | o | o | o | o | o |
| I try to see my failings as part of the human condition. (5) | o | o | o | o | o |
| When I'm going through a very hard time, I give myself the caring and tenderness I need. (6) | o | o | o | o | o |

| | | | | | |
|---|---|---|---|---|---|
| When something upsets me I try to keep my emotions in balance. (7) | ○ | ○ | ○ | ○ | ○ |
| When I fail at something that's important to me, I tend to feel alone in my failure. (8) | ○ | ○ | ○ | ○ | ○ |
| When I'm feeling down I tend to obsess and fixate on everything that's wrong. (9) | ○ | ○ | ○ | ○ | ○ |
| When I feel inadequate in some way, I try to remind myself that feelings of inadequacy are shared by most people. (10) | ○ | ○ | ○ | ○ | ○ |
| I'm disapproving and judgmental about my own flaws and inadequacies. (11) | ○ | ○ | ○ | ○ | ○ |
| I'm intolerant and impatient towards those aspects of my personality I don't like. (12) | ○ | ○ | ○ | ○ | ○ |

End of Block: Self-Compassion Scale Short Form

---

Start of Block: Ten-Item Personality Inventory- TIPI

Here are a number of personality traits that may or may not apply to you. Please indicate the extent to which you agree or disagree the traits apply to you. You should rate the extent to which the pair of traits applies to you, even if one characteristic applies more strongly than the other. I see myself as:

| | Disagree strongly (9) | Disagree moderately (10) | Disagree a little (11) | Neither agree nor disagree (12) | Agree a little (13) | Agree moderately (14) | Agree strongly (15) |
|---|---|---|---|---|---|---|---|
| Extraverted, enthusiastic. (1) | ○ | ○ | ○ | ○ | ○ | ○ | ○ |
| Critical, quarrelsome. (2) | ○ | ○ | ○ | ○ | ○ | ○ | ○ |
| Dependable, self-disciplined. (3) | ○ | ○ | ○ | ○ | ○ | ○ | ○ |
| Anxious, easily upset. (4) | ○ | ○ | ○ | ○ | ○ | ○ | ○ |
| Open to new experiences, complex. (5) | ○ | ○ | ○ | ○ | ○ | ○ | ○ |
| Reserved, quiet. (6) | ○ | ○ | ○ | ○ | ○ | ○ | ○ |
| Sympathetic, warm. (7) | ○ | ○ | ○ | ○ | ○ | ○ | ○ |
| Disorganized, careless. (8) | ○ | ○ | ○ | ○ | ○ | ○ | ○ |
| Calm, emotionally stable. (9) | ○ | ○ | ○ | ○ | ○ | ○ | ○ |
| Conventional, uncreative. (10) | ○ | ○ | ○ | ○ | ○ | ○ | ○ |

**End of Block: Ten-Item Personality Inventory- TIPI**

---

**Start of Block: CommuniT1D check-in measure**

Please tell us how much you agree or disagree with the following statements. Participating in CommuniT1D is helping me to …

| | Completely disagree | Neither agree nor disagree | Completely agree |
|---|---|---|---|
| | 0 | | 100 |

| |
|---|
| be more physically active with T1D or help my loved one be more physically active with T1D () |
| build community () |
| communicate better with others about T1D (e.g., teach another adult how to care for my child, talk to friends/family/teachers/coworkers/etc. about T1D) () |
| feel less alone () |
| have less conflict in my family/immediate circle () |
| increase my T1D knowledge () |
| improve my blood glucose numbers () |
| improve my loved one's blood glucose numbers () |
| improve my loved one's mental health () |
| improve my mental health () |
| improve my loved one's overall well-being () |
| improve my overall well-being () |
| improve my loved one's quality of life () |
| improve my quality of life () |
| learn more T1D tips, tricks, and/or coping strategies () |
| live more easily with T1D () |
| make friends () |
| manage stress better () |
| reduce the burden T1D puts on me () |
| sleep better () |

End of Block: CommuniT1D check-in measure

---

Start of Block: Caregiver check-in

*Display this question:*

*If dlovedone = 1*

In the past 6 months, have you done any of the following things to help your loved one manage their diabetes? (check all that apply)

- ☐ Brought them something when their blood sugar is low (1)
- ☐ Called 911 for them (2)
- ☐ Changed plans to accommodate diabetes-related needs (3)
- ☐ Changed a pump site for them or helped them change a pump site (4)
- ☐ Checked their blood sugar for them or help them check their blood sugar (5)
- ☐ Checked their blood sugar for them while they were sleeping (6)
- ☐ Counted carbs for them or helped them count carbs (7)
- ☐ Figured out their insulin doses or helped them figure out their doses (8)
- ☐ Filled out and/or submitted diabetes-related paperwork (e.g., private or government insurance paperwork) or helped them fill out and/or submit paperwork (9)
- ☐ Gave them insulin (injected insulin, used their insulin pump to give them insulin) (10)
- ☐ Helped them build a do-it-yourself system of some kind or built a system for them (e.g., xDrip, Nightscout site, AAPS, Loop, iAPS, etc.) (11)
- ☐ Helped them deal with high blood sugar while awake (12)
- ☐ Helped them deal with low blood sugar while awake (13)
- ☐ Helped them deal with high blood sugar while sleeping (14)
- ☐ Helped them deal with low blood sugar while sleeping (15)

☐ Inserted a continuous glucose monitoring (CGM) sensor or helped them insert a sensor (16)

☐ Made them meals or snacks (17)

☐ Ordered or picked up medications or supplies (at the pharmacy or from a supplier like Diabetes Express) (18)

☐ Paid for diabetes medications or supplies in whole or in part (19)

☐ Reminded them about diabetes-related tasks or goals (20)

☐ Supported them emotionally about their diabetes (21)

☐ Took them to medical appointments (22)

☐ Went to their school, work, or other location to give them insulin, bring them something they needed to manage diabetes (e.g., a blood test meter or pump supplies), or for another diabetes-related reason (23)

☐ Other (24) ________________________________________________

☐ Other (25) ________________________________________________

☐ Other (26) ________________________________________________

**End of Block: Caregiver check-in**

---

**Start of Block: Health care & access**

*Display this question:*

*If Do you yourself have type 1 diabetes (T1D) or a similar type of diabetes, such as LADA, MODY, or... = Yes*

Do you have one or more health professionals (doctors, nurses, dietitians, pharmacists, etc.) who provide regular T1D care for you? By “regular” we mean at least once a year. (choose one)

- Yes (1)
- No (2)
- I don’t know (3)
- I prefer not to answer (4)

---

*Display this question:*

*If Do you have one or more health professionals (doctors, nurses, dietitians, pharmacists, etc.) who... = Yes*

What kind(s) of health professionals provide regular (at least once a year) diabetes care for you? (check all that apply)

- ☐ Diabetes educator (also known as certified diabetes educator/CDE) (13)
- ☐ Diabetologist (14)
- ☐ Dietitian (15)
- ☐ Doctor (but I don't know what kind) (16)
- ☐ Endocrinologist (17)
- ☐ Family doctor (18)
- ☐ Internal medicine doctor (19)
- ☐ Mental health specialist (20)
- ☐ Nurse (21)
- ☐ Pharmacist (22)
- ☐ I don't know (23)
- ☐ I prefer not to answer (24)

---

*Display this question:*

*If Do you yourself have type 1 diabetes (T1D) or a similar type of diabetes, such as LADA, MODY, or... = Yes*

Do you have a specific goal for your hemoglobin A1c (the blood test that many people with diabetes get every 3-6 months)? (choose one)

- Yes (1)
- No (2)
- I don't know (3)
- I prefer not to answer (4)

*Display this question:*

*If Do you have a specific goal for your hemoglobin A1c (the blood test that many people with diabete... = Yes*

What is your goal for your hemoglobin A1c? (choose one) *In this question, we present A1c in %, the common presentation in Canada. If you use mmol/mol (the current international standard), here are the equivalent numbers: 4.0% = 20 mmol/mol; 5.6% = 38 mmol/mol; 6.0% =*

*42 mmol/mol; 6.1% = 43 mmol/mol; 6.5% = 48 mmol/mol; 7.0% = 53 mmol/mol; 7.5% = 58 mmol/mol; 8.0% = 64 mmol/mol; 8.5% = 69 mmol/mol*

○ 4.0% to 5.6% (the nondiabetic range) (1)

○ 6.0% or lower (2)

○ 6.1% or lower (the recommended goal in Canada when pregnant with T1D) (3)

○ 6.5% or lower (the recommended goal in Canada when planning a pregnancy with T1D) (4)

○ 7.0% or lower (the recommended goal in Canada for most adults with T1D) (5)

○ 7.5% or lower (the recommended goal in Canada for most children and adolescents under 18 years old with T1D) (6)

○ 8.0% or lower (the recommended goal in Canada for some people who are older or for other reasons) (7)

○ 8.5% or lower (the recommended goal in Canada for some people who are older, who have severe hypo unawareness, or for other reasons) (8)

○ other (9) ________________________________________________

○ I prefer not to answer (10)

*Display this question:*

*If What is your goal for your hemoglobin A1c? (choose one) In this question, we present A1c in %, th... = 4.0% to 5.6% (the nondiabetic range)*

*Or What is your goal for your hemoglobin A1c? (choose one) In this question, we present A1c in %, th... = 6.0% or lower*

*Or What is your goal for your hemoglobin A1c? (choose one) In this question, we present A1c in %, th... = 6.1% or lower (the recommended goal in Canada when pregnant with T1D)*

*Or What is your goal for your hemoglobin A1c? (choose one) In this question, we present A1c in %, th... = 6.5% or lower (the recommended goal in Canada when planning a pregnancy with T1D)*

*Or What is your goal for your hemoglobin A1c? (choose one) In this question, we present A1c in %, th... = 7.0% or lower (the recommended goal in Canada for most adults with T1D)*

*Or What is your goal for your hemoglobin A1c? (choose one) In this question, we present A1c in %, th... = 7.5% or lower (the recommended goal in Canada for most children and adolescents under 18 years old with T1D)*

*Or What is your goal for your hemoglobin A1c? (choose one) In this question, we present A1c in %, th... = 8.0% or lower (the recommended goal in Canada for some people who are older or for other reasons)*

*Or What is your goal for your hemoglobin A1c? (choose one) In this question, we present A1c in %, th... = 8.5% or lower (the recommended goal in Canada for some people who are older, who have severe hypo unawareness, or for other reasons)*

*Or Or What is your goal for your hemoglobin A1c? (choose one)In this question, we present A1c in %, the common presentation in Canada. If you use mmol/mol (the current international standard), here are t... Text Response Is Not Empty*

Who set this goal? (choose one)

○ My health care provider (endocrinologist, other doctor, diabetes educator, etc.) set the goal. (7)

○ My health care provider and I set the goal together. (8)

○ I set the goal for myself. (9)

○ Other (10) ________________________________________________

○ I don’t know (11)

○ I prefer not to answer (12)

---

*Display this question:*

*If Do you yourself have type 1 diabetes (T1D) or a similar type of diabetes, such as LADA, MODY, or... = Yes*

Do you use a continuous glucose monitor or flash glucose monitor (e.g., Dexcom, Libre, Medtronic Guardian sensor)?

◯ Yes (1)

◯ No (2)

◯ I don't know (3)

◯ I prefer not to answer (4)

---

*Display this question:*

*If Do you use a continuous glucose monitor or flash glucose monitor (e.g., Dexcom, Libre, Medtronic... = Yes*

Do you have a specific time in range goal? (choose one)

◯ Yes (1)

◯ No (2)

◯ I don't know (3)

◯ I prefer not to answer (4)

---

*Display this question:*

*If Do you have a specific time in range goal? (choose one) = Yes*

What is the lower limit of the target range for your time in range goal? (choose one) *In this question, we present blood glucose values in mmol/L, the common presentation in Canada. If you use mg/dl, here are some equivalent numbers: 70 mg/dl = 3.9 mmol/L; 80 mg/dl = 4.4 mmol/L; 90 mg/dl = 5.0 mmol/L; 100 mg/dl = 5.6 mmol/L; 110 mg/dl = 6.1 mmol/L; 120 mg/dl = 6.7 mmol/L; 130 mg/dl = 7.2 mmol/L; 140 mg/dl = 7.8 mmol/L; 150 mg/dl = 8.3 mmol/L; 160*

*mg/dl = 8.9 mmol/L; 170 mg/dl = 9.4 mmol/L; 180 mg/dl = 10.0 mmol/L; 190 mg/dl = 10.6 mmol/L; 200 mg/dl = 11.1 mmol/L*

- ○ below 3.6 mmol/L (5)
- ○ 3.6 mmol/L (105)
- ○ 3.7 mmol/L (106)
- ○ 3.8 mmol/L (107)
- ○ 3.9 mmol/L (consensus value used in Ambulatory Glucose Profile reports) (108)
- ○ 4.0 mmol/L (109)
- ○ 4.1 mmol/L (110)
- ○ 4.2 mmol/L (111)
- ○ 4.3 mmol/L (112)
- ○ 4.4 mmol/L (113)
- ○ 4.5 mmol/L (114)
- ○ 4.6 mmol/L (115)
- ○ 4.7 mmol/L (116)
- ○ 4.8 mmol/L (117)
- ○ 4.9 mmol/L (118)
- ○ 5.0 mmol/L (119)
- ○ 5.1 mmol/L (120)
- ○ 5.2 mmol/L (121)
- ○ 5.3 mmol/L (122)
- ○ 5.4 mmol/L (123)

○ 5.5 mmol/L (124)

○ 5.6 mmol/L (125)

○ 5.7 mmol/L (126)

○ 5.8 mmol/L (127)

○ 5.9 mmol/L (128)

○ 6.0 mmol/L (129)

○ 6.1 mmol/L (130)

○ 6.2 mmol/L (131)

○ 6.3 mmol/L (132)

○ 6.4 mmol/L (133)

○ 6.5 mmol/L (134)

○ 6.6 mmol/L (135)

○ 6.7 mmol/L (136)

○ 6.8 mmol/L (137)

○ 6.9 mmol/L (138)

○ 7.0 mmol/L (139)

○ above 7.0 mmol/L (140)

○ I don't know (141)

○ I prefer not to answer (142)

---

*Display this question:*

*If Do you have a specific time in range goal? (choose one) = Yes*

What is the upper limit of the target range for your time in range goal? (choose one) *In this question, we present blood glucose values in mmol/L, the common presentation in Canada. If you use mg/dl, here are some equivalent numbers: 70 mg/dl = 3.9 mmol/L; 80 mg/dl = 4.4 mmol/L; 90 mg/dl = 5.0 mmol/L; 100 mg/dl = 5.6 mmol/L; 110 mg/dl = 6.1 mmol/L; 120 mg/dl = 6.7 mmol/L; 130 mg/dl = 7.2 mmol/L; 140 mg/dl = 7.8 mmol/L; 150 mg/dl = 8.3 mmol/L; 160*

*mg/dl = 8.9 mmol/L; 170 mg/dl = 9.4 mmol/L; 180 mg/dl = 10.0 mmol/L; 190 mg/dl = 10.6 mmol/L; 200 mg/dl = 11.1 mmol/L*

○ under 5.0 mmol/L (5)

○ 5.0 mmol/L (143)

○ 5.1 mmol/L (144)

○ 5.2 mmol/L (145)

○ 5.3 mmol/L (146)

○ 5.4 mmol/L (147)

○ 5.5 mmol/L (148)

○ 5.6 mmol/L (149)

○ 5.7 mmol/L (150)

○ 5.8 mmol/L (151)

○ 5.9 mmol/L (152)

○ 6.0 mmol/L (153)

○ 6.1 mmol/L (154)

○ 6.2 mmol/L (155)

○ 6.3 mmol/L (156)

○ 6.4 mmol/L (157)

○ 6.5 mmol/L (158)

○ 6.6 mmol/L (159)

○ 6.7 mmol/L (160)

○ 6.8 mmol/L (161)

○ 6.9 mmol/L (162)

○ 7.0 mmol/L (163)

○ 7.1 mmol/L (164)

○ 7.2 mmol/L (165)

○ 7.3 mmol/L (166)

○ 7.4 mmol/L (167)

○ 7.5 mmol/L (168)

○ 7.6 mmol/L (169)

○ 7.7 mmol/L (170)

○ 7.8 mmol/L (171)

○ 7.9 mmol/L (172)

○ 8.0 mmol/L (173)

○ 8.1 mmol/L (174)

○ 8.2 mmol/L (175)

○ 8.3 mmol/L (176)

○ 8.4 mmol/L (177)

○ 8.5 mmol/L (178)

○ 8.6 mmol/L (179)

○ 8.7 mmol/L (180)

○ 8.8 mmol/L (181)

○ 8.9 mmol/L (182)

- 9.0 mmol/L  (183)
- 9.1 mmol/L  (184)
- 9.2 mmol/L  (185)
- 9.3 mmol/L  (186)
- 9.4 mmol/L  (187)
- 9.5 mmol/L  (188)
- 9.6 mmol/L  (189)
- 9.7 mmol/L  (190)
- 9.8 mmol/L  (191)
- 9.9 mmol/L  (192)
- 10.0 mmol/L (consensus value used in Ambulatory Glucose Profile reports)  (193)
- 10.1 mmol/L  (194)
- 10.2 mmol/L  (195)
- 10.3 mmol/L  (196)
- 10.4 mmol/L  (197)
- 10.5 mmol/L  (198)
- 10.6 mmol/L  (199)
- 10.7 mmol/L  (200)
- 10.8 mmol/L  (201)
- 10.9 mmol/L  (202)
- 11.0 mmol/L  (203)

- 11.1 mmol/L (204)
- 11.2 mmol/L (205)
- 11.3 mmol/L (206)
- 11.4 mmol/L (207)
- 11.5 mmol/L (208)
- 11.6 mmol/L (209)
- 11.7 mmol/L (210)
- 11.8 mmol/L (211)
- 11.9 mmol/L (212)
- 12.0 mmol/L (213)
- 12.1 mmol/L (214)
- 12.2 mmol/L (215)
- 12.3 mmol/L (216)
- 12.4 mmol/L (217)
- 12.5 mmol/L (218)
- 12.6 mmol/L (219)
- 12.7 mmol/L (220)
- 12.8 mmol/L (221)
- 12.9 mmol/L (222)
- 13.0 mmol/L (223)
- 13.1 mmol/L (224)

- 13.2 mmol/L (225)
- 13.3 mmol/L (226)
- 13.4 mmol/L (227)
- 13.5 mmol/L (228)
- 13.6 mmol/L (229)
- 13.7 mmol/L (230)
- 13.8 mmol/L (231)
- 13.9 mmol/L (232)
- 14.0 mmol/L (233)
- 14.1 mmol/L (234)
- 14.2 mmol/L (235)
- 14.3 mmol/L (236)
- 14.4 mmol/L (237)
- 14.5 mmol/L (238)
- 14.6 mmol/L (239)
- 14.7 mmol/L (240)
- 14.8 mmol/L (241)
- 14.9 mmol/L (242)
- 15.0 mmol/L (243)
- above 15.0 mmol/L (244)
- I don’t know (245)

○ I prefer not to answer (246)

*Display this question:*

*If Do you have a specific time in range goal? (choose one) = Yes*

Comments about your target range (optional)

________________________________________________________________

________________________________________________________________

________________________________________________________________

________________________________________________________________

________________________________________________________________

*Display this question:*

*If Do you have a specific time in range goal? (choose one) = Yes*

What is your goal for the percentage of time you spend in your target range? My goal is to spend at least this percent of time in my target range:

- ○ 0% (4)
- ○ 1% (5)
- ○ 2% (6)
- ○ 3% (7)
- ○ 4% (8)
- ○ 5% (9)
- ○ 6% (10)
- ○ 7% (11)
- ○ 8% (12)
- ○ 9% (13)
- ○ 10% (14)
- ○ 11% (15)
- ○ 12% (16)
- ○ 13% (17)
- ○ 14% (18)
- ○ 15% (19)
- ○ 16% (20)
- ○ 17% (21)
- ○ 18% (22)
- ○ 19% (23)

○ 20% (24)

○ 21% (25)

○ 22% (26)

○ 23% (27)

○ 24% (28)

○ 25% (29)

○ 26% (30)

○ 27% (31)

○ 28% (32)

○ 29% (33)

○ 30% (34)

○ 31% (35)

○ 32% (36)

○ 33% (37)

○ 34% (38)

○ 35% (39)

○ 36% (40)

○ 37% (41)

○ 38% (42)

○ 39% (43)

○ 40% (44)

- 41% (45)
- 42% (46)
- 43% (47)
- 44% (48)
- 45% (49)
- 46% (50)
- 47% (51)
- 48% (52)
- 49% (53)
- 50% (54)
- 51% (55)
- 52% (56)
- 53% (57)
- 54% (58)
- 55% (59)
- 56% (60)
- 57% (61)
- 58% (62)
- 59% (63)
- 60% (64)
- 61% (65)

- 62%  (66)
- 63%  (67)
- 64%  (68)
- 65%  (69)
- 66%  (70)
- 67%  (71)
- 68%  (72)
- 69%  (73)
- 70% (consensus value used in Ambulatory Glucose Profile reports)  (74)
- 71%  (75)
- 72%  (76)
- 73%  (77)
- 74%  (78)
- 75%  (79)
- 76%  (80)
- 77%  (81)
- 78%  (82)
- 79%  (83)
- 80%  (84)
- 81%  (85)
- 82%  (86)

○ 83% (87)

○ 84% (88)

○ 85% (89)

○ 86% (90)

○ 87% (91)

○ 88% (92)

○ 89% (93)

○ 90% (94)

○ 91% (95)

○ 92% (96)

○ 93% (97)

○ 94% (98)

○ 95% (99)

○ 96% (100)

○ 97% (101)

○ 98% (102)

○ 99% (103)

○ 100% (104)

○ I don’t know (105)

○ I prefer not to answer (106)

*Display this question:*

*If Do you have a specific time in range goal? (choose one) = Yes*

Who set this goal? (choose one)

- ◯ My health care provider (endocrinologist, other doctor, diabetes educator, etc.) set the goal. (7)
- ◯ My health care provider and I set the goal together. (8)
- ◯ I set the goal for myself. (9)
- ◯ Other (10)
- ◯ I don’t know (11)
- ◯ I prefer not to answer (12)

---

*Display this question:*

*If Do you yourself have type 1 diabetes (T1D) or a similar type of diabetes, such as LADA, MODY, or... = Yes*

Have you had a hemoglobin A1c test within the past 6 months? (choose one)

- ◯ Yes (1)
- ◯ No (2)
- ◯ I don’t know (3)
- ◯ I prefer not to answer (4)

---

*Display this question:*

*If Have you had a hemoglobin A1c test within the past 6 months? (choose one) = Yes*

What was your most recent hemoglobin A1c result?

- under 4.0% (under 20 mmol/mol) (346)
- 4.0% (20 mmol/mol) (347)
- 4.1% (21 mmol/mol) (348)
- 4.2% (22 mmol/mol) (349)
- 4.3% (23 mmol/mol) (350)
- 4.4% (25 mmol/mol) (351)
- 4.5% (26 mmol/mol) (352)
- 4.6% (27 mmol/mol) (353)
- 4.7% (28 mmol/mol) (354)
- 4.8% (29 mmol/mol) (355)
- 4.9% (30 mmol/mol) (356)
- 5.0% (31 mmol/mol) (357)
- 5.1% (32 mmol/mol) (358)
- 5.2% (33 mmol/mol) (359)
- 5.3% (34 mmol/mol) (360)
- 5.4% (36 mmol/mol) (361)
- 5.5% (37 mmol/mol) (362)
- 5.6% (38 mmol/mol) (363)
- 5.7% (39 mmol/mol) (364)
- 5.8% (40 mmol/mol) (365)
- 5.9% (41 mmol/mol) (366)

○ 6.0% (42 mmol/mol) (367)

○ 6.1% (43 mmol/mol) (368)

○ 6.2% (44 mmol/mol) (369)

○ 6.3% (45 mmol/mol) (370)

○ 6.4% (46 mmol/mol) (371)

○ 6.5% (48 mmol/mol) (372)

○ 6.6% (49 mmol/mol) (373)

○ 6.7% (50 mmol/mol) (374)

○ 6.8% (51 mmol/mol) (375)

○ 6.9% (52 mmol/mol) (376)

○ 7.0% (53 mmol/mol) (377)

○ 7.1% (54 mmol/mol) (378)

○ 7.2% (55 mmol/mol) (379)

○ 7.3% (56 mmol/mol) (380)

○ 7.4% (57 mmol/mol) (381)

○ 7.5% (58 mmol/mol) (382)

○ 7.6% (59 mmol/mol) (383)

○ 7.7% (60 mmol/mol) (384)

○ 7.8% (62 mmol/mol) (385)

○ 7.9% (63 mmol/mol) (386)

○ 8.0% (64 mmol/mol) (387)

○ 8.1% (65 mmol/mol) (388)

○ 8.2% (66 mmol/mol) (389)

○ 8.3% (67 mmol/mol) (390)

○ 8.4% (68 mmol/mol) (391)

○ 8.5% (69 mmol/mol) (392)

○ 8.6% (70 mmol/mol) (393)

○ 8.7% (72 mmol/mol) (394)

○ 8.8% (73 mmol/mol) (395)

○ 8.9% (74 mmol/mol) (396)

○ 9.0% (75 mmol/mol) (397)

○ 9.1% (76 mmol/mol) (398)

○ 9.2% (77 mmol/mol) (399)

○ 9.3% (78 mmol/mol) (400)

○ 9.4% (79 mmol/mol) (401)

○ 9.5% (80 mmol/mol) (402)

○ 9.6% (81 mmol/mol) (403)

○ 9.7% (83 mmol/mol) (404)

○ 9.8% (84 mmol/mol) (405)

○ 9.9% (85 mmol/mol) (406)

○ 10.0% (86 mmol/mol) (407)

○ 10.1% (87 mmol/mol) (408)

- 10.2% (88 mmol/mol) (409)
- 10.3% (89 mmol/mol) (410)
- 10.4% (90 mmol/mol) (411)
- 10.5% (91 mmol/mol) (412)
- 10.6% (92 mmol/mol) (413)
- 10.7% (93 mmol/mol) (414)
- 10.8% (94 mmol/mol) (415)
- 10.9% (95 mmol/mol) (416)
- 11.0% (97 mmol/mol) (417)
- 11.1% (98 mmol/mol) (418)
- 11.2% (99 mmol/mol) (419)
- 11.3% (100 mmol/mol) (420)
- 11.4% (101 mmol/mol) (421)
- 11.5% (102 mmol/mol) (422)
- 11.6% (103 mmol/mol) (423)
- 11.7% (104 mmol/mol) (424)
- 11.8% (105 mmol/mol) (425)
- 11.9% (106 mmol/mol) (426)
- 12.0% (108 mmol/mol) (427)
- 12.1% (109 mmol/mol) (428)
- 12.2% (110 mmol/mol) (429)

○ 12.3% (111 mmol/mol) (430)

○ 12.4% (112 mmol/mol) (431)

○ 12.5% (113 mmol/mol) (432)

○ 12.6% (114 mmol/mol) (433)

○ 12.7% (115 mmol/mol) (434)

○ 12.8% (116 mmol/mol) (435)

○ 12.9% (117 mmol/mol) (436)

○ 13.0% (119 mmol/mol) (437)

○ 13.1% (120 mmol/mol) (438)

○ 13.2% (121 mmol/mol) (439)

○ 13.3% (122 mmol/mol) (440)

○ 13.4% (123 mmol/mol) (441)

○ 13.5% (124 mmol/mol) (442)

○ 13.6% (125 mmol/mol) (443)

○ 13.7% (126 mmol/mol) (444)

○ 13.8% (127 mmol/mol) (445)

○ 13.9% (128 mmol/mol) (446)

○ 14.0% (130 mmol/mol) (447)

○ 14.1% (131 mmol/mol) (448)

○ 14.2% (132 mmol/mol) (449)

○ 14.3% (133 mmol/mol) (450)

- ○ 14.4% (134 mmol/mol) (451)
- ○ 14.5% (135 mmol/mol) (452)
- ○ 14.6% (136 mmol/mol) (453)
- ○ 14.7% (137 mmol/mol) (454)
- ○ 14.8% (138 mmol/mol) (455)
- ○ 14.9% (139 mmol/mol) (456)
- ○ 15.0% (140 mmol/mol) (457)
- ○ Above 15.0% (above 140 mmol/mol) (458)
- ○ I don't know (459)
- ○ I prefer not to answer (460)

---

*Display this question:*

*If Have you had a hemoglobin A1c test within the past 6 months? (choose one) = Yes*

When was this hemoglobin A1c test done?

- This month (1)
- Last month (2)
- 2 months ago (3)
- 3 months ago (4)
- 4 months ago (5)
- 5 months ago (6)
- 6 months ago (7)
- More than 6 months ago (possible if they had a test more recently but don’t know the result) (8)
- I don’t know (9)
- I prefer not to answer (10)

---

*Display this question:*

*If Do you use a continuous glucose monitor or flash glucose monitor (e.g., Dexcom, Libre, Medtronic... = Yes*

Do you know how to check your Time in Range in your CGM app? (choose one)

- Yes (1)
- No (2)
- I don’t know (3)
- I prefer not to answer (4)

---

*Display this question:*

*If Do you know how to check your Time in Range in your CGM app? (choose one) = Yes*

Over the past 90 days, what was your Time in Range for the standard range 3.9 mmol/L - 10 mmol/L?

- ○ 0% (1)
- ○ 1% (2)
- ○ 2% (3)
- ○ 3% (4)
- ○ 4% (5)
- ○ 5% (6)
- ○ 6% (7)
- ○ 7% (8)
- ○ 8% (9)
- ○ 9% (10)
- ○ 10% (11)
- ○ 11% (12)
- ○ 12% (13)
- ○ 13% (14)
- ○ 14% (15)
- ○ 15% (16)
- ○ 16% (17)
- ○ 17% (18)
- ○ 18% (19)
- ○ 19% (20)

- 20% (21)
- 21% (22)
- 22% (23)
- 23% (24)
- 24% (25)
- 25% (26)
- 26% (27)
- 27% (28)
- 28% (29)
- 29% (30)
- 30% (31)
- 31% (32)
- 32% (33)
- 33% (34)
- 34% (35)
- 35% (36)
- 36% (37)
- 37% (38)
- 38% (39)
- 39% (40)
- 40% (41)

- 41% (42)
- 42% (43)
- 43% (44)
- 44% (45)
- 45% (46)
- 46% (47)
- 47% (48)
- 48% (49)
- 49% (50)
- 50% (51)
- 51% (52)
- 52% (53)
- 53% (54)
- 54% (55)
- 55% (56)
- 56% (57)
- 57% (58)
- 58% (59)
- 59% (60)
- 60% (61)
- 61% (62)

○ 62% (63)

○ 63% (64)

○ 64% (65)

○ 65% (66)

○ 66% (67)

○ 67% (68)

○ 68% (69)

○ 69% (70)

○ 70% (71)

○ 71% (72)

○ 72% (73)

○ 73% (74)

○ 74% (75)

○ 75% (76)

○ 76% (77)

○ 77% (78)

○ 78% (79)

○ 79% (80)

○ 80% (81)

○ 81% (82)

○ 82% (83)

- 83%  (84)
- 84%  (85)
- 85%  (86)
- 86%  (87)
- 87%  (88)
- 88%  (89)
- 89%  (90)
- 90%  (91)
- 91%  (92)
- 92%  (93)
- 93%  (94)
- 94%  (95)
- 95%  (96)
- 96%  (97)
- 97%  (98)
- 98%  (99)
- 99%  (100)
- 100%  (101)
- I don’t know    (102)
- I prefer not to answer  (103)

*Display this question:*

*If Do you yourself have type 1 diabetes (T1D) or a similar type of diabetes, such as LADA, MODY, or... = Yes*

Over the past 6 months, have you had any **severe** hypoglycemic (low blood sugar) events in which you required help from someone else? (choose one)

- Yes (1)
- No (2)
- I don't know (3)
- I prefer not to answer (4)

*Display this question:*

*If Over the past 6 months, have you had any severe hypoglycemic (low blood sugar) events in which yo... = Yes*

When you required help from someone else for the severe hypoglycemic (low blood sugar) event(s), did that help include: (check all that apply)

- Emergency services (e.g., someone called 911) (1)
- Glucagon (e.g., someone gave you glucagon by needle or by nose) (2)
- Hospitalization (you received treatment at a hospital) (3)
- Someone getting low treatment (e.g., juice) and bringing it to you (4)
- I don't know (5)
- I prefer not to answer (6)

*Display this question:*

*If How many children under age 18 with type 1 diabetes (T1D) or a similar type of diabetes (such as... = 2*

We are now going to ask you some more questions about your 2 children under age 18 with T1D (or a similar type of diabetes). Please answer about your older child first, then your younger child.

*Display this question:*

*If How many children under age 18 with type 1 diabetes (T1D) or a similar type of diabetes (such as... = 3*

We are now going to ask you some more questions about your 3 children under age 18 with T1D (or a similar type of diabetes). Please answer about your eldest child first, then your next-eldest, and finally, your youngest child.

*Display this question:*

*If How many children under age 18 with type 1 diabetes (T1D) or a similar type of diabetes (such as... = More than 3*

You indicated that you have more than 3 children under age 18 with T1D (or a similar type of diabetes). This survey is designed for up to 3 children. For the following set of questions, please answer about your three children who have had T1D the longest. Please answer about the eldest child first, then the next-eldest, and finally, the youngest child.

*Display this question:*

*If Are you the parent or guardian of one or more children under age 18 with type 1 diabetes (T1D) or... = Yes*

Does your child have one or more health professionals (doctors, nurses, dietitians, pharmacists, etc.) who provide regular T1D care for them? By “regular” we mean at least once a year. (choose one)

- Yes (1)
- No (2)
- I don’t know (3)
- I prefer not to answer (4)

---

*Display this question:*

*If Does your child have one or more health professionals (doctors, nurses, dietitians, pharmacists,... = Yes*

What kind(s) of health professionals provide regular (at least once a year) diabetes care for your child? (check all that apply)

- ▢ Diabetes educator (also known as certified diabetes educator/CDE) (13)
- ▢ Diabetologist (14)
- ▢ Dietitian (15)
- ▢ Doctor (but I don't know what kind) (16)
- ▢ Endocrinologist (17)
- ▢ Family doctor (18)
- ▢ Internal medicine doctor (19)
- ▢ Mental health specialist (20)
- ▢ Nurse (21)
- ▢ Pharmacist (22)
- ▢ I don't know (23)
- ▢ I prefer not to answer (24)

---

*Display this question:*

*If Are you the parent or guardian of one or more children under age 18 with type 1 diabetes (T1D) or... = Yes*

Does your child have a specific goal for their hemoglobin A1c (the blood test that many people with diabetes get every 3-6 months)? (choose one)

- Yes (1)
- No (2)
- I don’t know (3)
- I prefer not to answer (4)

---

*Display this question:*

*If Does your child have a specific goal for their hemoglobin A1c (the blood test that many people wi... = Yes*

What is their goal for their hemoglobin A1c? (choose one) *In this question, we present A1c in %, the common presentation in Canada. If you use mmol/mol (the current international standard), here are the equivalent numbers: 4.0% = 20 mmol/mol; 5.6% = 38 mmol/mol; 6.0% =*

*42 mmol/mol; 6.1% = 43 mmol/mol; 6.5% = 48 mmol/mol; 7.0% = 53 mmol/mol; 7.5% = 58 mmol/mol; 8.0% = 64 mmol/mol; 8.5% = 69 mmol/mol*

○ 4.0% to 5.6% (the nondiabetic range) (1)

○ 6.0% or lower (2)

○ 6.1% or lower (the recommended goal in Canada when pregnant with T1D) (3)

○ 6.5% or lower (the recommended goal in Canada when planning a pregnancy with T1D) (4)

○ 7.0% or lower (the recommended goal in Canada for most adults with T1D) (5)

○ 7.5% or lower (the recommended goal in Canada for most children and adolescents under 18 years old with T1D) (6)

○ 8.0% or lower (the recommended goal in Canada for some people who are older or for other reasons) (7)

○ 8.5% or lower (the recommended goal in Canada for some people who are older, who have severe hypo unawareness, or for other reasons) (8)

○ other (9) ________________________________________________

○ I prefer not to answer (10)

*Display this question:*

*If What is their goal for their hemoglobin A1c? (choose one) In this question, we present A1c in %,... = 4.0% to 5.6% (the nondiabetic range)*

*Or What is their goal for their hemoglobin A1c? (choose one) In this question, we present A1c in %,... = 6.0% or lower*

*Or What is their goal for their hemoglobin A1c? (choose one) In this question, we present A1c in %,... = 6.1% or lower (the recommended goal in Canada when pregnant with T1D)*

*Or What is their goal for their hemoglobin A1c? (choose one) In this question, we present A1c in %,... = 6.5% or lower (the recommended goal in Canada when planning a pregnancy with T1D)*

*Or What is their goal for their hemoglobin A1c? (choose one) In this question, we present A1c in %,... = 7.0% or lower (the recommended goal in Canada for most adults with T1D)*

*Or What is their goal for their hemoglobin A1c? (choose one) In this question, we present A1c in %,... = 7.5% or lower (the recommended goal in Canada for most children and adolescents under 18 years old with T1D)*

*Or What is their goal for their hemoglobin A1c? (choose one) In this question, we present A1c in %,... = 8.0% or lower (the recommended goal in Canada for some people who are older or for other reasons)*

*Or What is their goal for their hemoglobin A1c? (choose one) In this question, we present A1c in %,... = 8.5% or lower (the recommended goal in Canada for some people who are older, who have severe hypo unawareness, or for other reasons)*

*Or Or What is their goal for their hemoglobin A1c? (choose one) Text Response Is Not Empty*

Who set this goal? (choose one)

- ◯ My child's health care provider (endocrinologist, other doctor, diabetes educator, etc.) set the goal. (7)
- ◯ My child's health care provider and our family set the goal together. (8)
- ◯ Our family set the goal. (9)
- ◯ Other (10) ________________________________________________
- ◯ I don't know (11)
- ◯ I prefer not to answer (12)

*Display this question:*

*If Are you the parent or guardian of one or more children under age 18 with type 1 diabetes (T1D) or... = Yes*

Does your child use a continuous glucose monitor or flash glucose monitor (e.g., Dexcom, Libre, Medtronic Guardian sensor)?

- ◯ Yes (1)
- ◯ No (2)
- ◯ I don't know (3)
- ◯ I prefer not to answer (4)

---

*Display this question:*

*If Does your child use a continuous glucose monitor or flash glucose monitor (e.g., Dexcom, Libre, M... = Yes*

Does your child have a specific time in range goal? (choose one)

- ◯ Yes (1)
- ◯ No (2)
- ◯ I don't know (3)
- ◯ I prefer not to answer (4)

---

*Display this question:*

*If Does your child have a specific time in range goal? (choose one) = Yes*

What is the lower limit of the target range for their time in range goal? (choose one) *In this question, we present blood glucose values in mmol/L, the common presentation in Canada. If you use mg/dl, here are some equivalent numbers: 70 mg/dl = 3.9 mmol/L; 80 mg/dl = 4.4 mmol/L; 90 mg/dl = 5.0 mmol/L; 100 mg/dl = 5.6 mmol/L; 110 mg/dl = 6.1 mmol/L; 120 mg/dl = 6.7 mmol/L; 130 mg/dl = 7.2 mmol/L; 140 mg/dl = 7.8 mmol/L; 150 mg/dl = 8.3 mmol/L; 160*

*mg/dl = 8.9 mmol/L; 170 mg/dl = 9.4 mmol/L; 180 mg/dl = 10.0 mmol/L; 190 mg/dl = 10.6 mmol/L; 200 mg/dl = 11.1 mmol/L*

- ◯ below 3.6 mmol/L (5)
- ◯ 3.6 mmol/L (105)
- ◯ 3.7 mmol/L (106)
- ◯ 3.8 mmol/L (107)
- ◯ 3.9 mmol/L (consensus value used in Ambulatory Glucose Profile reports) (108)
- ◯ 4.0 mmol/L (109)
- ◯ 4.1 mmol/L (110)
- ◯ 4.2 mmol/L (111)
- ◯ 4.3 mmol/L (112)
- ◯ 4.4 mmol/L (113)
- ◯ 4.5 mmol/L (114)
- ◯ 4.6 mmol/L (115)
- ◯ 4.7 mmol/L (116)
- ◯ 4.8 mmol/L (117)
- ◯ 4.9 mmol/L (118)
- ◯ 5.0 mmol/L (119)
- ◯ 5.1 mmol/L (120)
- ◯ 5.2 mmol/L (121)
- ◯ 5.3 mmol/L (122)
- ◯ 5.4 mmol/L (123)

○ 5.5 mmol/L (124)

○ 5.6 mmol/L (125)

○ 5.7 mmol/L (126)

○ 5.8 mmol/L (127)

○ 5.9 mmol/L (128)

○ 6.0 mmol/L (129)

○ 6.1 mmol/L (130)

○ 6.2 mmol/L (131)

○ 6.3 mmol/L (132)

○ 6.4 mmol/L (133)

○ 6.5 mmol/L (134)

○ 6.6 mmol/L (135)

○ 6.7 mmol/L (136)

○ 6.8 mmol/L (137)

○ 6.9 mmol/L (138)

○ 7.0 mmol/L (139)

○ above 7.0 mmol/L (140)

○ I don't know (141)

○ I prefer not to answer (142)

---

*Display this question:*

*If Does your child have a specific time in range goal? (choose one) = Yes*

What is the upper limit of the target range for their time in range goal? (choose one) *In this question, we present blood glucose values in mmol/L, the common presentation in Canada. If you use mg/dl, here are some equivalent numbers: 70 mg/dl = 3.9 mmol/L; 80 mg/dl = 4.4 mmol/L; 90 mg/dl = 5.0 mmol/L; 100 mg/dl = 5.6 mmol/L; 110 mg/dl = 6.1 mmol/L; 120 mg/dl = 6.7 mmol/L; 130 mg/dl = 7.2 mmol/L; 140 mg/dl = 7.8 mmol/L; 150 mg/dl = 8.3 mmol/L; 160*

*mg/dl = 8.9 mmol/L; 170 mg/dl = 9.4 mmol/L; 180 mg/dl = 10.0 mmol/L; 190 mg/dl = 10.6 mmol/L; 200 mg/dl = 11.1 mmol/L*

- under 5.0 mmol/L (5)
- 5.0 mmol/L (143)
- 5.1 mmol/L (144)
- 5.2 mmol/L (145)
- 5.3 mmol/L (146)
- 5.4 mmol/L (147)
- 5.5 mmol/L (148)
- 5.6 mmol/L (149)
- 5.7 mmol/L (150)
- 5.8 mmol/L (151)
- 5.9 mmol/L (152)
- 6.0 mmol/L (153)
- 6.1 mmol/L (154)
- 6.2 mmol/L (155)
- 6.3 mmol/L (156)
- 6.4 mmol/L (157)
- 6.5 mmol/L (158)
- 6.6 mmol/L (159)
- 6.7 mmol/L (160)
- 6.8 mmol/L (161)

○ 6.9 mmol/L (162)

○ 7.0 mmol/L (163)

○ 7.1 mmol/L (164)

○ 7.2 mmol/L (165)

○ 7.3 mmol/L (166)

○ 7.4 mmol/L (167)

○ 7.5 mmol/L (168)

○ 7.6 mmol/L (169)

○ 7.7 mmol/L (170)

○ 7.8 mmol/L (171)

○ 7.9 mmol/L (172)

○ 8.0 mmol/L (173)

○ 8.1 mmol/L (174)

○ 8.2 mmol/L (175)

○ 8.3 mmol/L (176)

○ 8.4 mmol/L (177)

○ 8.5 mmol/L (178)

○ 8.6 mmol/L (179)

○ 8.7 mmol/L (180)

○ 8.8 mmol/L (181)

○ 8.9 mmol/L (182)

- 9.0 mmol/L (183)
- 9.1 mmol/L (184)
- 9.2 mmol/L (185)
- 9.3 mmol/L (186)
- 9.4 mmol/L (187)
- 9.5 mmol/L (188)
- 9.6 mmol/L (189)
- 9.7 mmol/L (190)
- 9.8 mmol/L (191)
- 9.9 mmol/L (192)
- 10.0 mmol/L (consensus value used in Ambulatory Glucose Profile reports) (193)
- 10.1 mmol/L (194)
- 10.2 mmol/L (195)
- 10.3 mmol/L (196)
- 10.4 mmol/L (197)
- 10.5 mmol/L (198)
- 10.6 mmol/L (199)
- 10.7 mmol/L (200)
- 10.8 mmol/L (201)
- 10.9 mmol/L (202)
- 11.0 mmol/L (203)

- 11.1 mmol/L (204)
- 11.2 mmol/L (205)
- 11.3 mmol/L (206)
- 11.4 mmol/L (207)
- 11.5 mmol/L (208)
- 11.6 mmol/L (209)
- 11.7 mmol/L (210)
- 11.8 mmol/L (211)
- 11.9 mmol/L (212)
- 12.0 mmol/L (213)
- 12.1 mmol/L (214)
- 12.2 mmol/L (215)
- 12.3 mmol/L (216)
- 12.4 mmol/L (217)
- 12.5 mmol/L (218)
- 12.6 mmol/L (219)
- 12.7 mmol/L (220)
- 12.8 mmol/L (221)
- 12.9 mmol/L (222)
- 13.0 mmol/L (223)
- 13.1 mmol/L (224)

○ 13.2 mmol/L (225)

○ 13.3 mmol/L (226)

○ 13.4 mmol/L (227)

○ 13.5 mmol/L (228)

○ 13.6 mmol/L (229)

○ 13.7 mmol/L (230)

○ 13.8 mmol/L (231)

○ 13.9 mmol/L (232)

○ 14.0 mmol/L (233)

○ 14.1 mmol/L (234)

○ 14.2 mmol/L (235)

○ 14.3 mmol/L (236)

○ 14.4 mmol/L (237)

○ 14.5 mmol/L (238)

○ 14.6 mmol/L (239)

○ 14.7 mmol/L (240)

○ 14.8 mmol/L (241)

○ 14.9 mmol/L (242)

○ 15.0 mmol/L (243)

○ above 15.0 mmol/L (244)

○ I don’t know (245)

○ I prefer not to answer (246)

*Display this question:*

*If Does your child have a specific time in range goal? (choose one) = Yes*

Comments about your child's target range (optional)

________________________________________________________________

________________________________________________________________

________________________________________________________________

________________________________________________________________

________________________________________________________________

*Display this question:*

*If Does your child have a specific time in range goal? (choose one) = Yes*

What is the goal for the percentage of time they spend in their target range? The goal is to spend at least this percent of time in their target range:

- 0% (4)
- 1% (5)
- 2% (6)
- 3% (7)
- 4% (8)
- 5% (9)
- 6% (10)
- 7% (11)
- 8% (12)
- 9% (13)
- 10% (14)
- 11% (15)
- 12% (16)
- 13% (17)
- 14% (18)
- 15% (19)
- 16% (20)
- 17% (21)
- 18% (22)
- 19% (23)

- 20% (24)
- 21% (25)
- 22% (26)
- 23% (27)
- 24% (28)
- 25% (29)
- 26% (30)
- 27% (31)
- 28% (32)
- 29% (33)
- 30% (34)
- 31% (35)
- 32% (36)
- 33% (37)
- 34% (38)
- 35% (39)
- 36% (40)
- 37% (41)
- 38% (42)
- 39% (43)
- 40% (44)

- 41% (45)
- 42% (46)
- 43% (47)
- 44% (48)
- 45% (49)
- 46% (50)
- 47% (51)
- 48% (52)
- 49% (53)
- 50% (54)
- 51% (55)
- 52% (56)
- 53% (57)
- 54% (58)
- 55% (59)
- 56% (60)
- 57% (61)
- 58% (62)
- 59% (63)
- 60% (64)
- 61% (65)

○ 62%  (66)

○ 63%  (67)

○ 64%  (68)

○ 65%  (69)

○ 66%  (70)

○ 67%  (71)

○ 68%  (72)

○ 69%  (73)

○ 70% (consensus value used in Ambulatory Glucose Profile reports)  (74)

○ 71%  (75)

○ 72%  (76)

○ 73%  (77)

○ 74%  (78)

○ 75%  (79)

○ 76%  (80)

○ 77%  (81)

○ 78%  (82)

○ 79%  (83)

○ 80%  (84)

○ 81%  (85)

○ 82%  (86)

○ 83% (87)

○ 84% (88)

○ 85% (89)

○ 86% (90)

○ 87% (91)

○ 88% (92)

○ 89% (93)

○ 90% (94)

○ 91% (95)

○ 92% (96)

○ 93% (97)

○ 94% (98)

○ 95% (99)

○ 96% (100)

○ 97% (101)

○ 98% (102)

○ 99% (103)

○ 100% (104)

○ I don’t know (105)

○ I prefer not to answer (106)

*Display this question:*

*If Does your child have a specific time in range goal? (choose one) = Yes*

Who set this goal? (choose one)

- ◯ My child’s health care provider (endocrinologist, other doctor, diabetes educator, etc.) set the goal. (7)
- ◯ My child’s health care provider and our family set the goal together. (8)
- ◯ Our family set the goal. (9)
- ◯ Other (10)
- ◯ I don’t know (11)
- ◯ I prefer not to answer (12)

---

*Display this question:*

*If Are you the parent or guardian of one or more children under age 18 with type 1 diabetes (T1D) or... = Yes*

Has your child had a hemoglobin A1c test within the past 6 months? (choose one)

- ◯ Yes (1)
- ◯ No (2)
- ◯ I don’t know (3)
- ◯ I prefer not to answer (4)

---

*Display this question:*

*If Has your child had a hemoglobin A1c test within the past 6 months? (choose one) = Yes*

What was their most recent hemoglobin A1c result?

- under 4.0% (under 20 mmol/mol) (346)
- 4.0% (20 mmol/mol) (347)
- 4.1% (21 mmol/mol) (348)
- 4.2% (22 mmol/mol) (349)
- 4.3% (23 mmol/mol) (350)
- 4.4% (25 mmol/mol) (351)
- 4.5% (26 mmol/mol) (352)
- 4.6% (27 mmol/mol) (353)
- 4.7% (28 mmol/mol) (354)
- 4.8% (29 mmol/mol) (355)
- 4.9% (30 mmol/mol) (356)
- 5.0% (31 mmol/mol) (357)
- 5.1% (32 mmol/mol) (358)
- 5.2% (33 mmol/mol) (359)
- 5.3% (34 mmol/mol) (360)
- 5.4% (36 mmol/mol) (361)
- 5.5% (37 mmol/mol) (362)
- 5.6% (38 mmol/mol) (363)
- 5.7% (39 mmol/mol) (364)
- 5.8% (40 mmol/mol) (365)
- 5.9% (41 mmol/mol) (366)

○ 6.0% (42 mmol/mol) (367)

○ 6.1% (43 mmol/mol) (368)

○ 6.2% (44 mmol/mol) (369)

○ 6.3% (45 mmol/mol) (370)

○ 6.4% (46 mmol/mol) (371)

○ 6.5% (48 mmol/mol) (372)

○ 6.6% (49 mmol/mol) (373)

○ 6.7% (50 mmol/mol) (374)

○ 6.8% (51 mmol/mol) (375)

○ 6.9% (52 mmol/mol) (376)

○ 7.0% (53 mmol/mol) (377)

○ 7.1% (54 mmol/mol) (378)

○ 7.2% (55 mmol/mol) (379)

○ 7.3% (56 mmol/mol) (380)

○ 7.4% (57 mmol/mol) (381)

○ 7.5% (58 mmol/mol) (382)

○ 7.6% (59 mmol/mol) (383)

○ 7.7% (60 mmol/mol) (384)

○ 7.8% (62 mmol/mol) (385)

○ 7.9% (63 mmol/mol) (386)

○ 8.0% (64 mmol/mol) (387)

○ 8.1% (65 mmol/mol) (388)

○ 8.2% (66 mmol/mol) (389)

○ 8.3% (67 mmol/mol) (390)

○ 8.4% (68 mmol/mol) (391)

○ 8.5% (69 mmol/mol) (392)

○ 8.6% (70 mmol/mol) (393)

○ 8.7% (72 mmol/mol) (394)

○ 8.8% (73 mmol/mol) (395)

○ 8.9% (74 mmol/mol) (396)

○ 9.0% (75 mmol/mol) (397)

○ 9.1% (76 mmol/mol) (398)

○ 9.2% (77 mmol/mol) (399)

○ 9.3% (78 mmol/mol) (400)

○ 9.4% (79 mmol/mol) (401)

○ 9.5% (80 mmol/mol) (402)

○ 9.6% (81 mmol/mol) (403)

○ 9.7% (83 mmol/mol) (404)

○ 9.8% (84 mmol/mol) (405)

○ 9.9% (85 mmol/mol) (406)

○ 10.0% (86 mmol/mol) (407)

○ 10.1% (87 mmol/mol) (408)

○ 10.2% (88 mmol/mol) (409)

○ 10.3% (89 mmol/mol) (410)

○ 10.4% (90 mmol/mol) (411)

○ 10.5% (91 mmol/mol) (412)

○ 10.6% (92 mmol/mol) (413)

○ 10.7% (93 mmol/mol) (414)

○ 10.8% (94 mmol/mol) (415)

○ 10.9% (95 mmol/mol) (416)

○ 11.0% (97 mmol/mol) (417)

○ 11.1% (98 mmol/mol) (418)

○ 11.2% (99 mmol/mol) (419)

○ 11.3% (100 mmol/mol) (420)

○ 11.4% (101 mmol/mol) (421)

○ 11.5% (102 mmol/mol) (422)

○ 11.6% (103 mmol/mol) (423)

○ 11.7% (104 mmol/mol) (424)

○ 11.8% (105 mmol/mol) (425)

○ 11.9% (106 mmol/mol) (426)

○ 12.0% (108 mmol/mol) (427)

○ 12.1% (109 mmol/mol) (428)

○ 12.2% (110 mmol/mol) (429)

- 12.3% (111 mmol/mol)  (430)
- 12.4% (112 mmol/mol)  (431)
- 12.5% (113 mmol/mol)  (432)
- 12.6% (114 mmol/mol)  (433)
- 12.7% (115 mmol/mol)  (434)
- 12.8% (116 mmol/mol)  (435)
- 12.9% (117 mmol/mol)  (436)
- 13.0% (119 mmol/mol)  (437)
- 13.1% (120 mmol/mol)  (438)
- 13.2% (121 mmol/mol)  (439)
- 13.3% (122 mmol/mol)  (440)
- 13.4% (123 mmol/mol)  (441)
- 13.5% (124 mmol/mol)  (442)
- 13.6% (125 mmol/mol)  (443)
- 13.7% (126 mmol/mol)  (444)
- 13.8% (127 mmol/mol)  (445)
- 13.9% (128 mmol/mol)  (446)
- 14.0% (130 mmol/mol)  (447)
- 14.1% (131 mmol/mol)  (448)
- 14.2% (132 mmol/mol)  (449)
- 14.3% (133 mmol/mol)  (450)

○ 14.4% (134 mmol/mol) (451)

○ 14.5% (135 mmol/mol) (452)

○ 14.6% (136 mmol/mol) (453)

○ 14.7% (137 mmol/mol) (454)

○ 14.8% (138 mmol/mol) (455)

○ 14.9% (139 mmol/mol) (456)

○ 15.0% (140 mmol/mol) (457)

○ Above 15.0% (above 140 mmol/mol) (458)

○ I don't know (459)

○ I prefer not to answer (460)

---

*Display this question:*

*If Has your child had a hemoglobin A1c test within the past 6 months? (choose one) = Yes*

When was this hemoglobin A1c test done?

- This month (1)
- Last month (2)
- 2 months ago (3)
- 3 months ago (4)
- 4 months ago (5)
- 5 months ago (6)
- 6 months ago (7)
- More than 6 months ago (8)
- I don’t know (9)
- I prefer not to answer (10)

---

*Display this question:*

*If Does your child use a continuous glucose monitor or flash glucose monitor (e.g., Dexcom, Libre, M... = Yes*

Do you know how to check Time in Range in your child's or children's CGM app(s)? (choose one)

- Yes (1)
- No (2)
- I don’t know (3)
- I prefer not to answer (4)

---

*Display this question:*

*If Do you know how to check Time in Range in your child's or children's CGM app(s)? (choose one) = Yes*

Over the past 90 days, what was your child's Time in Range for the standard range 3.9 mmol/L - 10 mmol/L?

- ○ 0% (1)
- ○ 1% (2)
- ○ 2% (3)
- ○ 3% (4)
- ○ 4% (5)
- ○ 5% (6)
- ○ 6% (7)
- ○ 7% (8)
- ○ 8% (9)
- ○ 9% (10)
- ○ 10% (11)
- ○ 11% (12)
- ○ 12% (13)
- ○ 13% (14)
- ○ 14% (15)
- ○ 15% (16)
- ○ 16% (17)
- ○ 17% (18)
- ○ 18% (19)
- ○ 19% (20)

○ 20% (21)

○ 21% (22)

○ 22% (23)

○ 23% (24)

○ 24% (25)

○ 25% (26)

○ 26% (27)

○ 27% (28)

○ 28% (29)

○ 29% (30)

○ 30% (31)

○ 31% (32)

○ 32% (33)

○ 33% (34)

○ 34% (35)

○ 35% (36)

○ 36% (37)

○ 37% (38)

○ 38% (39)

○ 39% (40)

○ 40% (41)

- 41% (42)
- 42% (43)
- 43% (44)
- 44% (45)
- 45% (46)
- 46% (47)
- 47% (48)
- 48% (49)
- 49% (50)
- 50% (51)
- 51% (52)
- 52% (53)
- 53% (54)
- 54% (55)
- 55% (56)
- 56% (57)
- 57% (58)
- 58% (59)
- 59% (60)
- 60% (61)
- 61% (62)

○ 62% (63)

○ 63% (64)

○ 64% (65)

○ 65% (66)

○ 66% (67)

○ 67% (68)

○ 68% (69)

○ 69% (70)

○ 70% (71)

○ 71% (72)

○ 72% (73)

○ 73% (74)

○ 74% (75)

○ 75% (76)

○ 76% (77)

○ 77% (78)

○ 78% (79)

○ 79% (80)

○ 80% (81)

○ 81% (82)

○ 82% (83)

○ 83%  (84)

○ 84%  (85)

○ 85%  (86)

○ 86%  (87)

○ 87%  (88)

○ 88%  (89)

○ 89%  (90)

○ 90%  (91)

○ 91%  (92)

○ 92%  (93)

○ 93%  (94)

○ 94%  (95)

○ 95%  (96)

○ 96%  (97)

○ 97%  (98)

○ 98%  (99)

○ 99%  (100)

○ 100%  (101)

○ I don't know    (102)

○ I prefer not to answer  (103)

*Display this question:*

*If Are you the parent or guardian of one or more children under age 18 with type 1 diabetes (T1D) or... = Yes*

Over the past 6 months, has your child had any **severe** hypoglycemic (low blood sugar) events in which they required help from someone else? (choose one)

- Yes (1)
- No (2)
- I don't know (3)
- I prefer not to answer (4)

---

*Display this question:*

*If Over the past 6 months, has your child had any severe hypoglycemic (low blood sugar) events in wh... = Yes*

When your child required help from someone else for the severe hypoglycemic (low blood sugar) event(s), did that help include: (check all that apply)

- ▢ Emergency services (e.g., someone called 911) (1)
- ▢ Glucagon (e.g., someone gave your child glucagon by needle or by nose) (2)
- ▢ Hospitalization (your child received treatment at a hospital) (3)
- ▢ Someone getting low treatment (e.g., juice) and bringing it to your child (4)
- ▢ I don't know (5)
- ▢ I prefer not to answer (6)

---

*Display this question:*

*If How many children under age 18 with type 1 diabetes (T1D) or a similar type of diabetes (such as... = 2*

Thank you. Now we will repeat the previous questions, this time for your younger child with T1D (or a similar type of diabetes).

*Display this question:*

*If How many children under age 18 with type 1 diabetes (T1D) or a similar type of diabetes (such as... = 3*

*Or How many children under age 18 with type 1 diabetes (T1D) or a similar type of diabetes (such as... = More than 3*

Thank you. Now we will repeat the previous questions, this time for your second-oldest child with T1D (or a similar type of diabetes).

*Display this question:*

*If How many children under age 18 with type 1 diabetes (T1D) or a similar type of diabetes (such as... = 2*

*Or How many children under age 18 with type 1 diabetes (T1D) or a similar type of diabetes (such as... = 3*

*Or How many children under age 18 with type 1 diabetes (T1D) or a similar type of diabetes (such as... = More than 3*

Does your child have one or more health professionals (doctors, nurses, dietitians, pharmacists, etc.) who provide regular T1D care for them? By “regular” we mean at least once a year. (choose one)

- ◯ Yes (1)
- ◯ No (2)
- ◯ I don’t know (3)
- ◯ I prefer not to answer (4)

*Display this question:*

*If Does your child have one or more health professionals (doctors, nurses, dietitians, pharmacists,... = Yes*

What kind(s) of health professionals provide regular (at least once a year) diabetes care for your child? (check all that apply)

- ▢ Diabetes educator (also known as certified diabetes educator/CDE) (13)
- ▢ Diabetologist (14)
- ▢ Dietitian (15)
- ▢ Doctor (but I don't know what kind) (16)
- ▢ Endocrinologist (17)
- ▢ Family doctor (18)
- ▢ Internal medicine doctor (19)
- ▢ Mental health specialist (20)
- ▢ Nurse (21)
- ▢ Pharmacist (22)
- ▢ I don't know (23)
- ▢ I prefer not to answer (24)

*Display this question:*

*If How many children under age 18 with type 1 diabetes (T1D) or a similar type of diabetes (such as... = 2*

*Or How many children under age 18 with type 1 diabetes (T1D) or a similar type of diabetes (such as... = 3*

*Or How many children under age 18 with type 1 diabetes (T1D) or a similar type of diabetes (such as... = More than 3*

Does your child have a specific goal for their hemoglobin A1c (the blood test that many people with diabetes get every 3-6 months)? (choose one)

- ○ Yes (1)
- ○ No (2)
- ○ I don't know (3)
- ○ I prefer not to answer (4)

---

*Display this question:*

*If Does your child have a specific goal for their hemoglobin A1c (the blood test that many people wi... = Yes*

What is their goal for their hemoglobin A1c? (choose one) *In this question, we present A1c in %, the common presentation in Canada. If you use mmol/mol (the current international standard), here are the equivalent numbers: 4.0% = 20 mmol/mol; 5.6% = 38 mmol/mol; 6.0% =*

*42 mmol/mol; 6.1% = 43 mmol/mol; 6.5% = 48 mmol/mol; 7.0% = 53 mmol/mol; 7.5% = 58 mmol/mol; 8.0% = 64 mmol/mol; 8.5% = 69 mmol/mol*

- ○ 4.0% to 5.6% (the nondiabetic range) (1)
- ○ 6.0% or lower (2)
- ○ 6.1% or lower (the recommended goal in Canada when pregnant with T1D) (3)
- ○ 6.5% or lower (the recommended goal in Canada when planning a pregnancy with T1D) (4)
- ○ 7.0% or lower (the recommended goal in Canada for most adults with T1D) (5)
- ○ 7.5% or lower (the recommended goal in Canada for most children and adolescents under 18 years old with T1D) (6)
- ○ 8.0% or lower (the recommended goal in Canada for some people who are older or for other reasons) (7)
- ○ 8.5% or lower (the recommended goal in Canada for some people who are older, who have severe hypo unawareness, or for other reasons) (8)
- ○ other (9) ________________________________________________
- ○ I prefer not to answer (10)

*Display this question:*

*If What is their goal for their hemoglobin A1c? (choose one) In this question, we present A1c in %,... = 4.0% to 5.6% (the nondiabetic range)*

*Or What is their goal for their hemoglobin A1c? (choose one) In this question, we present A1c in %,... = 6.0% or lower*

*Or What is their goal for their hemoglobin A1c? (choose one) In this question, we present A1c in %,... = 6.1% or lower (the recommended goal in Canada when pregnant with T1D)*

*Or What is their goal for their hemoglobin A1c? (choose one) In this question, we present A1c in %,... = 6.5% or lower (the recommended goal in Canada when planning a pregnancy with T1D)*

*Or What is their goal for their hemoglobin A1c? (choose one) In this question, we present A1c in %,... = 7.0% or lower (the recommended goal in Canada for most adults with T1D)*

*Or What is their goal for their hemoglobin A1c? (choose one) In this question, we present A1c in %,... = 7.5% or lower (the recommended goal in Canada for most children and adolescents under 18 years old with T1D)*

*Or What is their goal for their hemoglobin A1c? (choose one) In this question, we present A1c in %,... = 8.0% or lower (the recommended goal in Canada for some people who are older or for other reasons)*

*Or What is their goal for their hemoglobin A1c? (choose one) In this question, we present A1c in %,... = 8.5% or lower (the recommended goal in Canada for some people who are older, who have severe hypo unawareness, or for other reasons)*

*Or Or Text Response Is Not Empty*

Who set this goal? (choose one)

- ◯ My child’s health care provider (endocrinologist, other doctor, diabetes educator, etc.) set the goal. (7)
- ◯ My child’s health care provider and our family set the goal together. (8)
- ◯ Our family set the goal. (9)
- ◯ Other (10) ________________________________________________
- ◯ I don’t know (11)
- ◯ I prefer not to answer (12)

*Display this question:*

*If How many children under age 18 with type 1 diabetes (T1D) or a similar type of diabetes (such as... = 2*

*Or How many children under age 18 with type 1 diabetes (T1D) or a similar type of diabetes (such as... = 3*

*Or How many children under age 18 with type 1 diabetes (T1D) or a similar type of diabetes (such as... = More than 3*

Does your child use a continuous glucose monitor or flash glucose monitor (e.g., Dexcom, Libre, Medtronic Guardian sensor)?

- ○ Yes (1)
- ○ No (2)
- ○ I don't know (3)
- ○ I prefer not to answer (4)

---

*Display this question:*

*If Does your child use a continuous glucose monitor or flash glucose monitor (e.g., Dexcom, Libre, M... = Yes*

Does your child have a specific time in range goal? (choose one)

- ○ Yes (1)
- ○ No (2)
- ○ I don't know (3)
- ○ I prefer not to answer (4)

---

*Display this question:*

*If Does your child have a specific time in range goal? (choose one) = Yes*

What is the lower limit of the target range for their time in range goal? (choose one) *In this question, we present blood glucose values in mmol/L, the common presentation in Canada. If you use mg/dl, here are some equivalent numbers: 70 mg/dl = 3.9 mmol/L; 80 mg/dl = 4.4 mmol/L; 90 mg/dl = 5.0 mmol/L; 100 mg/dl = 5.6 mmol/L; 110 mg/dl = 6.1 mmol/L; 120 mg/dl =*

*6.7 mmol/L; 130 mg/dl = 7.2 mmol/L; 140 mg/dl = 7.8 mmol/L; 150 mg/dl = 8.3 mmol/L; 160 mg/dl = 8.9 mmol/L; 170 mg/dl = 9.4 mmol/L; 180 mg/dl = 10.0 mmol/L; 190 mg/dl = 10.6 mmol/L; 200 mg/dl = 11.1 mmol/L*

- ○ below 3.6 mmol/L (5)
- ○ 3.6 mmol/L (105)
- ○ 3.7 mmol/L (106)
- ○ 3.8 mmol/L (107)
- ○ 3.9 mmol/L (consensus value used in Ambulatory Glucose Profile reports) (108)
- ○ 4.0 mmol/L (109)
- ○ 4.1 mmol/L (110)
- ○ 4.2 mmol/L (111)
- ○ 4.3 mmol/L (112)
- ○ 4.4 mmol/L (113)
- ○ 4.5 mmol/L (114)
- ○ 4.6 mmol/L (115)
- ○ 4.7 mmol/L (116)
- ○ 4.8 mmol/L (117)
- ○ 4.9 mmol/L (118)
- ○ 5.0 mmol/L (119)
- ○ 5.1 mmol/L (120)
- ○ 5.2 mmol/L (121)
- ○ 5.3 mmol/L (122)
- ○ 5.4 mmol/L (123)

○ 5.5 mmol/L (124)

○ 5.6 mmol/L (125)

○ 5.7 mmol/L (126)

○ 5.8 mmol/L (127)

○ 5.9 mmol/L (128)

○ 6.0 mmol/L (129)

○ 6.1 mmol/L (130)

○ 6.2 mmol/L (131)

○ 6.3 mmol/L (132)

○ 6.4 mmol/L (133)

○ 6.5 mmol/L (134)

○ 6.6 mmol/L (135)

○ 6.7 mmol/L (136)

○ 6.8 mmol/L (137)

○ 6.9 mmol/L (138)

○ 7.0 mmol/L (139)

○ above 7.0 mmol/L (140)

○ I don’t know (141)

○ I prefer not to answer (142)

---

*Display this question:*

*If Does your child have a specific time in range goal? (choose one) = Yes*

What is the upper limit of the target range for their time in range goal? (choose one) *In this question, we present blood glucose values in mmol/L, the common presentation in Canada. If you use mg/dl, here are some equivalent numbers: 70 mg/dl = 3.9 mmol/L; 80 mg/dl = 4.4 mmol/L; 90 mg/dl = 5.0 mmol/L; 100 mg/dl = 5.6 mmol/L; 110 mg/dl = 6.1 mmol/L; 120 mg/dl = 6.7 mmol/L; 130 mg/dl = 7.2 mmol/L; 140 mg/dl = 7.8 mmol/L; 150 mg/dl = 8.3 mmol/L; 160*

*mg/dl = 8.9 mmol/L; 170 mg/dl = 9.4 mmol/L; 180 mg/dl = 10.0 mmol/L; 190 mg/dl = 10.6 mmol/L; 200 mg/dl = 11.1 mmol/L*

- ◯ under 5.0 mmol/L (5)
- ◯ 5.0 mmol/L (143)
- ◯ 5.1 mmol/L (144)
- ◯ 5.2 mmol/L (145)
- ◯ 5.3 mmol/L (146)
- ◯ 5.4 mmol/L (147)
- ◯ 5.5 mmol/L (148)
- ◯ 5.6 mmol/L (149)
- ◯ 5.7 mmol/L (150)
- ◯ 5.8 mmol/L (151)
- ◯ 5.9 mmol/L (152)
- ◯ 6.0 mmol/L (153)
- ◯ 6.1 mmol/L (154)
- ◯ 6.2 mmol/L (155)
- ◯ 6.3 mmol/L (156)
- ◯ 6.4 mmol/L (157)
- ◯ 6.5 mmol/L (158)
- ◯ 6.6 mmol/L (159)
- ◯ 6.7 mmol/L (160)
- ◯ 6.8 mmol/L (161)

- 6.9 mmol/L (162)
- 7.0 mmol/L (163)
- 7.1 mmol/L (164)
- 7.2 mmol/L (165)
- 7.3 mmol/L (166)
- 7.4 mmol/L (167)
- 7.5 mmol/L (168)
- 7.6 mmol/L (169)
- 7.7 mmol/L (170)
- 7.8 mmol/L (171)
- 7.9 mmol/L (172)
- 8.0 mmol/L (173)
- 8.1 mmol/L (174)
- 8.2 mmol/L (175)
- 8.3 mmol/L (176)
- 8.4 mmol/L (177)
- 8.5 mmol/L (178)
- 8.6 mmol/L (179)
- 8.7 mmol/L (180)
- 8.8 mmol/L (181)
- 8.9 mmol/L (182)

- 9.0 mmol/L  (183)
- 9.1 mmol/L  (184)
- 9.2 mmol/L  (185)
- 9.3 mmol/L  (186)
- 9.4 mmol/L  (187)
- 9.5 mmol/L  (188)
- 9.6 mmol/L  (189)
- 9.7 mmol/L  (190)
- 9.8 mmol/L  (191)
- 9.9 mmol/L  (192)
- 10.0 mmol/L (consensus value used in Ambulatory Glucose Profile reports)  (193)
- 10.1 mmol/L  (194)
- 10.2 mmol/L  (195)
- 10.3 mmol/L  (196)
- 10.4 mmol/L  (197)
- 10.5 mmol/L  (198)
- 10.6 mmol/L  (199)
- 10.7 mmol/L  (200)
- 10.8 mmol/L  (201)
- 10.9 mmol/L  (202)
- 11.0 mmol/L  (203)

- ○ 11.1 mmol/L (204)
- ○ 11.2 mmol/L (205)
- ○ 11.3 mmol/L (206)
- ○ 11.4 mmol/L (207)
- ○ 11.5 mmol/L (208)
- ○ 11.6 mmol/L (209)
- ○ 11.7 mmol/L (210)
- ○ 11.8 mmol/L (211)
- ○ 11.9 mmol/L (212)
- ○ 12.0 mmol/L (213)
- ○ 12.1 mmol/L (214)
- ○ 12.2 mmol/L (215)
- ○ 12.3 mmol/L (216)
- ○ 12.4 mmol/L (217)
- ○ 12.5 mmol/L (218)
- ○ 12.6 mmol/L (219)
- ○ 12.7 mmol/L (220)
- ○ 12.8 mmol/L (221)
- ○ 12.9 mmol/L (222)
- ○ 13.0 mmol/L (223)
- ○ 13.1 mmol/L (224)

○ 13.2 mmol/L (225)

○ 13.3 mmol/L (226)

○ 13.4 mmol/L (227)

○ 13.5 mmol/L (228)

○ 13.6 mmol/L (229)

○ 13.7 mmol/L (230)

○ 13.8 mmol/L (231)

○ 13.9 mmol/L (232)

○ 14.0 mmol/L (233)

○ 14.1 mmol/L (234)

○ 14.2 mmol/L (235)

○ 14.3 mmol/L (236)

○ 14.4 mmol/L (237)

○ 14.5 mmol/L (238)

○ 14.6 mmol/L (239)

○ 14.7 mmol/L (240)

○ 14.8 mmol/L (241)

○ 14.9 mmol/L (242)

○ 15.0 mmol/L (243)

○ above 15.0 mmol/L (244)

○ I don’t know (245)

◯ I prefer not to answer (246)

*Display this question:*

*If Does your child have a specific time in range goal? (choose one) = Yes*

Comments about your child's target range (optional)

________________________________________________________________

________________________________________________________________

________________________________________________________________

________________________________________________________________

________________________________________________________________

*Display this question:*

*If Does your child have a specific time in range goal? (choose one) = Yes*

What is the goal for the percentage of time they spend in their target range? The goal is to spend at least this percent of time in their target range:

- ○ 0% (4)
- ○ 1% (5)
- ○ 2% (6)
- ○ 3% (7)
- ○ 4% (8)
- ○ 5% (9)
- ○ 6% (10)
- ○ 7% (11)
- ○ 8% (12)
- ○ 9% (13)
- ○ 10% (14)
- ○ 11% (15)
- ○ 12% (16)
- ○ 13% (17)
- ○ 14% (18)
- ○ 15% (19)
- ○ 16% (20)
- ○ 17% (21)
- ○ 18% (22)
- ○ 19% (23)

- 20% (24)
- 21% (25)
- 22% (26)
- 23% (27)
- 24% (28)
- 25% (29)
- 26% (30)
- 27% (31)
- 28% (32)
- 29% (33)
- 30% (34)
- 31% (35)
- 32% (36)
- 33% (37)
- 34% (38)
- 35% (39)
- 36% (40)
- 37% (41)
- 38% (42)
- 39% (43)
- 40% (44)

- ○ 41% (45)
- ○ 42% (46)
- ○ 43% (47)
- ○ 44% (48)
- ○ 45% (49)
- ○ 46% (50)
- ○ 47% (51)
- ○ 48% (52)
- ○ 49% (53)
- ○ 50% (54)
- ○ 51% (55)
- ○ 52% (56)
- ○ 53% (57)
- ○ 54% (58)
- ○ 55% (59)
- ○ 56% (60)
- ○ 57% (61)
- ○ 58% (62)
- ○ 59% (63)
- ○ 60% (64)
- ○ 61% (65)

○ 62%  (66)

○ 63%  (67)

○ 64%  (68)

○ 65%  (69)

○ 66%  (70)

○ 67%  (71)

○ 68%  (72)

○ 69%  (73)

○ 70% (consensus value used in Ambulatory Glucose Profile reports)  (74)

○ 71%  (75)

○ 72%  (76)

○ 73%  (77)

○ 74%  (78)

○ 75%  (79)

○ 76%  (80)

○ 77%  (81)

○ 78%  (82)

○ 79%  (83)

○ 80%  (84)

○ 81%  (85)

○ 82%  (86)

○ 83% (87)

○ 84% (88)

○ 85% (89)

○ 86% (90)

○ 87% (91)

○ 88% (92)

○ 89% (93)

○ 90% (94)

○ 91% (95)

○ 92% (96)

○ 93% (97)

○ 94% (98)

○ 95% (99)

○ 96% (100)

○ 97% (101)

○ 98% (102)

○ 99% (103)

○ 100% (104)

○ I don’t know (105)

○ I prefer not to answer (106)

*Display this question:*

*If Does your child have a specific time in range goal? (choose one) = Yes*

Who set this goal? (choose one)

- ◯ My child's health care provider (endocrinologist, other doctor, diabetes educator, etc.) set the goal. (7)
- ◯ My child's health care provider and our family set the goal together. (8)
- ◯ Our family set the goal. (9)
- ◯ Other (10)
- ◯ I don't know (11)
- ◯ I prefer not to answer (12)

---

*Display this question:*

*If How many children under age 18 with type 1 diabetes (T1D) or a similar type of diabetes (such as... = 2*

*Or How many children under age 18 with type 1 diabetes (T1D) or a similar type of diabetes (such as... = 3*

*Or How many children under age 18 with type 1 diabetes (T1D) or a similar type of diabetes (such as... = More than 3*

Has your child had a hemoglobin A1c test within the past 6 months? (choose one)

- ◯ Yes (1)
- ◯ No (2)
- ◯ I don't know (3)
- ◯ I prefer not to answer (4)

---

*Display this question:*

*If Has your child had a hemoglobin A1c test within the past 6 months? (choose one) = Yes*

What was their most recent hemoglobin A1c result?

- under 4.0% (under 20 mmol/mol) (346)
- 4.0% (20 mmol/mol) (347)
- 4.1% (21 mmol/mol) (348)
- 4.2% (22 mmol/mol) (349)
- 4.3% (23 mmol/mol) (350)
- 4.4% (25 mmol/mol) (351)
- 4.5% (26 mmol/mol) (352)
- 4.6% (27 mmol/mol) (353)
- 4.7% (28 mmol/mol) (354)
- 4.8% (29 mmol/mol) (355)
- 4.9% (30 mmol/mol) (356)
- 5.0% (31 mmol/mol) (357)
- 5.1% (32 mmol/mol) (358)
- 5.2% (33 mmol/mol) (359)
- 5.3% (34 mmol/mol) (360)
- 5.4% (36 mmol/mol) (361)
- 5.5% (37 mmol/mol) (362)
- 5.6% (38 mmol/mol) (363)
- 5.7% (39 mmol/mol) (364)
- 5.8% (40 mmol/mol) (365)
- 5.9% (41 mmol/mol) (366)

- ○ 6.0% (42 mmol/mol) (367)
- ○ 6.1% (43 mmol/mol) (368)
- ○ 6.2% (44 mmol/mol) (369)
- ○ 6.3% (45 mmol/mol) (370)
- ○ 6.4% (46 mmol/mol) (371)
- ○ 6.5% (48 mmol/mol) (372)
- ○ 6.6% (49 mmol/mol) (373)
- ○ 6.7% (50 mmol/mol) (374)
- ○ 6.8% (51 mmol/mol) (375)
- ○ 6.9% (52 mmol/mol) (376)
- ○ 7.0% (53 mmol/mol) (377)
- ○ 7.1% (54 mmol/mol) (378)
- ○ 7.2% (55 mmol/mol) (379)
- ○ 7.3% (56 mmol/mol) (380)
- ○ 7.4% (57 mmol/mol) (381)
- ○ 7.5% (58 mmol/mol) (382)
- ○ 7.6% (59 mmol/mol) (383)
- ○ 7.7% (60 mmol/mol) (384)
- ○ 7.8% (62 mmol/mol) (385)
- ○ 7.9% (63 mmol/mol) (386)
- ○ 8.0% (64 mmol/mol) (387)

○ 8.1% (65 mmol/mol) (388)

○ 8.2% (66 mmol/mol) (389)

○ 8.3% (67 mmol/mol) (390)

○ 8.4% (68 mmol/mol) (391)

○ 8.5% (69 mmol/mol) (392)

○ 8.6% (70 mmol/mol) (393)

○ 8.7% (72 mmol/mol) (394)

○ 8.8% (73 mmol/mol) (395)

○ 8.9% (74 mmol/mol) (396)

○ 9.0% (75 mmol/mol) (397)

○ 9.1% (76 mmol/mol) (398)

○ 9.2% (77 mmol/mol) (399)

○ 9.3% (78 mmol/mol) (400)

○ 9.4% (79 mmol/mol) (401)

○ 9.5% (80 mmol/mol) (402)

○ 9.6% (81 mmol/mol) (403)

○ 9.7% (83 mmol/mol) (404)

○ 9.8% (84 mmol/mol) (405)

○ 9.9% (85 mmol/mol) (406)

○ 10.0% (86 mmol/mol) (407)

○ 10.1% (87 mmol/mol) (408)

○ 10.2% (88 mmol/mol) (409)

○ 10.3% (89 mmol/mol) (410)

○ 10.4% (90 mmol/mol) (411)

○ 10.5% (91 mmol/mol) (412)

○ 10.6% (92 mmol/mol) (413)

○ 10.7% (93 mmol/mol) (414)

○ 10.8% (94 mmol/mol) (415)

○ 10.9% (95 mmol/mol) (416)

○ 11.0% (97 mmol/mol) (417)

○ 11.1% (98 mmol/mol) (418)

○ 11.2% (99 mmol/mol) (419)

○ 11.3% (100 mmol/mol) (420)

○ 11.4% (101 mmol/mol) (421)

○ 11.5% (102 mmol/mol) (422)

○ 11.6% (103 mmol/mol) (423)

○ 11.7% (104 mmol/mol) (424)

○ 11.8% (105 mmol/mol) (425)

○ 11.9% (106 mmol/mol) (426)

○ 12.0% (108 mmol/mol) (427)

○ 12.1% (109 mmol/mol) (428)

○ 12.2% (110 mmol/mol) (429)

○ 12.3% (111 mmol/mol) (430)

○ 12.4% (112 mmol/mol) (431)

○ 12.5% (113 mmol/mol) (432)

○ 12.6% (114 mmol/mol) (433)

○ 12.7% (115 mmol/mol) (434)

○ 12.8% (116 mmol/mol) (435)

○ 12.9% (117 mmol/mol) (436)

○ 13.0% (119 mmol/mol) (437)

○ 13.1% (120 mmol/mol) (438)

○ 13.2% (121 mmol/mol) (439)

○ 13.3% (122 mmol/mol) (440)

○ 13.4% (123 mmol/mol) (441)

○ 13.5% (124 mmol/mol) (442)

○ 13.6% (125 mmol/mol) (443)

○ 13.7% (126 mmol/mol) (444)

○ 13.8% (127 mmol/mol) (445)

○ 13.9% (128 mmol/mol) (446)

○ 14.0% (130 mmol/mol) (447)

○ 14.1% (131 mmol/mol) (448)

○ 14.2% (132 mmol/mol) (449)

○ 14.3% (133 mmol/mol) (450)

- ◯ 14.4% (134 mmol/mol) (451)
- ◯ 14.5% (135 mmol/mol) (452)
- ◯ 14.6% (136 mmol/mol) (453)
- ◯ 14.7% (137 mmol/mol) (454)
- ◯ 14.8% (138 mmol/mol) (455)
- ◯ 14.9% (139 mmol/mol) (456)
- ◯ 15.0% (140 mmol/mol) (457)
- ◯ Above 15.0% (above 140 mmol/mol) (458)
- ◯ I don't know (459)
- ◯ I prefer not to answer (460)

---

*Display this question:*

*If Has your child had a hemoglobin A1c test within the past 6 months? (choose one) = Yes*

When was this hemoglobin A1c test done?

- This month (1)
- Last month (2)
- 2 months ago (3)
- 3 months ago (4)
- 4 months ago (5)
- 5 months ago (6)
- 6 months ago (7)
- More than 6 months ago (8)
- I don’t know (9)
- I prefer not to answer (10)

---

*Display this question:*

*If Do you know how to check Time in Range in your child's or children's CGM app(s)? (choose one) = Yes*

*And If*

*How many children under age 18 with type 1 diabetes (T1D) or a similar type of diabetes (such as... = 2*

*Or How many children under age 18 with type 1 diabetes (T1D) or a similar type of diabetes (such as... = 3*

*Or How many children under age 18 with type 1 diabetes (T1D) or a similar type of diabetes (such as... = More than 3*

Over the past 90 days, what was your child's Time in Range for the standard range 3.9 mmol/L - 10 mmol/L?

- ◯ 0% (1)
- ◯ 1% (2)
- ◯ 2% (3)
- ◯ 3% (4)
- ◯ 4% (5)
- ◯ 5% (6)
- ◯ 6% (7)
- ◯ 7% (8)
- ◯ 8% (9)
- ◯ 9% (10)
- ◯ 10% (11)
- ◯ 11% (12)
- ◯ 12% (13)
- ◯ 13% (14)
- ◯ 14% (15)
- ◯ 15% (16)
- ◯ 16% (17)
- ◯ 17% (18)
- ◯ 18% (19)
- ◯ 19% (20)

- ○ 20% (21)
- ○ 21% (22)
- ○ 22% (23)
- ○ 23% (24)
- ○ 24% (25)
- ○ 25% (26)
- ○ 26% (27)
- ○ 27% (28)
- ○ 28% (29)
- ○ 29% (30)
- ○ 30% (31)
- ○ 31% (32)
- ○ 32% (33)
- ○ 33% (34)
- ○ 34% (35)
- ○ 35% (36)
- ○ 36% (37)
- ○ 37% (38)
- ○ 38% (39)
- ○ 39% (40)
- ○ 40% (41)

○ 41% (42)

○ 42% (43)

○ 43% (44)

○ 44% (45)

○ 45% (46)

○ 46% (47)

○ 47% (48)

○ 48% (49)

○ 49% (50)

○ 50% (51)

○ 51% (52)

○ 52% (53)

○ 53% (54)

○ 54% (55)

○ 55% (56)

○ 56% (57)

○ 57% (58)

○ 58% (59)

○ 59% (60)

○ 60% (61)

○ 61% (62)

- 62% (63)
- 63% (64)
- 64% (65)
- 65% (66)
- 66% (67)
- 67% (68)
- 68% (69)
- 69% (70)
- 70% (71)
- 71% (72)
- 72% (73)
- 73% (74)
- 74% (75)
- 75% (76)
- 76% (77)
- 77% (78)
- 78% (79)
- 79% (80)
- 80% (81)
- 81% (82)
- 82% (83)

○ 83% (84)

○ 84% (85)

○ 85% (86)

○ 86% (87)

○ 87% (88)

○ 88% (89)

○ 89% (90)

○ 90% (91)

○ 91% (92)

○ 92% (93)

○ 93% (94)

○ 94% (95)

○ 95% (96)

○ 96% (97)

○ 97% (98)

○ 98% (99)

○ 99% (100)

○ 100% (101)

○ I don't know (102)

○ I prefer not to answer (103)

*Display this question:*

*If How many children under age 18 with type 1 diabetes (T1D) or a similar type of diabetes (such as... = 2*

*Or How many children under age 18 with type 1 diabetes (T1D) or a similar type of diabetes (such as... = 3*

*Or How many children under age 18 with type 1 diabetes (T1D) or a similar type of diabetes (such as... = More than 3*

Over the past 6 months, has your child had any **severe** hypoglycemic (low blood sugar) events in which they required help from someone else? (choose one)

- ○ Yes (1)
- ○ No (2)
- ○ I don’t know (3)
- ○ I prefer not to answer (4)

---

*Display this question:*

*If Over the past 6 months, has your child had any severe hypoglycemic (low blood sugar) events in wh... = Yes*

When your child required help from someone else for the severe hypoglycemic (low blood sugar) event(s), did that help include: (check all that apply)

- ▢ Emergency services (e.g., someone called 911) (1)
- ▢ Glucagon (e.g., someone gave your child glucagon by needle or by nose) (2)
- ▢ Hospitalization (your child received treatment at a hospital) (3)
- ▢ Someone getting low treatment (e.g., juice) and bringing it to your child (4)
- ▢ I don’t know (5)
- ▢ I prefer not to answer (6)

Display this question:

*If How many children under age 18 with type 1 diabetes (T1D) or a similar type of diabetes (such as... = 3*

*Or How many children under age 18 with type 1 diabetes (T1D) or a similar type of diabetes (such as... = More than 3*

Thank you. Now we will repeat the previous questions, this time for your youngest child with T1D (or a similar type of diabetes).

*Display this question:*

*If How many children under age 18 with type 1 diabetes (T1D) or a similar type of diabetes (such as... = 3*

*Or How many children under age 18 with type 1 diabetes (T1D) or a similar type of diabetes (such as... = More than 3*

Does your child have one or more health professionals (doctors, nurses, dietitians, pharmacists, etc.) who provide regular T1D care for them? By "regular" we mean at least once a year. (choose one)

- Yes (1)
- No (2)
- I don't know (3)
- I prefer not to answer (4)

*Display this question:*

*If Does your child have one or more health professionals (doctors, nurses, dietitians, pharmacists,... = Yes*

What kind(s) of health professionals provide regular (at least once a year) diabetes care for your child? (check all that apply)

- ☐ Diabetes educator (also known as certified diabetes educator/CDE) (13)
- ☐ Diabetologist (14)
- ☐ Dietitian (15)
- ☐ Doctor (but I don't know what kind) (16)
- ☐ Endocrinologist (17)
- ☐ Family doctor (18)
- ☐ Internal medicine doctor (19)
- ☐ Mental health specialist (20)
- ☐ Nurse (21)
- ☐ Pharmacist (22)
- ☐ I don't know (23)
- ☐ I prefer not to answer (24)

---

*Display this question:*

*If How many children under age 18 with type 1 diabetes (T1D) or a similar type of diabetes (such as... = 3*

*Or How many children under age 18 with type 1 diabetes (T1D) or a similar type of diabetes (such as... = More than 3*

Does your child have a specific goal for their hemoglobin A1c (the blood test that many people with diabetes get every 3-6 months)? (choose one)

- ◯ Yes (1)
- ◯ No (2)
- ◯ I don't know (3)
- ◯ I prefer not to answer (4)

---

*Display this question:*

*If Does your child have a specific goal for their hemoglobin A1c (the blood test that many people wi... = Yes*

What is their goal for their hemoglobin A1c? (choose one) *In this question, we present A1c in %, the common presentation in Canada. If you use mmol/mol (the current international standard), here are the equivalent numbers: 4.0% = 20 mmol/mol; 5.6% = 38 mmol/mol; 6.0% =*

*42 mmol/mol; 6.1% = 43 mmol/mol; 6.5% = 48 mmol/mol; 7.0% = 53 mmol/mol; 7.5% = 58 mmol/mol; 8.0% = 64 mmol/mol; 8.5% = 69 mmol/mol*

◯ 4.0% to 5.6% (the nondiabetic range) (1)

◯ 6.0% or lower (2)

◯ 6.1% or lower (the recommended goal in Canada when pregnant with T1D) (3)

◯ 6.5% or lower (the recommended goal in Canada when planning a pregnancy with T1D) (4)

◯ 7.0% or lower (the recommended goal in Canada for most adults with T1D) (5)

◯ 7.5% or lower (the recommended goal in Canada for most children and adolescents under 18 years old with T1D) (6)

◯ 8.0% or lower (the recommended goal in Canada for some people who are older or for other reasons) (7)

◯ 8.5% or lower (the recommended goal in Canada for some people who are older, who have severe hypo unawareness, or for other reasons) (8)

◯ other (9) ________________________________________________

◯ I prefer not to answer (10)

*Display this question:*

*If What is their goal for their hemoglobin A1c? (choose one) In this question, we present A1c in %,... = 4.0% to 5.6% (the nondiabetic range)*

*Or What is their goal for their hemoglobin A1c? (choose one) In this question, we present A1c in %,... = 6.0% or lower*

*Or What is their goal for their hemoglobin A1c? (choose one) In this question, we present A1c in %,... = 6.1% or lower (the recommended goal in Canada when pregnant with T1D)*

*Or What is their goal for their hemoglobin A1c? (choose one) In this question, we present A1c in %,... = 6.5% or lower (the recommended goal in Canada when planning a pregnancy with T1D)*

*Or What is their goal for their hemoglobin A1c? (choose one) In this question, we present A1c in %,... = 7.0% or lower (the recommended goal in Canada for most adults with T1D)*

*Or What is their goal for their hemoglobin A1c? (choose one) In this question, we present A1c in %,... = 7.5% or lower (the recommended goal in Canada for most children and adolescents under 18 years old with T1D)*

*Or What is their goal for their hemoglobin A1c? (choose one) In this question, we present A1c in %,... = 8.0% or lower (the recommended goal in Canada for some people who are older or for other reasons)*

*Or What is their goal for their hemoglobin A1c? (choose one) In this question, we present A1c in %,... = 8.5% or lower (the recommended goal in Canada for some people who are older, who have severe hypo unawareness, or for other reasons)*

*Or Or Text Response Is Not Empty*

Who set this goal? (choose one)

- ◯ My child’s health care provider (endocrinologist, other doctor, diabetes educator, etc.) set the goal. (7)
- ◯ My child’s health care provider and our family set the goal together. (8)
- ◯ Our family set the goal. (9)
- ◯ Other (10) ________________________________________________
- ◯ I don’t know (11)
- ◯ I prefer not to answer (12)

---

*Display this question:*

*If How many children under age 18 with type 1 diabetes (T1D) or a similar type of diabetes (such as... = 3*

*Or How many children under age 18 with type 1 diabetes (T1D) or a similar type of diabetes (such as... = More than 3*

Does your child use a continuous glucose monitor or flash glucose monitor (e.g., Dexcom, Libre, Medtronic Guardian sensor)?

- ◯ Yes (1)
- ◯ No (2)
- ◯ I don’t know (3)
- ◯ I prefer not to answer (4)

---

*Display this question:*

*If Does your child use a continuous glucose monitor or flash glucose monitor (e.g., Dexcom, Libre, M... = Yes*

Does your child have a specific time in range goal? (choose one)

- ◯ Yes (1)
- ◯ No (2)
- ◯ I don’t know (3)
- ◯ I prefer not to answer (4)

---

*Display this question:*

*If Does your child have a specific time in range goal? (choose one) = Yes*

What is the lower limit of the target range for their time in range goal? (choose one) *In this question, we present blood glucose values in mmol/L, the common presentation in Canada. If you use mg/dl, here are some equivalent numbers: 70 mg/dl = 3.9 mmol/L; 80 mg/dl = 4.4 mmol/L; 90 mg/dl = 5.0 mmol/L; 100 mg/dl = 5.6 mmol/L; 110 mg/dl = 6.1 mmol/L; 120 mg/dl = 6.7 mmol/L; 130 mg/dl = 7.2 mmol/L; 140 mg/dl = 7.8 mmol/L; 150 mg/dl = 8.3 mmol/L; 160*

*mg/dl = 8.9 mmol/L; 170 mg/dl = 9.4 mmol/L; 180 mg/dl = 10.0 mmol/L; 190 mg/dl = 10.6 mmol/L; 200 mg/dl = 11.1 mmol/L*

- ○ below 3.6 mmol/L (5)
- ○ 3.6 mmol/L (105)
- ○ 3.7 mmol/L (106)
- ○ 3.8 mmol/L (107)
- ○ 3.9 mmol/L (consensus value used in Ambulatory Glucose Profile reports) (108)
- ○ 4.0 mmol/L (109)
- ○ 4.1 mmol/L (110)
- ○ 4.2 mmol/L (111)
- ○ 4.3 mmol/L (112)
- ○ 4.4 mmol/L (113)
- ○ 4.5 mmol/L (114)
- ○ 4.6 mmol/L (115)
- ○ 4.7 mmol/L (116)
- ○ 4.8 mmol/L (117)
- ○ 4.9 mmol/L (118)
- ○ 5.0 mmol/L (119)
- ○ 5.1 mmol/L (120)
- ○ 5.2 mmol/L (121)
- ○ 5.3 mmol/L (122)
- ○ 5.4 mmol/L (123)

- 5.5 mmol/L (124)
- 5.6 mmol/L (125)
- 5.7 mmol/L (126)
- 5.8 mmol/L (127)
- 5.9 mmol/L (128)
- 6.0 mmol/L (129)
- 6.1 mmol/L (130)
- 6.2 mmol/L (131)
- 6.3 mmol/L (132)
- 6.4 mmol/L (133)
- 6.5 mmol/L (134)
- 6.6 mmol/L (135)
- 6.7 mmol/L (136)
- 6.8 mmol/L (137)
- 6.9 mmol/L (138)
- 7.0 mmol/L (139)
- above 7.0 mmol/L (140)
- I don’t know (141)
- I prefer not to answer (142)

---

*Display this question:*

*If Does your child have a specific time in range goal? (choose one) = Yes*

What is the upper limit of the target range for their time in range goal? (choose one) *In this question, we present blood glucose values in mmol/L, the common presentation in Canada. If you use mg/dl, here are some equivalent numbers: 70 mg/dl = 3.9 mmol/L; 80 mg/dl = 4.4 mmol/L; 90 mg/dl = 5.0 mmol/L; 100 mg/dl = 5.6 mmol/L; 110 mg/dl = 6.1 mmol/L; 120 mg/dl = 6.7 mmol/L; 130 mg/dl = 7.2 mmol/L; 140 mg/dl = 7.8 mmol/L; 150 mg/dl = 8.3 mmol/L; 160*

*mg/dl = 8.9 mmol/L; 170 mg/dl = 9.4 mmol/L; 180 mg/dl = 10.0 mmol/L; 190 mg/dl = 10.6 mmol/L; 200 mg/dl = 11.1 mmol/L*

- ○ under 5.0 mmol/L (5)
- ○ 5.0 mmol/L (143)
- ○ 5.1 mmol/L (144)
- ○ 5.2 mmol/L (145)
- ○ 5.3 mmol/L (146)
- ○ 5.4 mmol/L (147)
- ○ 5.5 mmol/L (148)
- ○ 5.6 mmol/L (149)
- ○ 5.7 mmol/L (150)
- ○ 5.8 mmol/L (151)
- ○ 5.9 mmol/L (152)
- ○ 6.0 mmol/L (153)
- ○ 6.1 mmol/L (154)
- ○ 6.2 mmol/L (155)
- ○ 6.3 mmol/L (156)
- ○ 6.4 mmol/L (157)
- ○ 6.5 mmol/L (158)
- ○ 6.6 mmol/L (159)
- ○ 6.7 mmol/L (160)
- ○ 6.8 mmol/L (161)

○ 6.9 mmol/L (162)

○ 7.0 mmol/L (163)

○ 7.1 mmol/L (164)

○ 7.2 mmol/L (165)

○ 7.3 mmol/L (166)

○ 7.4 mmol/L (167)

○ 7.5 mmol/L (168)

○ 7.6 mmol/L (169)

○ 7.7 mmol/L (170)

○ 7.8 mmol/L (171)

○ 7.9 mmol/L (172)

○ 8.0 mmol/L (173)

○ 8.1 mmol/L (174)

○ 8.2 mmol/L (175)

○ 8.3 mmol/L (176)

○ 8.4 mmol/L (177)

○ 8.5 mmol/L (178)

○ 8.6 mmol/L (179)

○ 8.7 mmol/L (180)

○ 8.8 mmol/L (181)

○ 8.9 mmol/L (182)

- 9.0 mmol/L (183)
- 9.1 mmol/L (184)
- 9.2 mmol/L (185)
- 9.3 mmol/L (186)
- 9.4 mmol/L (187)
- 9.5 mmol/L (188)
- 9.6 mmol/L (189)
- 9.7 mmol/L (190)
- 9.8 mmol/L (191)
- 9.9 mmol/L (192)
- 10.0 mmol/L (consensus value used in Ambulatory Glucose Profile reports) (193)
- 10.1 mmol/L (194)
- 10.2 mmol/L (195)
- 10.3 mmol/L (196)
- 10.4 mmol/L (197)
- 10.5 mmol/L (198)
- 10.6 mmol/L (199)
- 10.7 mmol/L (200)
- 10.8 mmol/L (201)
- 10.9 mmol/L (202)
- 11.0 mmol/L (203)

○ 11.1 mmol/L (204)

○ 11.2 mmol/L (205)

○ 11.3 mmol/L (206)

○ 11.4 mmol/L (207)

○ 11.5 mmol/L (208)

○ 11.6 mmol/L (209)

○ 11.7 mmol/L (210)

○ 11.8 mmol/L (211)

○ 11.9 mmol/L (212)

○ 12.0 mmol/L (213)

○ 12.1 mmol/L (214)

○ 12.2 mmol/L (215)

○ 12.3 mmol/L (216)

○ 12.4 mmol/L (217)

○ 12.5 mmol/L (218)

○ 12.6 mmol/L (219)

○ 12.7 mmol/L (220)

○ 12.8 mmol/L (221)

○ 12.9 mmol/L (222)

○ 13.0 mmol/L (223)

○ 13.1 mmol/L (224)

○ 13.2 mmol/L (225)

○ 13.3 mmol/L (226)

○ 13.4 mmol/L (227)

○ 13.5 mmol/L (228)

○ 13.6 mmol/L (229)

○ 13.7 mmol/L (230)

○ 13.8 mmol/L (231)

○ 13.9 mmol/L (232)

○ 14.0 mmol/L (233)

○ 14.1 mmol/L (234)

○ 14.2 mmol/L (235)

○ 14.3 mmol/L (236)

○ 14.4 mmol/L (237)

○ 14.5 mmol/L (238)

○ 14.6 mmol/L (239)

○ 14.7 mmol/L (240)

○ 14.8 mmol/L (241)

○ 14.9 mmol/L (242)

○ 15.0 mmol/L (243)

○ above 15.0 mmol/L (244)

○ I don’t know (245)

◯ I prefer not to answer (246)

---

*Display this question:*

*If Does your child have a specific time in range goal? (choose one) = Yes*

Comments about your child's target range (optional)

________________________________________________________________

________________________________________________________________

________________________________________________________________

________________________________________________________________

________________________________________________________________

---

*Display this question:*

*If Does your child have a specific time in range goal? (choose one) = Yes*

What is the goal for the percentage of time they spend in their target range? The goal is to spend at least this percent of time in their target range:

- ○ 0% (4)
- ○ 1% (5)
- ○ 2% (6)
- ○ 3% (7)
- ○ 4% (8)
- ○ 5% (9)
- ○ 6% (10)
- ○ 7% (11)
- ○ 8% (12)
- ○ 9% (13)
- ○ 10% (14)
- ○ 11% (15)
- ○ 12% (16)
- ○ 13% (17)
- ○ 14% (18)
- ○ 15% (19)
- ○ 16% (20)
- ○ 17% (21)
- ○ 18% (22)
- ○ 19% (23)

- 20%  (24)
- 21%  (25)
- 22%  (26)
- 23%  (27)
- 24%  (28)
- 25%  (29)
- 26%  (30)
- 27%  (31)
- 28%  (32)
- 29%  (33)
- 30%  (34)
- 31%  (35)
- 32%  (36)
- 33%  (37)
- 34%  (38)
- 35%  (39)
- 36%  (40)
- 37%  (41)
- 38%  (42)
- 39%  (43)
- 40%  (44)

○ 41% (45)

○ 42% (46)

○ 43% (47)

○ 44% (48)

○ 45% (49)

○ 46% (50)

○ 47% (51)

○ 48% (52)

○ 49% (53)

○ 50% (54)

○ 51% (55)

○ 52% (56)

○ 53% (57)

○ 54% (58)

○ 55% (59)

○ 56% (60)

○ 57% (61)

○ 58% (62)

○ 59% (63)

○ 60% (64)

○ 61% (65)

- 62% (66)
- 63% (67)
- 64% (68)
- 65% (69)
- 66% (70)
- 67% (71)
- 68% (72)
- 69% (73)
- 70% (consensus value used in Ambulatory Glucose Profile reports) (74)
- 71% (75)
- 72% (76)
- 73% (77)
- 74% (78)
- 75% (79)
- 76% (80)
- 77% (81)
- 78% (82)
- 79% (83)
- 80% (84)
- 81% (85)
- 82% (86)

- 83% (87)
- 84% (88)
- 85% (89)
- 86% (90)
- 87% (91)
- 88% (92)
- 89% (93)
- 90% (94)
- 91% (95)
- 92% (96)
- 93% (97)
- 94% (98)
- 95% (99)
- 96% (100)
- 97% (101)
- 98% (102)
- 99% (103)
- 100% (104)
- I don’t know (105)
- I prefer not to answer (106)

*Display this question:*

*If Does your child have a specific time in range goal? (choose one) = Yes*

Who set this goal? (choose one)

- ◯ My child's health care provider (endocrinologist, other doctor, diabetes educator, etc.) set the goal. (7)
- ◯ My child's health care provider and our family set the goal together. (8)
- ◯ Our family set the goal. (9)
- ◯ Other (10)
- ◯ I don't know (11)
- ◯ I prefer not to answer (12)

---

*Display this question:*

*If How many children under age 18 with type 1 diabetes (T1D) or a similar type of diabetes (such as... = 3*

*Or How many children under age 18 with type 1 diabetes (T1D) or a similar type of diabetes (such as... = More than 3*

Has your child had a hemoglobin A1c test within the past 6 months? (choose one)

- ◯ Yes (1)
- ◯ No (2)
- ◯ I don't know (3)
- ◯ I prefer not to answer (4)

---

*Display this question:*

*If Has your child had a hemoglobin A1c test within the past 6 months? (choose one) = Yes*

What was their most recent hemoglobin A1c result?

- under 4.0% (under 20 mmol/mol) (346)
- 4.0% (20 mmol/mol) (347)
- 4.1% (21 mmol/mol) (348)
- 4.2% (22 mmol/mol) (349)
- 4.3% (23 mmol/mol) (350)
- 4.4% (25 mmol/mol) (351)
- 4.5% (26 mmol/mol) (352)
- 4.6% (27 mmol/mol) (353)
- 4.7% (28 mmol/mol) (354)
- 4.8% (29 mmol/mol) (355)
- 4.9% (30 mmol/mol) (356)
- 5.0% (31 mmol/mol) (357)
- 5.1% (32 mmol/mol) (358)
- 5.2% (33 mmol/mol) (359)
- 5.3% (34 mmol/mol) (360)
- 5.4% (36 mmol/mol) (361)
- 5.5% (37 mmol/mol) (362)
- 5.6% (38 mmol/mol) (363)
- 5.7% (39 mmol/mol) (364)
- 5.8% (40 mmol/mol) (365)
- 5.9% (41 mmol/mol) (366)

- 6.0% (42 mmol/mol)  (367)
- 6.1% (43 mmol/mol)  (368)
- 6.2% (44 mmol/mol)  (369)
- 6.3% (45 mmol/mol)  (370)
- 6.4% (46 mmol/mol)  (371)
- 6.5% (48 mmol/mol)  (372)
- 6.6% (49 mmol/mol)  (373)
- 6.7% (50 mmol/mol)  (374)
- 6.8% (51 mmol/mol)  (375)
- 6.9% (52 mmol/mol)  (376)
- 7.0% (53 mmol/mol)  (377)
- 7.1% (54 mmol/mol)  (378)
- 7.2% (55 mmol/mol)  (379)
- 7.3% (56 mmol/mol)  (380)
- 7.4% (57 mmol/mol)  (381)
- 7.5% (58 mmol/mol)  (382)
- 7.6% (59 mmol/mol)  (383)
- 7.7% (60 mmol/mol)  (384)
- 7.8% (62 mmol/mol)  (385)
- 7.9% (63 mmol/mol)  (386)
- 8.0% (64 mmol/mol)  (387)

- 8.1% (65 mmol/mol) (388)
- 8.2% (66 mmol/mol) (389)
- 8.3% (67 mmol/mol) (390)
- 8.4% (68 mmol/mol) (391)
- 8.5% (69 mmol/mol) (392)
- 8.6% (70 mmol/mol) (393)
- 8.7% (72 mmol/mol) (394)
- 8.8% (73 mmol/mol) (395)
- 8.9% (74 mmol/mol) (396)
- 9.0% (75 mmol/mol) (397)
- 9.1% (76 mmol/mol) (398)
- 9.2% (77 mmol/mol) (399)
- 9.3% (78 mmol/mol) (400)
- 9.4% (79 mmol/mol) (401)
- 9.5% (80 mmol/mol) (402)
- 9.6% (81 mmol/mol) (403)
- 9.7% (83 mmol/mol) (404)
- 9.8% (84 mmol/mol) (405)
- 9.9% (85 mmol/mol) (406)
- 10.0% (86 mmol/mol) (407)
- 10.1% (87 mmol/mol) (408)

○ 10.2% (88 mmol/mol) (409)

○ 10.3% (89 mmol/mol) (410)

○ 10.4% (90 mmol/mol) (411)

○ 10.5% (91 mmol/mol) (412)

○ 10.6% (92 mmol/mol) (413)

○ 10.7% (93 mmol/mol) (414)

○ 10.8% (94 mmol/mol) (415)

○ 10.9% (95 mmol/mol) (416)

○ 11.0% (97 mmol/mol) (417)

○ 11.1% (98 mmol/mol) (418)

○ 11.2% (99 mmol/mol) (419)

○ 11.3% (100 mmol/mol) (420)

○ 11.4% (101 mmol/mol) (421)

○ 11.5% (102 mmol/mol) (422)

○ 11.6% (103 mmol/mol) (423)

○ 11.7% (104 mmol/mol) (424)

○ 11.8% (105 mmol/mol) (425)

○ 11.9% (106 mmol/mol) (426)

○ 12.0% (108 mmol/mol) (427)

○ 12.1% (109 mmol/mol) (428)

○ 12.2% (110 mmol/mol) (429)

○ 12.3% (111 mmol/mol) (430)

○ 12.4% (112 mmol/mol) (431)

○ 12.5% (113 mmol/mol) (432)

○ 12.6% (114 mmol/mol) (433)

○ 12.7% (115 mmol/mol) (434)

○ 12.8% (116 mmol/mol) (435)

○ 12.9% (117 mmol/mol) (436)

○ 13.0% (119 mmol/mol) (437)

○ 13.1% (120 mmol/mol) (438)

○ 13.2% (121 mmol/mol) (439)

○ 13.3% (122 mmol/mol) (440)

○ 13.4% (123 mmol/mol) (441)

○ 13.5% (124 mmol/mol) (442)

○ 13.6% (125 mmol/mol) (443)

○ 13.7% (126 mmol/mol) (444)

○ 13.8% (127 mmol/mol) (445)

○ 13.9% (128 mmol/mol) (446)

○ 14.0% (130 mmol/mol) (447)

○ 14.1% (131 mmol/mol) (448)

○ 14.2% (132 mmol/mol) (449)

○ 14.3% (133 mmol/mol) (450)

○ 14.4% (134 mmol/mol) (451)

○ 14.5% (135 mmol/mol) (452)

○ 14.6% (136 mmol/mol) (453)

○ 14.7% (137 mmol/mol) (454)

○ 14.8% (138 mmol/mol) (455)

○ 14.9% (139 mmol/mol) (456)

○ 15.0% (140 mmol/mol) (457)

○ Above 15.0% (above 140 mmol/mol) (458)

○ I don't know (459)

○ I prefer not to answer (460)

---

*Display this question:*

*If Has your child had a hemoglobin A1c test within the past 6 months? (choose one) = Yes*

When was this hemoglobin A1c test done?

- This month (1)
- Last month (2)
- 2 months ago (3)
- 3 months ago (4)
- 4 months ago (5)
- 5 months ago (6)
- 6 months ago (7)
- More than 6 months ago (8)
- I don't know (9)
- I prefer not to answer (10)

*Display this question:*

*If Do you know how to check Time in Range in your child's or children's CGM app(s)? (choose one) = Yes*

*And If*

*How many children under age 18 with type 1 diabetes (T1D) or a similar type of diabetes (such as... = 3*

*Or How many children under age 18 with type 1 diabetes (T1D) or a similar type of diabetes (such as... = More than 3*

Over the past 90 days, what was your child's Time in Range for the standard range 3.9 mmol/L - 10 mmol/L?

- ○ 0% (1)
- ○ 1% (2)
- ○ 2% (3)
- ○ 3% (4)
- ○ 4% (5)
- ○ 5% (6)
- ○ 6% (7)
- ○ 7% (8)
- ○ 8% (9)
- ○ 9% (10)
- ○ 10% (11)
- ○ 11% (12)
- ○ 12% (13)
- ○ 13% (14)
- ○ 14% (15)
- ○ 15% (16)
- ○ 16% (17)
- ○ 17% (18)
- ○ 18% (19)
- ○ 19% (20)

- 20% (21)
- 21% (22)
- 22% (23)
- 23% (24)
- 24% (25)
- 25% (26)
- 26% (27)
- 27% (28)
- 28% (29)
- 29% (30)
- 30% (31)
- 31% (32)
- 32% (33)
- 33% (34)
- 34% (35)
- 35% (36)
- 36% (37)
- 37% (38)
- 38% (39)
- 39% (40)
- 40% (41)

○ 41% (42)

○ 42% (43)

○ 43% (44)

○ 44% (45)

○ 45% (46)

○ 46% (47)

○ 47% (48)

○ 48% (49)

○ 49% (50)

○ 50% (51)

○ 51% (52)

○ 52% (53)

○ 53% (54)

○ 54% (55)

○ 55% (56)

○ 56% (57)

○ 57% (58)

○ 58% (59)

○ 59% (60)

○ 60% (61)

○ 61% (62)

- ○ 62% (63)
- ○ 63% (64)
- ○ 64% (65)
- ○ 65% (66)
- ○ 66% (67)
- ○ 67% (68)
- ○ 68% (69)
- ○ 69% (70)
- ○ 70% (71)
- ○ 71% (72)
- ○ 72% (73)
- ○ 73% (74)
- ○ 74% (75)
- ○ 75% (76)
- ○ 76% (77)
- ○ 77% (78)
- ○ 78% (79)
- ○ 79% (80)
- ○ 80% (81)
- ○ 81% (82)
- ○ 82% (83)

- 83%  (84)
- 84%  (85)
- 85%  (86)
- 86%  (87)
- 87%  (88)
- 88%  (89)
- 89%  (90)
- 90%  (91)
- 91%  (92)
- 92%  (93)
- 93%  (94)
- 94%  (95)
- 95%  (96)
- 96%  (97)
- 97%  (98)
- 98%  (99)
- 99%  (100)
- 100%  (101)
- I don’t know  (102)
- I prefer not to answer  (103)

*Display this question:*

*If How many children under age 18 with type 1 diabetes (T1D) or a similar type of diabetes (such as... = 3*

*Or How many children under age 18 with type 1 diabetes (T1D) or a similar type of diabetes (such as... = More than 3*

Over the past 6 months, has your child had any **severe** hypoglycemic (low blood sugar) events in which they required help from someone else? (choose one)

- ○ Yes (1)
- ○ No (2)
- ○ I don't know (3)
- ○ I prefer not to answer (4)

---

*Display this question:*

*If Over the past 6 months, has your child had any severe hypoglycemic (low blood sugar) events in wh... = Yes*

When your child required help from someone else for the severe hypoglycemic (low blood sugar) event(s), did that help include: (check all that apply)

- ▢ Emergency services (e.g., someone called 911) (1)
- ▢ Glucagon (e.g., someone gave your child glucagon by needle or by nose) (2)
- ▢ Hospitalization (your child received treatment at a hospital) (3)
- ▢ Someone getting low treatment (e.g., juice) and bringing it to your child (4)
- ▢ I don't know (5)
- ▢ I prefer not to answer (6)

**End of Block: Health care & access**

Start of Block: Physical activity and insurance

*Display this question:*

*If Are you the parent or guardian of one or more children under age 18 with type 1 diabetes (T1D) or... = Yes*

*And How old is/are your child/children with type 1 diabetes (T1D) or a similar type of diabetes, such... = Under 1 year old*

**Over the past 6 months**, how frequently has your child with T1D (or similar type of diabetes) and aged under 1 year old: (check all that apply)

| | Always (1) | Often (2) | Sometimes (3) | Rarely (4) | Never (5) | I don’t know (6) | I prefer not to answer (7) |
|---|---|---|---|---|---|---|---|
| Gotten 12-16 hours (if 4-11 months old) or 14-17 hours (if 0-3 months old) of good-quality sleep per 24-hour period (including naps) (1) | ○ | ○ | ○ | ○ | ○ | ○ | ○ |
| Been physically active several times each day in a variety of ways, particularly through interactive floor-based play (2) | ○ | ○ | ○ | ○ | ○ | ○ | ○ |
| Been restrained for more than 1 hour at a time (for example, in a stroller or car seat) or sat for extended periods (3) | ○ | ○ | ○ | ○ | ○ | ○ | ○ |

| | | | | | | | |
|---|---|---|---|---|---|---|---|
| Had any screen time (for example, watching tv or movies) (4) | ○ | ○ | ○ | ○ | ○ | ○ | ○ |
| Spent time sitting or laying down reading or storytelling with a caregiver (5) | ○ | ○ | ○ | ○ | ○ | ○ | ○ |

Page Break

*Display this question:*

*If Are you the parent or guardian of one or more children under age 18 with type 1 diabetes (T1D) or... = Yes*

*And How old is/are your child/children with type 1 diabetes (T1D) or a similar type of diabetes, such... = 1 year old*

**Over the past 6 months**, how frequently has your child with T1D (or similar type of diabetes) and aged 1 year old: (check all that apply)

| | Always (1) | Often (2) | Sometimes (3) | Rarely (4) | Never (5) | I don't know (6) | I prefer not to answer (7) |
|---|---|---|---|---|---|---|---|
| Gotten 11-14 hours of good-quality sleep per 24-hour period (including naps) with consistent bed and wake-up times (1) | ○ | ○ | ○ | ○ | ○ | ○ | ○ |
| Spent at least 180 minutes (3 hours) per day in a variety of physical activities (climbing, dancing, running around) at any intensity, including energetic play, spread throughout the day (2) | ○ | ○ | ○ | ○ | ○ | ○ | ○ |

| | | | | | | | |
|---|---|---|---|---|---|---|---|
| Been restrained for more than 1 hour at a time (for example, in a stroller or car seat) or sat for extended periods (3) | ○ | ○ | ○ | ○ | ○ | ○ | ○ |
| Had any sedentary screen time (for example, watching tv or movies while sitting or laying down) (4) | ○ | ○ | ○ | ○ | ○ | ○ | ○ |
| Spent time sitting or laying down reading or storytelling with a caregiver (5) | ○ | ○ | ○ | ○ | ○ | ○ | ○ |

Page Break

*Display this question:*

*If Are you the parent or guardian of one or more children under age 18 with type 1 diabetes (T1D) or... = Yes*

*And How old is/are your child/children with type 1 diabetes (T1D) or a similar type of diabetes, such... = 2 years old*

**Over the past 6 months**, how frequently has your child with T1D (or similar type of diabetes) and aged 2 years old: (check all that apply)

| | Always (1) | Often (2) | Sometimes (3) | Rarely (4) | Never (5) | I don’t know (6) | I prefer not to answer (7) |
|---|---|---|---|---|---|---|---|
| Gotten 11-14 hours of good-quality sleep per 24-hour period (including naps) with consistent bed and wake-up times (1) | o | o | o | o | o | o | o |
| Spent at least 180 minutes (3 hours) per day in a variety of physical activities (climbing, dancing, running around) at any intensity, including energetic play, spread throughout the day (2) | o | o | o | o | o | o | o |

| | | | | | | | |
|---|---|---|---|---|---|---|---|
| Been restrained for more than 1 hour at a time (for example, in a stroller or car seat) or sat for extended periods (3) | ○ | ○ | ○ | ○ | ○ | ○ | ○ |
| Had more than 1 hour per day of sedentary screen time (for example, watching tv or movies while sitting or laying down) (4) | ○ | ○ | ○ | ○ | ○ | ○ | ○ |
| Spent time sitting or laying down reading or storytelling with a caregiver (5) | ○ | ○ | ○ | ○ | ○ | ○ | ○ |

Page Break

*Display this question:*

*If Are you the parent or guardian of one or more children under age 18 with type 1 diabetes (T1D) or... = Yes*

*And How old is/are your child/children with type 1 diabetes (T1D) or a similar type of diabetes, such... = 3-4 years old*

**Over the past 6 months**, how frequently has your child with T1D (or similar type of diabetes) and aged 3-4 years old: (check all that apply)

| | Always (1) | Often (2) | Sometimes (3) | Rarely (4) | Never (5) | I don't know (6) | I prefer not to answer (7) |
|---|---|---|---|---|---|---|---|
| Gotten 10-13 hours of good-quality sleep per 24-hour period (either all at night or mostly at night with a daytime nap) with consistent bed and wake-up times (1) | ○ | ○ | ○ | ○ | ○ | ○ | ○ |
| Spent at least 180 minutes (3 hours) per day in a variety of physical activities (climbing, dancing, running around) spread throughout the day, of which at least 60 minutes (1 hour) was energetic play (2) | ○ | ○ | ○ | ○ | ○ | ○ | ○ |

| | | | | | | | |
|---|---|---|---|---|---|---|---|
| Been restrained for more than 1 hour at a time (for example, in a stroller or car seat) or sat for extended periods (3) | ○ | ○ | ○ | ○ | ○ | ○ | ○ |
| Had more than 1 hour per day of sedentary screen time (for example, watching tv or movies while sitting or laying down) (4) | ○ | ○ | ○ | ○ | ○ | ○ | ○ |
| Spent time sitting or laying down reading or storytelling with a caregiver (5) | ○ | ○ | ○ | ○ | ○ | ○ | ○ |

Page Break

*Display this question:*

*If Are you the parent or guardian of one or more children under age 18 with type 1 diabetes (T1D) or... = Yes*

*And If*

*How old is/are your child/children with type 1 diabetes (T1D) or a similar type of diabetes, such... = 13 years old*

*Or How old is/are your child/children with type 1 diabetes (T1D) or a similar type of diabetes, such... = 5-12 years old*

**Over the past 6 months**, how frequently has your child with T1D (or similar type of diabetes) and aged 5-13 years old: (check all that apply)

| | Always (1) | Often (2) | Sometimes (3) | Rarely (4) | Never (5) | I don't know (6) | I prefer not to answer (7) |
|---|---|---|---|---|---|---|---|
| Gotten 9-11 hours of sleep per night, with consistent bed and wake-up times (1) | ○ | ○ | ○ | ○ | ○ | ○ | ○ |
| Gotten at least 60 minutes (1 hour) per day of moderate to vigorous physical activity (anything that makes them out of breath) (2) | ○ | ○ | ○ | ○ | ○ | ○ | ○ |
| Done vigorous physical activities (activities that make them very out of breath) at least 3 days each week (3) | ○ | ○ | ○ | ○ | ○ | ○ | ○ |

| | | | | | | | |
|---|---|---|---|---|---|---|---|
| Done muscle and bone-strengthening activities (for example, activities that involve jumping on hard surfaces or lifting things that are heavy for them) 3 days each week (4) | ○ | ○ | ○ | ○ | ○ | ○ | ○ |
| Had several hours each day doing structured and unstructured light physical activities (for example, walking, standing) per day (5) | ○ | ○ | ○ | ○ | ○ | ○ | ○ |
| Sat for extended periods (for example, sitting without moving in school or in front of a screen) (6) | ○ | ○ | ○ | ○ | ○ | ○ | ○ |
| Had more than 2 hours per day of recreational screen time (for example, tv, movies, social media, video games) (7) | ○ | ○ | ○ | ○ | ○ | ○ | ○ |

Page Break

*Display this question:*

*If Are you the parent or guardian of one or more children under age 18 with type 1 diabetes (T1D) or... = Yes*

*And How old is/are your child/children with type 1 diabetes (T1D) or a similar type of diabetes, such... = 14-17 years old*

**Over the past 6 months**, how frequently has your child with T1D (or similar type of diabetes) and aged 14-17 years: (check all that apply)

| | Always (1) | Often (2) | Sometimes (3) | Rarely (4) | Never (5) | I don't know (6) | I prefer not to answer (7) |
|---|---|---|---|---|---|---|---|
| Gotten 8-10 hours of sleep per night, with consistent bed and wake-up times (1) | o | o | o | o | o | o | o |
| Gotten at least 60 minutes (1 hour) per day of moderate to vigorous physical activity (anything that makes them out of breath) (2) | o | o | o | o | o | o | o |
| Done vigorous physical activities (activities that make them very out of breath) at least 3 days each week (3) | o | o | o | o | o | o | o |

| | | | | | | | |
|---|---|---|---|---|---|---|---|
| Done muscle and bone-strengthening activities (for example, activities that involve jumping on hard surfaces or lifting things that are heavy for them) 3 days each week (4) | ○ | ○ | ○ | ○ | ○ | ○ | ○ |
| Had several hours each day doing structured and unstructured light physical activities (for example, walking, standing) per day (5) | ○ | ○ | ○ | ○ | ○ | ○ | ○ |
| Sat for extended periods (for example, sitting without moving in school or in front of a screen) (6) | ○ | ○ | ○ | ○ | ○ | ○ | ○ |
| Had more than 2 hours per day of recreational screen time (for example, tv, movies, social media, video games) (7) | ○ | ○ | ○ | ○ | ○ | ○ | ○ |

Page Break

*Display this question:*

*If Do you yourself have type 1 diabetes (T1D) or a similar type of diabetes, such as LADA, MODY, or... = Yes*

*And To which age group do you belong? (choose one) = 18-64 years old*

**Over the past 6 months**, how frequently have you: (check all that apply)

| | Always (1) | Often (2) | Sometimes (3) | Rarely (4) | Never (5) | I don't know (6) | I prefer not to answer (7) |
|---|---|---|---|---|---|---|---|
| Gotten 7 to 9 hours of good-quality sleep per day on a regular basis, with consistent bed and wake-up times (1) | ○ | ○ | ○ | ○ | ○ | ○ | ○ |
| Done moderate to vigorous aerobic physical activities (for example: jogging, playing basketball, swimming, or other activities that make you out of breath) for at least 150 minutes (2.5 hours) total over the week (for example, 30 minutes per day, 5 days a week) (2) | ○ | ○ | ○ | ○ | ○ | ○ | ○ |

| | | | | | | | |
|---|---|---|---|---|---|---|---|
| Done muscle strengthening activities (for example, lifting weights or doing bodyweight exercises) using major muscle groups at least twice a week (3) | ○ | ○ | ○ | ○ | ○ | ○ | ○ |
| Had several hours of light physical activities (for example, walking or standing) per day most days (4) | ○ | ○ | ○ | ○ | ○ | ○ | ○ |
| Sat more than 8 hours per day (5) | ○ | ○ | ○ | ○ | ○ | ○ | ○ |
| Had more than 3 hours per day of recreational screen time (for example, tv, movies, social media, video games) (6) | ○ | ○ | ○ | ○ | ○ | ○ | ○ |

| | | | | | | | |
|---|---|---|---|---|---|---|---|
| Broke up long periods of sitting as often as possible (for example, by getting up and stretching, walking to get a glass of water or to use the washrooms) (7) | o | o | o | o | o | o | o |

Page Break

*Display this question:*

*If Do you yourself have type 1 diabetes (T1D) or a similar type of diabetes, such as LADA, MODY, or... = Yes*

*And To which age group do you belong? (choose one) = 65+ years old*

**Over the past 6 months**, how frequently have you: (check all that apply)

| | Always (1) | Often (2) | Sometimes (3) | Rarely (4) | Never (5) | I don't know (6) | I prefer not to answer (7) |
|---|---|---|---|---|---|---|---|
| Gotten 7 to 8 hours of good-quality sleep per day on a regular basis, with consistent bed and wake-up times (1) | o | o | o | o | o | o | o |
| Done moderate to vigorous aerobic physical activities (for example: jogging, playing basketball, swimming, or other activities that make you out of breath) for 150 minutes (2.5 hours) total each week (for example, 30 minutes per day, 5 days a week) or more (2) | o | o | o | o | o | o | o |

| | | | | | | | |
|---|---|---|---|---|---|---|---|
| Done muscle strengthening activities (for example, lifting weights or doing bodyweight exercises) using major muscle groups twice a week or more (3) | ○ | ○ | ○ | ○ | ○ | ○ | ○ |
| Done physical activities that challenge balance (for example, standing on one leg, tai chi) (4) | ○ | ○ | ○ | ○ | ○ | ○ | ○ |
| Had several hours of light physical activities (for example, walking or standing) per day (5) | ○ | ○ | ○ | ○ | ○ | ○ | ○ |
| Sat more than 8 hours per day (6) | ○ | ○ | ○ | ○ | ○ | ○ | ○ |
| Had more than 3 hours per day of recreational screen time (for example, tv, movies, social media, video games) (7) | ○ | ○ | ○ | ○ | ○ | ○ | ○ |

| | | | | | | | |
|---|---|---|---|---|---|---|---|
| Broke up long periods of sitting as often as possible (for example, by getting up and stretching, walking to get a glass of water or to use the washrooms) (8) | o | o | o | o | o | o | o |

Page Break

*Display this question:*

*If Do you yourself have type 1 diabetes (T1D) or a similar type of diabetes, such as LADA, MODY, or... = Yes*

Sometimes it can be easy to access tools and supports for managing diabetes. Other times, there are obstacles in the way (for example, lack of funding or long waiting lists) and it can be hard. How easy or hard does it feel for you to get:

| | Impossible (1) | Extremely hard (2) | Hard (3) | Neither hard nor easy (4) | Easy (5) | Extremely easy (6) | Not applicable or I don't know (7) | I prefer not to answer (8) |
|---|---|---|---|---|---|---|---|---|
| Appointments with diabetes specialists (doctors, diabetes educators, dietitians) (1) | ○ | ○ | ○ | ○ | ○ | ○ | ○ | ○ |
| Blood test meter (2) | ○ | ○ | ○ | ○ | ○ | ○ | ○ | ○ |
| Blood test strips (3) | ○ | ○ | ○ | ○ | ○ | ○ | ○ | ○ |
| Continuous glucose monitoring system & supplies (Libre, Dexcom, Medtronic, etc.) (4) | ○ | ○ | ○ | ○ | ○ | ○ | ○ | ○ |
| Healthy food (5) | ○ | ○ | ○ | ○ | ○ | ○ | ○ | ○ |
| Helpful diabetes education (6) | ○ | ○ | ○ | ○ | ○ | ○ | ○ | ○ |
| Hypo/low blood sugar treatments (7) | ○ | ○ | ○ | ○ | ○ | ○ | ○ | ○ |
| Insulin (8) | ○ | ○ | ○ | ○ | ○ | ○ | ○ | ○ |

| | | | | | | | | |
|---|---|---|---|---|---|---|---|---|
| Insulin pump (Medtronic, Tandem, Omnipod, Ypsomed, etc.) (9) | o | o | o | o | o | o | o | o |
| Insulin pump supplies (cartridges, infusion sets, etc.) (10) | o | o | o | o | o | o | o | o |
| Ketone testing supplies (blood testing or urine testing) (11) | o | o | o | o | o | o | o | o |
| Lancets (12) | o | o | o | o | o | o | o | o |
| Needles, syringes, or pens for giving insulin (13) | o | o | o | o | o | o | o | o |
| Support from diabetes specialists (doctors, diabetes educators, dietitians) (14) | o | o | o | o | o | o | o | o |
| Support from the people around you (family, friends, etc.) in managing T1D (15) | o | o | o | o | o | o | o | o |

Page Break

Page Break

*Display this question:*

*If Do you yourself have type 1 diabetes (T1D) or a similar type of diabetes, such as LADA, MODY, or... = Yes*

Do you currently have full or partial insurance coverage for the following diabetes-related costs? (check all that apply)

| | I have no coverage (9) | I have partial coverage with some out of pocket costs (10) | I have full (100%) coverage with no out of pocket costs (11) | Not applicable or I don't know (12) | I prefer not to answer (13) |
|---|---|---|---|---|---|
| Blood test meter (9) | o | o | o | o | o |
| Blood test strips (10) | o | o | o | o | o |
| Continuous glucose monitoring system & supplies (Libre, Dexcom, etc.) (11) | o | o | o | o | o |
| Hypo/low blood sugar treatments (12) | o | o | o | o | o |
| Insulin (13) | o | o | o | o | o |
| Insulin pump (Medtronic, Tandem, Omnipod, Ypsomed, etc.) (14) | o | o | o | o | o |
| Insulin pump supplies (cartridges, infusion sets, etc.) (15) | o | o | o | o | o |
| Ketone testing supplies (blood testing or urine testing) (16) | o | o | o | o | o |
| Lancets (17) | o | o | o | o | o |

| | | | | | |
|---|---|---|---|---|---|
| Needles, syringes, or pens for giving insulin (18) | ○ | ○ | ○ | ○ | ○ |

Page Break

Page Break

*Display this question:*

*If Do you currently have full or partial insurance coverage for the following diabetes-related costs... [ I have partial coverage with some out of pocket costs] (Count) > 0*

*Or Do you currently have full or partial insurance coverage for the following diabetes-related costs... [ I have full (100%) coverage with no out of pocket costs] (Count) > 0*

Is your insurance coverage:

- ○ Government insurance (provincial or federal) (8)
- ○ Private insurance (for example, workplace extended health benefits, group plans you purchase, etc.) (9)
- ○ A mix of government and private insurance (10)
- ○ I don’t know (11)
- ○ I prefer not to answer (12)

---

Page Break

Page Break

*Display this question:*

*If Do you yourself have type 1 diabetes (T1D) or a similar type of diabetes, such as LADA, MODY, or... = Yes*

In the past 5 years, have you ever received a federal, provincial, or territorial disability tax credit? (check all that apply)

- ☐ Yes, I have received a provincial or territorial disability tax credit (4)
- ☐ Yes, I have received the federal disability tax credit (5)
- ☐ No, I have not received any disability tax credit (6)
- ☐ I don’t know (7)
- ☐ I prefer not to answer (8)

Page Break

*Display this question:*

*If Are you the parent or guardian of one or more children under age 18 with type 1 diabetes (T1D) or... = Yes*

Sometimes it can be easy to access tools and supports for managing diabetes. Other times, there are obstacles in the way (for example, lack of funding or long waiting lists) and it can be hard. How easy or hard does it feel for your family to get:

| | Impossible (1) | Extremely hard (2) | Hard (3) | Neither hard nor easy (4) | Easy (5) | Extremely easy (6) | Not applicable or I don’t know (7) | I prefer not to answer (8) |
|---|---|---|---|---|---|---|---|---|
| Appointments with diabetes specialists (doctors, diabetes educators, dietitians) (1) | ○ | ○ | ○ | ○ | ○ | ○ | ○ | ○ |
| Blood test meter (2) | ○ | ○ | ○ | ○ | ○ | ○ | ○ | ○ |
| Blood test strips (3) | ○ | ○ | ○ | ○ | ○ | ○ | ○ | ○ |
| Continuous glucose monitoring system & supplies (Libre, Dexcom, Medtronic, etc.) (4) | ○ | ○ | ○ | ○ | ○ | ○ | ○ | ○ |
| Healthy food (5) | ○ | ○ | ○ | ○ | ○ | ○ | ○ | ○ |
| Helpful diabetes education (6) | ○ | ○ | ○ | ○ | ○ | ○ | ○ | ○ |
| Hypo/low blood sugar treatments (7) | ○ | ○ | ○ | ○ | ○ | ○ | ○ | ○ |
| Insulin (8) | ○ | ○ | ○ | ○ | ○ | ○ | ○ | ○ |
| Insulin pump (Medtronic, Tandem, Omnipod, Ypsomed, etc.) (9) | ○ | ○ | ○ | ○ | ○ | ○ | ○ | ○ |

| | | | | | | | | |
|---|---|---|---|---|---|---|---|---|
| Insulin pump supplies (cartridges, infusion sets, etc.) (10) | ○ | ○ | ○ | ○ | ○ | ○ | ○ | ○ |
| Ketone testing supplies (blood testing or urine testing) (11) | ○ | ○ | ○ | ○ | ○ | ○ | ○ | ○ |
| Lancets (12) | ○ | ○ | ○ | ○ | ○ | ○ | ○ | ○ |
| Needles, syringes, or pens for giving insulin (13) | ○ | ○ | ○ | ○ | ○ | ○ | ○ | ○ |
| Support from diabetes specialists (doctors, diabetes educators, dietitians) (14) | ○ | ○ | ○ | ○ | ○ | ○ | ○ | ○ |
| Support from the people around you (family, friends, school/daycare staff, etc.) in managing T1D (15) | ○ | ○ | ○ | ○ | ○ | ○ | ○ | ○ |

*Display this question:*

*If Are you the parent or guardian of one or more children under age 18 with type 1 diabetes (T1D) or... = Yes*

Does your family currently have full or partial insurance coverage for the following diabetes-related costs? (check all that apply)

| | We have no coverage (9) | We have partial coverage with some out of pocket costs (10) | We have full (100%) coverage with no out of pocket costs (11) | Not applicable or I don’t know (12) | I prefer not to answer (13) |
|---|---|---|---|---|---|
| Blood test meter (9) | ○ | ○ | ○ | ○ | ○ |
| Blood test strips (10) | ○ | ○ | ○ | ○ | ○ |
| Continuous glucose monitoring system & supplies (Libre, Dexcom, etc.) (11) | ○ | ○ | ○ | ○ | ○ |
| Hypo/low blood sugar treatments (12) | ○ | ○ | ○ | ○ | ○ |
| Insulin (13) | ○ | ○ | ○ | ○ | ○ |
| Insulin pump (Medtronic, Tandem, Omnipod, Ypsomed, etc.) (14) | ○ | ○ | ○ | ○ | ○ |
| Insulin pump supplies (cartridges, infusion sets, etc.) (15) | ○ | ○ | ○ | ○ | ○ |
| Ketone testing supplies (blood testing or urine testing) (16) | ○ | ○ | ○ | ○ | ○ |
| Lancets (17) | ○ | ○ | ○ | ○ | ○ |

| | | | | | |
|---|---|---|---|---|---|
| Needles, syringes, or pens for giving insulin (18) | ○ | ○ | ○ | ○ | ○ |

*Display this question:*

*If Does your family currently have full or partial insurance coverage for the following diabetes-rel... [ We have partial coverage with some out of pocket costs] (Count) > 0*

*Or Does your family currently have full or partial insurance coverage for the following diabetes-rel... [ We have full (100%) coverage with no out of pocket costs] (Count) > 0*

Is your family insurance coverage:

- ○ Government insurance (provincial or federal) (5)
- ○ Private insurance (for example, workplace extended health benefits, group plans you purchase, etc.) (6)
- ○ A mix of government and private insurance (7)
- ○ I don't know (8)
- ○ I prefer not to answer (9)

*Display this question:*

*If Are you the parent or guardian of one or more children under age 18 with type 1 diabetes (T1D) or... = Yes*

In the past 5 years, has your family ever received a federal, provincial, or territorial disability tax credit? (check all that apply)

- ☐ Yes, we have received a provincial or territorial disability tax credit (4)
- ☐ Yes, we have received the federal disability tax credit (5)
- ☐ No, we have not received any disability tax credit (6)
- ☐ I don't know (7)
- ☐ I prefer not to answer (8)

**End of Block: Physical activity and insurance**

---

**Start of Block: Draw**

**Everyone who participates in the research study will have the option to be entered in a draw for a $250 gift card each time they complete a survey.** Each time you complete a survey, with your permission, your name is entered in the draw and will stay in the draw throughout the multiyear study. This means that the more surveys you complete, the more times your name will be in the draw. We will randomly draw one or more names every 6 months.

Would you like to have your name entered in the draw? (choose one)

- ○ Yes (4)
- ○ No (5)

*Display this question:*

*If Would you like to have your name entered in the draw? (choose one) = Yes*

To contact you if we draw your name to receive a $250 gift card, please provide your email address and a phone number as a backup. We will primarily contact you by email, but if we are unable to reach you by email, we will use your phone number.

- ○ Email: (4) ________________________________________________
- ○ Email (repeat): (5) ________________________________________________
- ○ Phone number: (6) ________________________________________________
- ○ Phone number (repeat): (7) ________________________________________________
- ○ Comments (optional): (8) ________________________________________________

End of Block: Draw

---

Start of Block: downloadreport

Do you want to download a report of your answers today? If you like, we can now show you your responses today. You can keep this file if you like.

- - - - - - - - - - - - - - - - - - - - - - - - - - - - - - - - - - - - - - - -

Do you want to see a report of your answers?

- ○ Yes (1)
- ○ No (2)

End of Block: downloadreport

---
